# Epidemiological and health economic implications of symptom propagation in respiratory pathogens: A mathematical modelling investigation

**DOI:** 10.1101/2023.07.12.23292544

**Authors:** Phoebe Asplin, Matt J. Keeling, Rebecca Mancy, Edward M. Hill

## Abstract

**Background:** Respiratory pathogens inflict a substantial burden on public health and the economy. Although the severity of symptoms caused by these pathogens can vary from asymptomatic to fatal, the factors that determine symptom severity are not fully understood. Correlations in symptoms between infector-infectee pairs, for which evidence is accumulating, can generate large-scale clusters of severe infections that could be devastating to those most at risk, whilst also conceivably leading to chains of mild or asymptomatic infections that generate widespread immunity with minimal cost to public health. Although this effect could be harnessed to amplify the impact of interventions that reduce symptom severity, the mechanistic representation of symptom propagation within mathematical and health economic modelling of respiratory diseases is understudied.

**Methods and Findings:** We propose a novel framework for incorporating different levels of symptom propagation into models of infectious disease transmission via a single parameter, *α*. Varying *α* tunes the model from having no symptom propagation (*α* = 0, as typically assumed) to one where symptoms always propagate (*α* = 1). For parameters corresponding to three respiratory pathogens — seasonal influenza, pandemic influenza and SARS-CoV-2 — we explored how symptom propagation impacted the relative epidemiological and health-economic performance of three interventions, conceptualised as vaccines with different actions: symptom-attenuating (labelled SA), infection-blocking (IB) and infection-blocking admitting only mild breakthrough infections (IB MB).

In the absence of interventions, with fixed underlying epidemiological parameters, stronger symptom propagation increased the proportion of cases that were severe. For SA and IB MB, interventions were more effective at reducing prevalence (all infections and severe cases) for higher strengths of symptom propagation. For IB, symptom propagation had no impact on effectiveness, and for seasonal influenza this intervention type was more effective than SA at reducing severe infections for all strengths of symptom propagation. For pandemic influenza and SARS-CoV-2, at low intervention uptake, SA was more effective than IB for all levels of symptom propagation; for high uptake, SA only became more effective under strong symptom propagation. Health economic assessments found that for SA-type interventions, the amount one could spend on control whilst maintaining a cost-effective intervention (termed threshold unit intervention cost) was very sensitive to the strength of symptom propagation.

**Conclusions:** Overall, the preferred intervention type depended on the combination of the strength of symptom propagation and uptake. Given the importance of determining robust public health responses, we highlight the need to gather further data on symptom propagation, with our modelling framework acting as a template for future analysis.

## 1 Introduction

Respiratory pathogens, of which influenza and SARS-CoV-2 are prominent examples, are those that cause infections in the respiratory tract, and are a major cause of mortality worldwide in high, medium and low income countries [1]. Many respiratory pathogens have demonstrated their capability to cause large-scale epidemics and/or pandemics. For example, seasonal influenza causes annual epidemics which, prior to the COVID-19 pandemic beginning in 2020, were estimated to result in symptomatic infection of 8% of the US population each year on average [2] and around 290,000 to 650,000 deaths globally [3]. Pandemic influenza has also inflicted devastating consequences on global public health; the 1918/19 Spanish flu pandemic is thought to have resulted in 50 million deaths worldwide [4], while the 2009 H1N1 pandemic caused 200,000 deaths in its first year of circulation [5]. Since its emergence in humans in 2019, SARS-CoV-2, the causative agent of COVID-19 disease, has resulted in an estimated number of global deaths exceeding 6.5 million by the end of 2022 [6].

Whilst the serious public health risks posed by respiratory diseases are evident, the resulting outbreaks also come with a considerable economic cost. COVID-19 has had a massive impact on the global economy, with the global cost in 2020 and 2021 estimated to be 14% of 2019 GDP [7]. These alarming valuations were partially due to the high cost of interventions. For example, by September 2021, the UK had spent £17.9bn on the test and trace programme, £13.8bn on the procurement of personal protective equipment and £1.8bn on vaccine and antibody supply [8].

Although the headline statistics on the health burden of these pathogens are somewhat bleak, many respiratory pathogens are capable of causing of range of symptomatic outcomes. Often, infected individuals experience, at worst, only mild symptoms, such as a runny nose, focused in the upper respiratory tract. On the other hand, such pathogens also have the potential to cause severe symptoms, primarily by infecting lower parts of the respiratory tract [9, 10]. Indeed, lower respiratory tract infections account for more than 2.4 million deaths worldwide each year [1] and are a leading cause of global mortality, especially amongst children and elderly people [11].

The COVID-19 pandemic has heralded a paradigm shift in modelling the actions of interventions when assessing public health control strategies, highlighting the importance of symptom severity and leading to an increased understanding of the action of interventions beyond being purely infection-blocking [12, 13]. Vaccines, whilst previously viewed in modelling terms as solely a way to prevent infection, are now being considered to have a dual action of reducing symptom severity [14–16]. Similarly, non-pharmaceutical interventions such as mask-wearing, social distancing and hand washing were previously only thought of as ways to reduce transmission but are now thought to additionally reduce the likelihood of symptomatic infection [17–21].

Preparedness efforts against respiratory disease outbreaks and the contemporary evaluations of intervention effectiveness motivate research into the relationship between the severity of illness, viral load and the transmission routes through which they spread. This research has revealed good biological grounds for investigating symptom propagation for respiratory pathogens, where we define symptom propagation as occurring when the symptoms of an infected individual depend, at least partially, on the symptoms of their infector. Symptom propagation has been documented for Yersinia pestis, the causative agent of plague. Those who develop the more severe form, pneumonic plague, are then able to infect via the aerosolised route, which results in pneumonic plague in those infected [22].

There is growing evidence that symptom propagation occurs for other respiratory pathogens. Prior studies of influenza and SARS-CoV-2, have suggested two pathways through which symptoms may propagate through chains of infection [23, 24]. The first pathway is through a dose-response relationship. Individuals presenting with severe disease tend to shed more viral particles [25–28], meaning that those they infect receive a larger infectious dose, in turn increasing the probability of more severe disease outcomes [24, 29–31]. The second pathway is through differential transmission routes: it is thought that severe disease arises more frequently for aerosol transmission (transmission involving particles smaller than 5*µm*, which are sufficiently small and light to travel on air flows and to enter the lower respiratory tract) than for close contact transmission (transmission involving direct or indirect contact with an infected individual or transmission via large droplets, which are more likely to lodge in the upper respiratory tract) [32]. The association with severity arises because aerosol transmission is more likely to cause infection in the lower respiratory tract, resulting in a higher probability of more severe disease [23, 30, 33]. These studies give evidence for symptom propagation between infector-infectee pairs, but its incorporation into epidemiological models is required to fully appreciate its importance at a larger scale.

Symptom propagation has the potential to create chains of severe infections, resulting in large, intensive outbreaks that could have devastating consequences for groups most at risk; on the other hand, it could result in mild or asymptomatic infection spreading through a population, creating widespread immunity whilst incurring a minimal cost to public health. In light of the observed differential symptom severity that may be experienced for many pathogens, we are interested in exploring the public health ramifications of a relationship between symptom severity of an infected individual and the symptom severity of any subsequent cases caused by onward transmission.

We view the absence of a mechanistic representation of the propagation of symptom sets as a modelling ‘blind spot’, in light of the earlier described biological evidence already giving strong support of it being a notable process for some pathogens [22]. Symptom severity has typically been modelled post-hoc or separately from epidemiological dynamics. For example, it has become commonplace for models to distinguish between asymptomatic and symptomatic infection, but asymptomatic infections are generally assumed to occur with a fixed probability, independent of other infected individuals [34]. An extension to this model has been explored by Paulo *et al.* [35], where the probability of severe disease depended on the proportion of the population infected at the time, although not on their severity. Other models in the literature capture multi-route transmission but do not invoke a relationship between the route of transmission and symptom severity [23, 36–38]. Initial attempts to incorporate symptom propagation into an epidemiological model of a respiratory tract infection can be seen in Earnest [39] and Harris *et al.* [40]; however, work in this area remains rudimentary.

Another aspect meriting greater attention is the impact of symptom propagation on health economic outcomes used to assess cost-effectiveness and help optimise public health strategies. For seasonal influenza, there has been a focus in previous health economic studies on the cost-effectiveness of vaccination scenarios [41, 42]. On the other hand, in the context of COVID-19, although health economic modelling studies have predominantly focused on vaccine rollout [43], there have been evaluations of symptom-dependent interventions, such as comparing the effectiveness of symptomatic versus asymptomatic testing [44] and considering quarantining measures that predominantly target symptomatic individuals [45]. At the time of writing, no work has been done to explore the effect of symptom propagation on health economic outcomes.

In this paper, we develop a mathematical modelling framework that incorporates symptom propagation and apply it to a range of pathogens to investigate the epidemiological and health-economic implications of symptom propagation. First, we develop a generalisable, mechanistic infectious disease transmission model that incorporates different strengths of symptom propagation via a single parameter, which we call *α*. Parameterising this model to capture three representative respiratory tract pathogens of public health concern (seasonal influenza, pandemic influenza and SARS-CoV-2), we conduct numerical experiments to explore the impacts of symptom propagation of different strengths on epidemiological and health economic outcomes, both with no intervention and under three interventions that we conceptualise of as vaccines with different modes of action. As well as symptom severity propagation having important impacts on natural epidemiological dynamics (in the absence of intervention), we found that when interventions were applied symptom propagation acted to amplify the beneficial effects of symptom-attenuating interventions on community-level epidemiological outcomes. These effects became even more stark for cost-effectiveness based assessments. For pathogens where the propagation of symptom severity is an important attribute, our findings motivate the development and use of a new class of models to help identify the most appropriate type of intervention to harness the beneficial attributes of symptom propagation, delivering a reduced burden on public health and more cost-effective control policy.

## 2 Methods

The propagation of symptom severity has been largely neglected in epidemiological modelling, with symptom severity having typically been modelled post-hoc or separately from epidemiological dynamics. With an application to outbreaks of respiratory pathogens of public health concern (namely seasonal influenza, pandemic influenza and SARS-CoV-2), we investigated the impact of the strength of symptom propagation on epidemiological and health economic measures. Our methodology comprised multiple aspects that we detail in turn: (i) a mathematical model of infectious disease transmission that included two symptom severity classes (mild and severe) and a mechanism for symptom propagation (Section 2.1); (ii) incorporation of interventions into the model, with our implementation roughly corresponding to three plausible modes of action of a vaccine (Section 2.2); (iii) approaches to assess the health economic implications of proposed strategies (Section 2.3).

### 2.1 Infectious disease model including symptom severity and propagation

The basis of our infectious disease model is a standard deterministic, compartmental susceptible-exposed-infectious-recovered (SEIR) model, described by a system of ordinary differential equations (ODEs) [46, 47]. We extended the framework by stratifying infections according to two levels of symptom severity, mild and severe, and mechanistically incorporating (the potential for) symptom propagation (Fig. 1).

**Figure 1:**
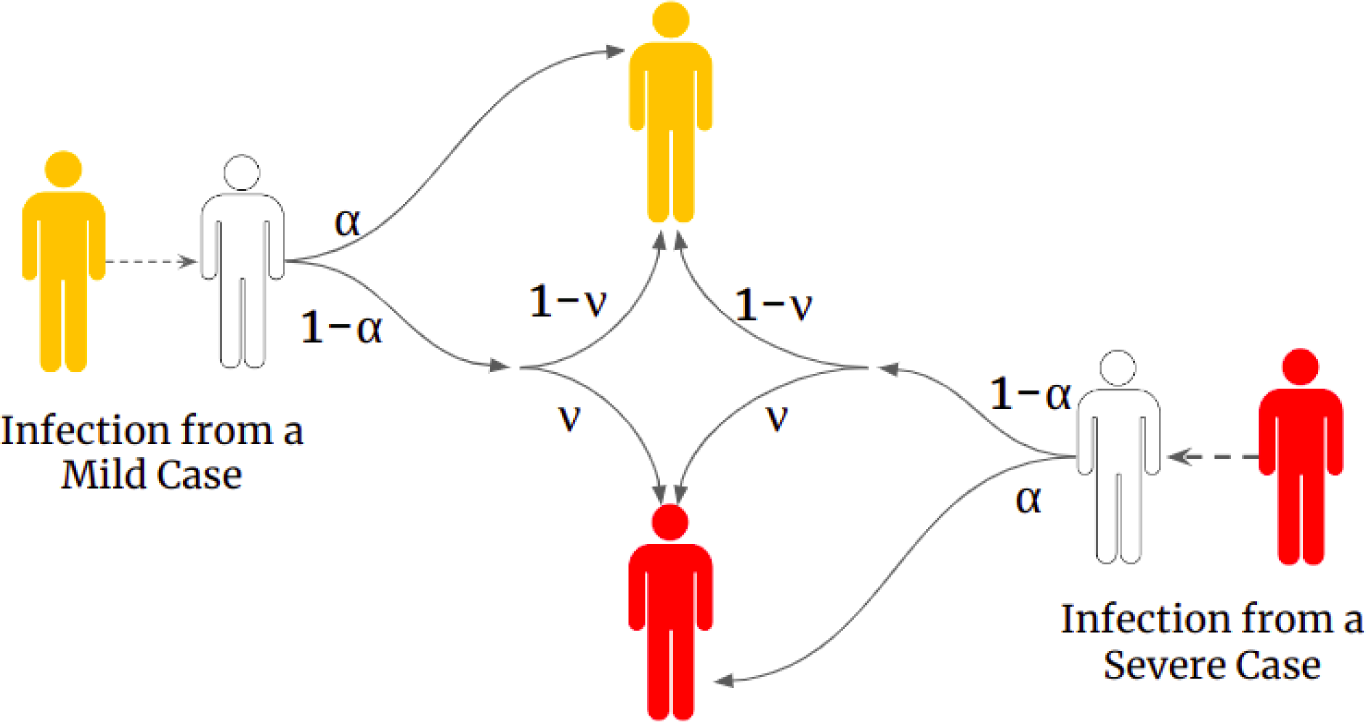
Schematic showing how symptom severity was determined according to the two symptom severity associated parameters, *α* and *ν*. White shaded individuals correspond to those susceptible to infection, yellow shaded individuals correspond to infectious cases with mild severity and red shaded individuals correspond to infectious cases with severe symptoms. The values on the arrows show the corresponding probability. In brief, an infected individual has probability *α* of copying the symptom severity of their infector and a probability 1 *− α* of reverting to the baseline probability of having severe disease, i.e. they developed severe disease with probability *ν*.

We expand below on: (a) the reasoning for the selection of two symptom severity classes; (b) a description of how symptom severity was augmented into the compartmental framework; (c) the transmission dynamics; (d) the mathematical formulation of the symptom propagation mechanism; (e) the collection of ODEs describing the system dynamics (in the absence of interventions); (f) the computational simulations performed to consider the implications of symptom propagation on our three exemplar pathogens (seasonal influenza, pandemic influenza, SARS-CoV-2) in the absence of interventions.

#### (a) Two symptom severity classes

No formal scheme exists for classifying disease severity, with differing interpretations possible across pathogens. For some pathogens, cases are typically stratified into “mild” or “severe”; in the context of influenza, severe disease is generally associated with the development of a cough or fever [34, 48, 49]. For other pathogens, severity is commonly stratified according to “asymptomatic” or “symptomatic” infection, with SARS-CoV-2 being a notable example [40, 50]. Due to the ubiquity of “mild” and “severe” within the literature for respiratory pathogens, we have decided to use this terminology throughout. It should be noted, however, that the parameters chosen for mild COVID-19 disease, such as the infectious period, are taken from estimates for the asymptomatic parameters. We model the propagation of mild and severe symptoms because of the clinical importance of symptom severity for respiratory tract infections but note that the framework can be applied to the propagation of symptom sets in general.

#### (b) Infectious disease model compartments

Under our model structure of compartmentalising the population into susceptible, exposed (infected but not yet infectious), infectious and recovered states, we further separated each of the exposed, infectious and recovered states into two classes representing the two symptom severity levels (“mild” and “severe”). In detail, *E_M_* (*E_S_*) contains individuals exposed to the disease who would go on to develop mild (severe) disease. *I_M_* (*I_S_*) contains individuals who had become infectious and exhibited mild (severe) symptoms. Note that we assumed there was no movement between the severity classes, meaning an individual’s severity would be constant across their exposed and infectious periods. *R_M_* (*R_S_*) contains individuals who had experienced mild (severe) disease and since recovered.

#### (c) Transmission dynamics

We assumed a dependence between disease severity and both the rate of transmission of infection, *β_M_, β_S_* (for “mild” and “severe” cases, respectively), and the recovery rate from infection, *γ_M_, γ_S_*. On the other hand, the incubation period, and thus the rate of becoming infectious, *ɛ*, took the same fixed value for both severity classes due to data suggesting there is limited variation between mild and severe cases [49, 50].

Additionally, we assumed there was no waning immunity after recovery and we ignored demographic processes (natural births and deaths) - as such we are modelling a single epidemic outbreak. In the majority of scenarios, the simulated outbreaks occurred over a short time frame where the impacts of these waning and demographic processes would be negligible. Equally, in the unusual case that outbreaks persisted over a sustained time period (a decade and above), for the purposes of simplicity, we wanted to maintain the focus on the impact of symptom propagation-associated factors.

We explored three disease parameter sets (Table 1), chosen to reflect a range of disease scenarios: influenza-like parameters with *R*_0_ = 1.5 (seasonal influenza), influenza-like parameters with *R*_0_ = 3.0 (pandemic influenza) and early SARS-CoV-2-like parameters with *R*_0_ = 3.0. Specifically, *R*_0_ = 1.5 represented a pathogen that, on average, would spread through a population slowly and require minimal interventions to be suppressed. In contrast, *R*_0_ = 3.0 represented a highly transmissible pathogen with the potential to infect the majority of the population in the absence of strong interventions. These values of *R*_0_ were chosen to reflect estimates in the literature of 1-1.69 for seasonal influenza [51–53], 1.95-3.5 for pandemic influenza [54–56] and 2.43-3.60 for wild-type SARS-CoV-2 [57–59]. To obtain these fixed values of *R*_0_, we computed the required value of the transmission rate *β* for each value of *α* by deriving an equation for *R*_0_ using the next-generation matrix approach (see Section S1). To better align with approaches taken when analysing real-world infections, we chose to fix *R*_0_ as opposed to fixing the value of beta; *R*_0_ is the parameter most likely to be known from available empirical measurements, with other model parameters (in this case *β*) inferred to generate the measured *R*_0_ value. Our analysis with the value of *β* fixed instead of *R*_0_ can be found in Section S2 (Figs. S1 and S2).

**Table 1:**
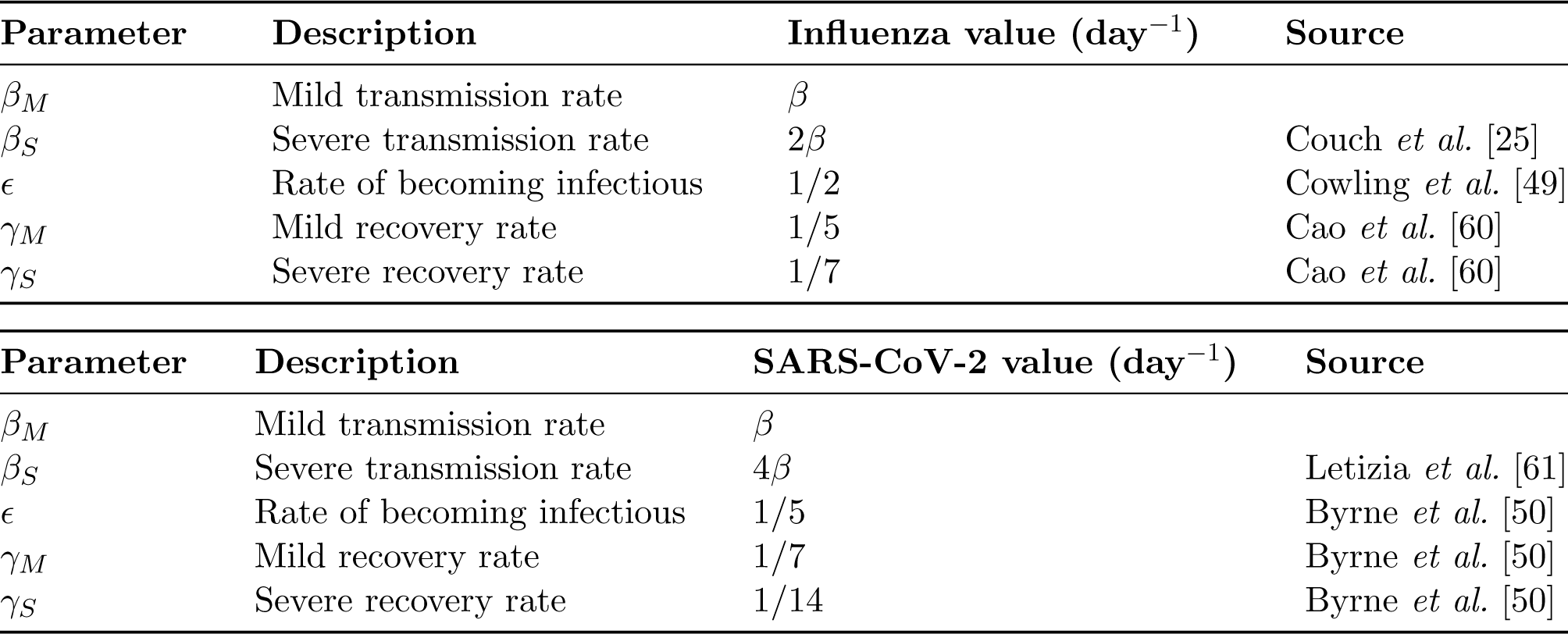
Epidemiological parameter values for the (seasonal and pandemic) influenza and SARS-CoV-2 scenarios. In each parameterisation, we calibrated the *β* value to acquire the stated value of *R*_0_ in the respective scenario. All rates have a unit of ‘per day’ (day*^−^*^1^). **(Top)** Parameters used for the two influenza scenarios: influenza-like parameters with *R*_0_ = 1.5 (seasonal influenza parameterisation), influenza-like parameters with *R*_0_ = 3.0 (pandemic influenza parameterisation). **(Bottom)** SARS-CoV-2-like parameters with *R*_0_ = 3.0.

Estimates for the other parameters were taken from studies of influenza A virus strains [25, 49, 60] and estimates for wild-type SARS-CoV-2 [50, 61]. Comparing between the pandemic influenza and SARS-CoV-2 parameter sets, the notable differences were SARS-CoV-2 having a higher ratio between mild and severe transmission rates (four for SARS-CoV-2, two for pandemic influenza), a longer incubation period (five days for SARS-CoV-2 versus two days for pandemic influenza) and a longer duration of infection for both mild and severe cases (full details in Table 1).

#### (d) Incorporation of symptom propagation into the model framework

We encapsulated symptom severity and symptom propagation into the model framework through two key parameters: *α* - the dependence on the symptom severity of the infector; *ν* - the baseline probability of the pathogen causing severe disease in the absence of propagation effects. The parameter *ν* is aligned with the idea of ‘virulence’ in that it is a measure of the intrinsic severity of the pathogen. When *α* = 0, the symptom severity of the infected individual has no dependence on the infector’s symptom severity; instead, the symptom severity of the infected individual depends entirely on *ν* - this corresponds to the typical assumption applied to compartmental infectious disease models. When *α* = 1, the symptom severity of an infected individual is wholly dependent on that of their infector, meaning that symptom severity is always passed on with infection; in the case of no interventions, this parameterisation is akin to a two-strain model, where one strain causes mild infections only and the other strain causes severe infections only. When *α ∈* (0, 1), the symptom severity of the infectee has a partial dependence on the infector’s symptom severity and a partial dependence on *ν*.

Overall, our model of symptom propagation means that an infected individual, with probability *α*, copies the symptom severity of their infector, whilst with probability 1 *− α*, their symptom severity is assigned randomly according to the underlying probability of having severe disease, *ν* (as depicted in Fig. 1).

#### (e) Baseline model equations (without interventions)

The rate of change for each disease state was governed by the system of differential equations shown in Eq. (1), with parameters as described in Table 1 and Fig. 1. This system of ODEs captures the different levels of disease severity, the dependence of the infectee’s symptom severity on the infector’s symptom severity (through the *α* parameter) and a baseline probability of an infected case having severe disease (through the *ν* parameter).

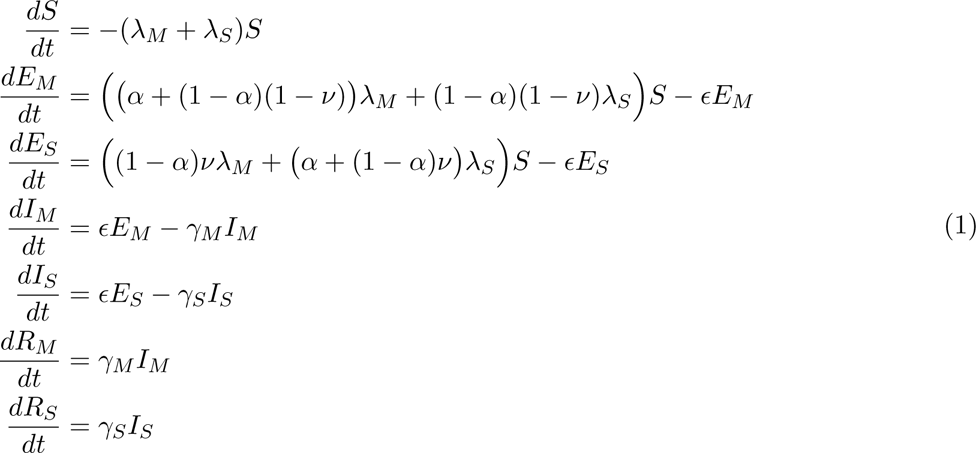

where the force of infection from mild cases, *λ_M_*, and severe cases, *λ_S_*, respectively, were given by:

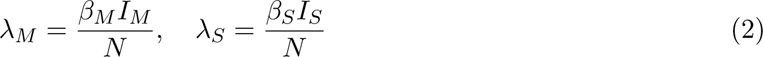

where N is the population size that is assumed to be constant.

#### (f) Exploring the effect of symptom propagation on epidemiological dynamics in the absence of interventions: Simulation overview

In all our model simulations described here and throughout the manuscript, we considered an outbreak arising within a population comparable in size to that of the UK (*N* = 67 million). All the scenarios began with one infectious individual. We assumed that this individual would have severe symptoms with probability given by the baseline probability of severe disease (*ν*). As the model used was fully deterministic, we chose to simulate this effect by splitting the single infectious individual between the two symptom severity classes so that *I_S_* was initialised to contain *ν* people and *I_M_* contained 1 *– ν* people. The remainder of the population were initially susceptible (in the *S* class). The simulations were run until there was less than one individual combined across all the infected classes (*E_M_, E_S_, I_M_* and *I_S_*).

All code was produced using Matlab R2022a and is available at https://github.com/pasplin/ symptom-propagation-mathematical-modelling.

To explore the effect of symptom propagation on the epidemiological dynamics of our three exemplar pathogens, for set values of *α* (0 through 1 in 0.1 increments) and a fixed value of *ν* (0.2), we calculated the following quantities: the final outbreak size (the number of recovered individuals at the end of the simulation), the peak prevalence (the maximum number of individuals infected at any one time) and the outbreak duration (the number of days until cumulatively there was less than one infected individual across all of the infected classes).

We also inspected the dependence of the proportion of infections that were severe with respect to changes in *α* and *ν* for the three exemplar pathogens. We tested combinations of *α*-*ν* values with an increment of 0.05 for each parameter.

Across all simulations, for each value of *α* (and *ν*), *β* was chosen to generate a given *R*_0_ of the required value for that disease parameterisation (further details in Section S1).

### 2.2 Modelling interventions

To investigate how the strength of symptom propagation could impact epidemiological and health economic assessments of infectious respiratory disease intervention strategies, we considered the roll-out of three vaccines with different mechanistic actions on the infectious disease dynamics: (a) a symptom-attenuating vaccine; (b) an infection-blocking vaccine (that did not impact the severity of any breakthrough infections); (c) an infection-blocking vaccine that only admitted mild breakthrough infections. Our vaccines having differing modes of action was motivated by contemporary studies on effectiveness of SARS-CoV-2 vaccines that have estimated effectiveness with respect to infection, symptoms, hospitalisation and mortality [14–16].

We assumed a proportion of the population, given by the vaccine uptake, *u*, were vaccinated before the start of the outbreak. In model terms, we moved a proportion, *u*, of those who would initially be in the susceptible (*S*) class to the vaccinated class, *V*.

We assumed that all three vaccines were imperfect (below 100% efficacy, with details on the assumed vaccine efficacies stated in the upcoming simulation overview subsection) and had a “leaky” action, meaning the susceptibility of all those who were vaccinated was modulated by the vaccine efficacy-in comparison, an “all-or-nothing” action would result in some of those who were vaccinated having full protection and the rest remaining fully susceptible. To minimise complexity, in each simulation, we only studied sole intervention use.

We highlight that the proportion of cases that were severe had a strong dependence on *α*, meaning *α* would have a notable impact on the observed effectiveness of vaccination strategies, even when we would intuitively expect symptom propagation to have no effect. Consequently, we sought to separate the impact of symptom propagation on vaccination strategies from confounding epidemiological factors that would result in an increase in severe cases. Therefore, we explored the effectiveness of the above-described three vaccines for two values of *α*, 0.2 and 0.8, where for each value of *α* we chose the appropriate value of *ν* to fix the proportion of cases (in the absence of interventions) that were severe at 80%. This value was chosen to allow a large value of *α* to be considered, as in this case, the proportion of cases that were severe was high regardless of the value of *ν*. Conceptually, we may consider that the proportion of severe cases is ‘known’ from epidemiological data, and we are fitting parameters to match this data.

We next provide an overview of the modifications to the model equations for each of the three vaccines (depicted in Fig. 2). For each vaccine, we show the modified equations for *E_M_* and *E_S_* states (with changes denoted by red text), with the equations for all other states being unchanged from those in Eq. (1). We also introduce a *V* class, corresponding to susceptible individuals who are vaccinated.

**Figure 2:**
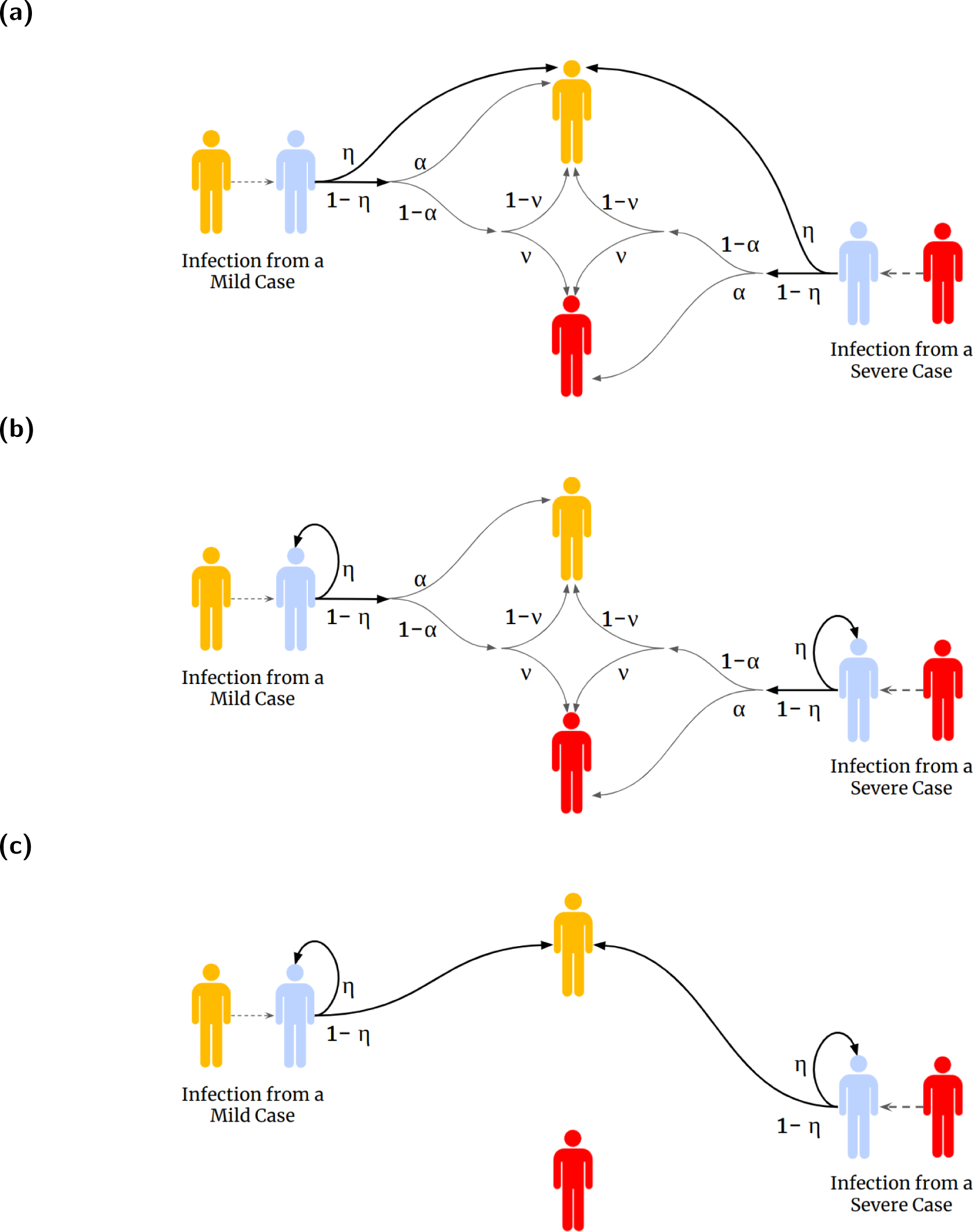
Schematics depicting the mechanistic actions of each vaccine. Across all panels, yellow shaded individuals correspond to infectious cases with mild symptoms, red shaded individuals correspond to infectious cases with severe symptoms, blue shaded individuals correspond to those who are vaccinated. The values on the arrows show the corresponding probability. The three vaccines displayed are: **(a)** symptom-attenuating vaccine - an infected individual who is vaccinated had a probability *η* of having mild disease, and a probability 1*−η* of their symptom severity being determined as usual; **(b)** an infection-blocking vaccine - a vaccinated individual had a probability *η* of their infection being prevented, and a probability 1 *− η* of being infected and their symptom severity being determined as usual; and **(c)** an infection-blocking vaccine that only admits mild breakthrough infections - a vaccinated individual had a probability *η* of their infection being prevented, and a probability 1 *− η* of being infected but only with mild disease.

#### (a) Symptom-attenuating vaccine

First, we considered a vaccine with a symptom-attenuating effect. Vaccinated individuals have probability *η* of having mild disease, and probability 1 *− η* of their symptom severity being determined as usual (Fig. 2(a)).

Updates to the *V* class, corresponding to susceptible individuals who are vaccinated, and the exposed classes (*E_S_* and *E_M_*, with changes from the original model without vaccination - Eq. (1) - shown in red font) followed these ODEs:

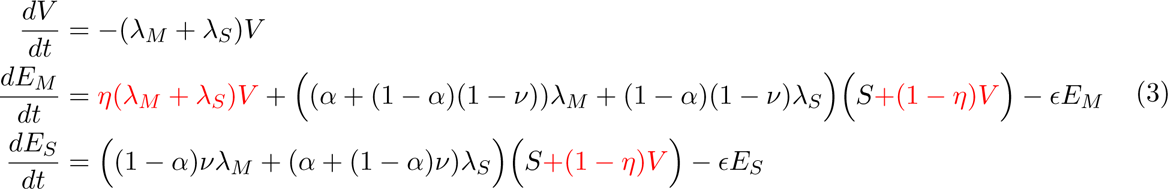

#### (b) Infection-blocking vaccine

Next, we considered a vaccine with an infection-blocking effect. Vaccinated individuals are not infected when exposed with probability *η* (Fig. 2(b)); otherwise infection and symptoms proceed as before. The revised system equations were:

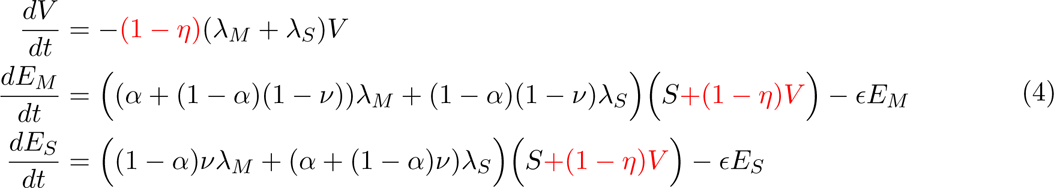

#### (c) Infection-blocking vaccine that only admits mild breakthrough infections

Lastly, we modelled a vaccine with a combined infection-blocking and symptom-blocking effect. The action of this vaccine meant those who were protected were not infected when exposed with probability *η*. Furthermore, all vaccinated individuals only develop mild disease, regardless of the efficacy of the vaccine in blocking infection (Fig. 2(c)).

The revised system equations were:

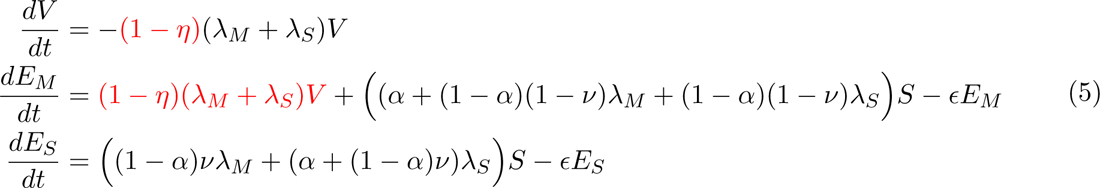

Additionally, we modelled another infection-blocking vaccine that had an alternative manner in which breakthrough infections could arise, namely breakthrough infections were only possible when the infector was a severe case. We found no difference in results between this vaccine type and the infection-blocking vaccine. We provide the model schematic, equations and summary of findings in Section S4.

#### (d) Exploring the effect of symptom propagation on epidemiological dynamics in the presence of vaccination: Simulation overview

Across our main vaccination analyses, the vaccine efficacy was fixed at 70% based on estimates of COVID-19 vaccine efficacy [14, 15]. This vaccine efficacy level was also a reasonable selection for influenza vaccines since the vaccine efficacy during the 2009/10 season was estimated to be 72% vs the pandemic H1N1 strain [62], and other studies have estimated influenza vaccine efficacy to be between 56-78% [63] and between 26-73% [64]. To assess sensitivity to vaccine efficacy, as supplementary analyses, we also considered vaccine efficacies of 50% and 90%, where we found qualitatively similar results (see Section S7, Figs. S16 to S23).

For the three vaccine types, two vaccine uptake levels (50% and 90%) and three sets of disease parameters, we calculated the proportion of the population in each recovered compartment (having had mild or severe symptoms) at the end of the outbreak for two values of *α* (0.2 and 0.8), with *ν* chosen to fix the proportion of cases that were severe without intervention at 80%.

For a range of values of *α* and *ν* (0 through 1 in 0.02 increments), we also calculated the difference in the number of individuals severely infected when an infection-blocking vaccine was used compared to a symptom-attenuating vaccine.

### 2.3 Health economic modelling

Often there are many potential intervention strategies that can be used to limit the spread of the disease. Since public health decision makers have a finite budget, the cost of the intervention is an important factor to consider alongside the resulting epidemiological outcomes. Thus, measures for both the benefit to public health and the costs associated with the intervention and treatment warrant consideration.

#### Measures of health quality and model parameterisation

The chosen measure to quantify disease burden was quality-adjusted life years (QALYs), which consider both the quality and quantity of years lived [65]. We assumed there were no QALY losses associated with mild cases. The magnitude of QALY loss from a severe case depended on whether the case was hospitalised and whether it wasfatal; a proportion of severe cases was assumed to lead to hospitalisation and fatalities, as dictated by the pathogen-specific hospitalisation rate and death rate (parameter details in Table 2). Hospitalisations and fatalities also had an associated monetary cost, where once again values differed for influenza and SARS-CoV-2 (Table 2). Further details of the health economic model parameters are provided in Section S3.

**Table 2:** Description of parameters used in the health economic modelling. **(Top)** Values applied to both the seasonal influenza and pandemic influenza disease parameterisations. **(Bottom)** Values applied to the SARS-CoV-2 disease parameterisation.

| Description | Influenza value | Source |
| --- | --- | --- |
| Probability of hospitalisation (given severe disease) | 0.01 | Hill *et al.* [41] |
| Probability of death (given severe disease) | 0.001 | Hill *et al.* [41] |
| QALY loss from a mild case | 0 QALYs | Assumption |
| QALY loss from a severe, non-hospitalised case | 0.008 QALYs | Hill *et al.* [41] |
| QALY loss from a non-fatal hospitalised case | 0.018 QALYs | Hill *et al.* [41] |
| QALY loss from a fatal hospitalised case | 37.5 QALYs | Hollmann *et al.* [66] |
| Total cost of a non-fatal hospitalised case | £1,300 | Hill *et al.* [41] |
| Total cost of a fatal hospitalised case | £2,600 | Hill *et al.* [41] |
| Willingness to pay threshold per QALY | £20,000 | NICE [67] |

| Description | SARS-CoV-2 value | Source |
| --- | --- | --- |
| Probability of hospitalisation (given severe disease) | 0.065 | Moran *et al.* [68] |
| Probability of death (given severe disease) | 0.02 | Moran *et al.* [68] |
| QALY loss from a mild case | 0 QALYs | Assumption |
| QALY loss from a severe, non-hospitalised case | 0.0035 QALYs | Moran *et al.* [68] |
| QALY loss from a non-fatal hospitalised case | 0.0059 QALYs | Moran *et al.* [68] |
| QALY loss from a fatal hospitalised case | 11.29 QALYs | Moran *et al.* [68] |
| Total cost of a non-fatal hospitalised case | £2,600* | Vekaria *et al.* [69] |
| Total cost of a fatal hospitalised case | £5,200* | Vekaria *et al.* [69] |
| Willingness to pay threshold per QALY | £20,000 | NICE [67] |

\* Hospitalisation costs for SARS-CoV-2 are double those of influenza, based on the average hospital stay being twice as long for SARS-CoV-2 versus influenza - further details available in Section S3.

We deemed an intervention to be cost-effective if the overall cost of implementing the intervention was less than or equal to the value of QALYs gained from doing so. In particular, we computed threshold unit intervention costs, the monetary cost of an intervention unit that would result in intervention costs equalling the monetary value of QALYs gained. In this case, the threshold unit intervention cost refers to the threshold cost per vaccine dose.

Determining whether an intervention is cost-effective requires setting a willingness to pay (WTP) threshold per QALY - the amount one is willing to pay to gain one QALY. We used a default WTP per QALY of £20,000, reflecting the typical criteria used in England that alternative intervention strategies need to satisfy to be judged as cost-effective [70]. Equivalently,

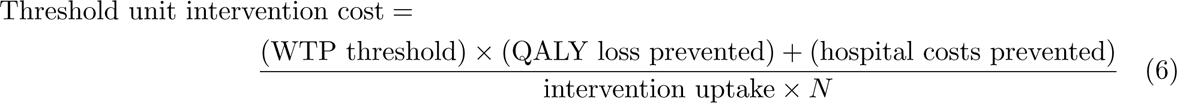

In all simulations, the threshold unit intervention cost was normalised with respect to the highest absolute threshold unit intervention cost attained for that disease parameterisation across the range of tested vaccine uptake values.

#### Exploring the effect of symptom propagation on health economic outcomes: Simulation overview

For the three vaccine types, two vaccine uptake levels (50% and 90%) and three disease parameterisations, we conducted comparisons of the relative threshold unit intervention cost between two values of *α* (0.2 and 0.8) with *ν* chosen to fix the proportion of cases that were severe without intervention at 80%. We then explored how the threshold unit intervention costs varied with the vaccine uptake, looking at uptake levels between 0% and 100% in 1% increments.

For the results presented in the main text, we applied a 3.5% discounting rate to both QALY losses and monetary costs, as recommended [71]. For sensitivity purposes, we also tested having no discounting, with results given in Section S6 (Figs. S14 and S15).

## 3 Results

### 3.1 Symptom propagation affects epidemiological dynamics

Initially, we explored the effect of varying the strength of symptom propagation on epidemiological outcomes for a fixed baseline probability of severe disease, *ν* = 0.2. Since, for each set of parameters, the value of *R*_0_ was fixed, the resultant outbreak size remained mostly constant as we varied the dependence on infector symptom severity, *α* (Figs. 3(a) to 3(c)), although there was a slight reduction in final outbreak size as *α* approached 0.5 due to numerical inaccuracies (Fig. S6). As these differences in case numbers were proportionally small, we assumed the final size to be fixed throughout the remainder of the analysis.

**Figure 3:**
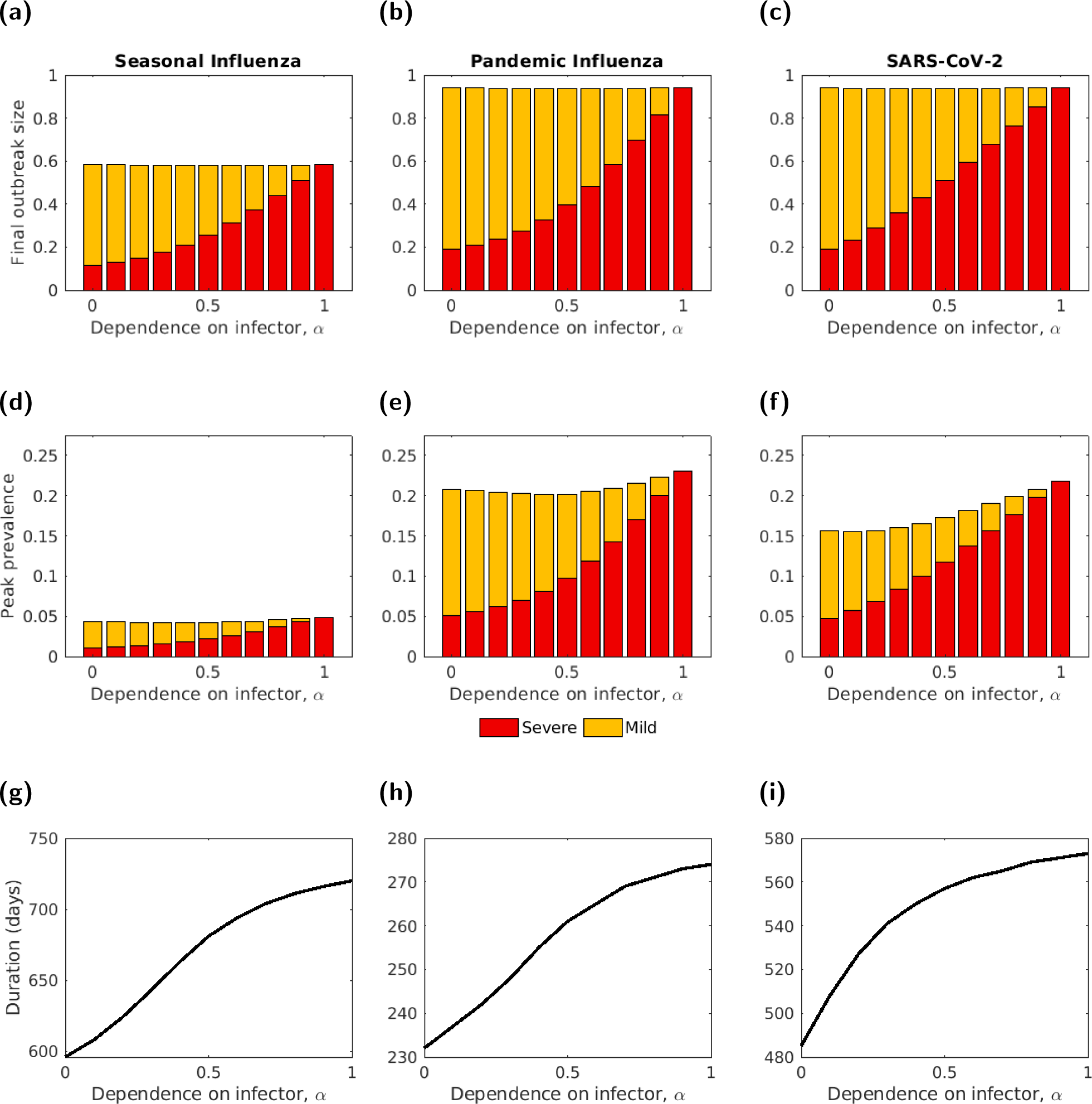
The final outbreak size, peak prevalence and outbreak duration, by severity, for three disease parameterisations, plotted for different symptom propagation strengths, *α*. (a-c) Final outbreak size by severity. **(d-f)** Peak prevalence by severity. The intensity of the shading denotes the symptom severity class, with severe cases in red and mild cases in yellow. **(g-i)** Outbreak duration (note the different y-axis scales). In all panels, *ν* = 0.2. The three disease parameterisations used were: **(a,d,g)** Seasonal influenza; **(b,e,h)** Pandemic influenza; **(c,f,i)** SARS-CoV-2, with parameters as given in Table 1.

Considering the stratification of cases by severity (mild or severe), the proportion of total cases that were severe monotonically increased with *α*. As expected, when *α* = 0, the proportion of cases that were severe was equal to the baseline probability of severe disease, *ν* = 0.2. This proportion increased to effectively all cases being severe when *α* = 1 (Figs. 3(a) to 3(c)). Similarly, the proportion of cases that were severe at the peak of the outbreak increased with *α*, with this proportion aligning with the proportion severe overall (Figs. 3(d) to 3(f)). The outbreak duration also increased with *α* (Figs. 3(d) to 3(i)) due to individuals with severe disease having a longer infectious duration. Comparing *α* = 1 with *α* = 0 across all three disease scenarios, we observed an increase of approximately 20% in the outbreak duration.

Overall, the qualitative patterns of the impact of symptom propagation differed relatively little between the three sets of disease parameters. Unsurprisingly, seasonal influenza had a much lower final outbreak size and peak prevalence than the other two disease parameterisations (Figs. 3(a) and 3(d), due to its lower assumed *R*_0_ value). More interestingly, SARS-CoV-2 had a lower peak prevalence than pandemic influenza, especially for low values of *α* (Figs. 3(e) and 3(f)), with longer generation times for SARS-CoV-2 dominating the effects of a higher *R*_0_ value. SARS-CoV-2, compared to the two influenza disease parameterisations, showed a slightly higher proportion of severe cases for intermediate values of *α*. Furthermore, the proportion of cases that were severe (both overall and at peak) for SARS-CoV-2 exhibited a roughly linear increase as *α* increased from 0 to 1, whereas for influenza these metrics increased sub-linearly for *α* between 0 and 0.5, and approximately linearly for increasing *α* between 0.5 and 1 (Figs. 3(d) to 3(f)).

Next, we considered how the value of the baseline probability of severe disease, *ν*, impacted epidemiological outcomes. As expected, the proportion of cases that were severe increased with *ν* (Fig. 4). For values of *α* close to 0, the value of *ν* mostly determined the proportion of cases that were severe. In contrast, when *α* was close to 1, the proportion of cases that were severe remained high, independent of the value of *ν*. The relationship between *α, ν* and the proportion of cases that were severe was consistent across parameter sets.

**Figure 4:**
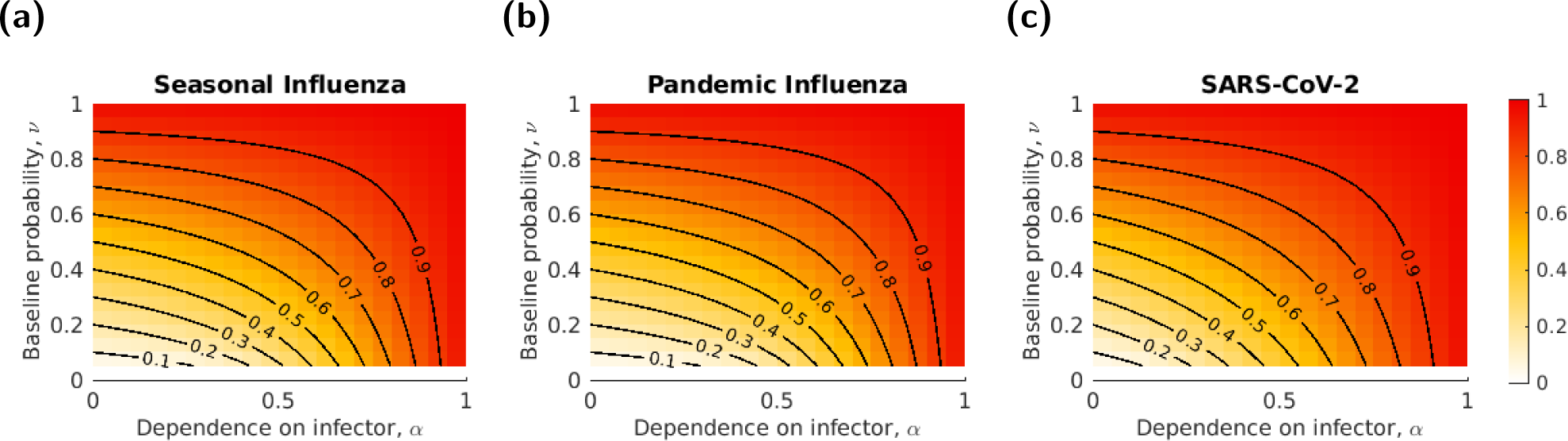
The proportion of infections that were severe against changes in *α* and *ν* for three parameter sets. The shading shows the proportion of infections that were severe, with darker shading corresponding to a higher proportion being severe. Black lines correspond to the contours taking values from 0.1 to 0.9, at increments of 0.1. The three disease parameterisations used were: **(a)** Seasonal influenza; **(b)** Pandemic influenza; **(c)** SARS-CoV-2. The parameters were as given in Table 1.

### 3.2 Symptom propagation increases the effectiveness of interventions that impact symptom severity

In this section, we explored three types of intervention, corresponding to three plausible vaccination scenarios: a symptom-attenuating vaccine (SA), an infection-blocking vaccine (IB) and an infection-blocking vaccine that only admits mild breakthrough infections (IB MB). We considered two vaccine uptake rates (50% and 90%) and two values of *α* (0.2 and 0.8). In order to highlight the differences between vaccine types, the value of *ν* was chosen (as a function of a given value of *α*) to fix the proportion of cases that were severe equal to 0.8. We additionally produced an analogous set of results with *ν* fixed equal to 0.2 for comparison (Figs. S12 and S13), noting there would be an inherent relative reduction in potential impact of SA interventions for that scenario.

For all parameter sets and both uptake and *α* values, the IB MB vaccine type was, unsurprisingly, the most effective at reducing both total and severe cases (Fig. 5). Similarly, a solely infection-blocking vaccine was always more effective at reducing total cases than a symptom-attenuating vaccine. In the case of seasonal influenza, both IB and IB MB were sufficient to fully suppress the outbreak (*<* 0.01% of the population was infected), even at 50% uptake, whereas 90% uptake was required for the SA vaccine to suppress the outbreak. However, in many cases, the symptom-attenuating vaccine was more effective at reducing severe cases than the solely infection-blocking vaccine. This effect was seen for both *α* values for pandemic influenza or SARS-CoV-2 and 50% uptake (Figs. 5(b) and 5(c)). For these two disease parameterisations, when the uptake was instead 90%, whether the SA or IB vaccine was more effective at reducing severe cases depended on the value of *α*, with the higher value of *α* = 0.8 resulting in the SA vaccine being more effective (Figs. 5(e) and 5(f)).

**Figure 5:**
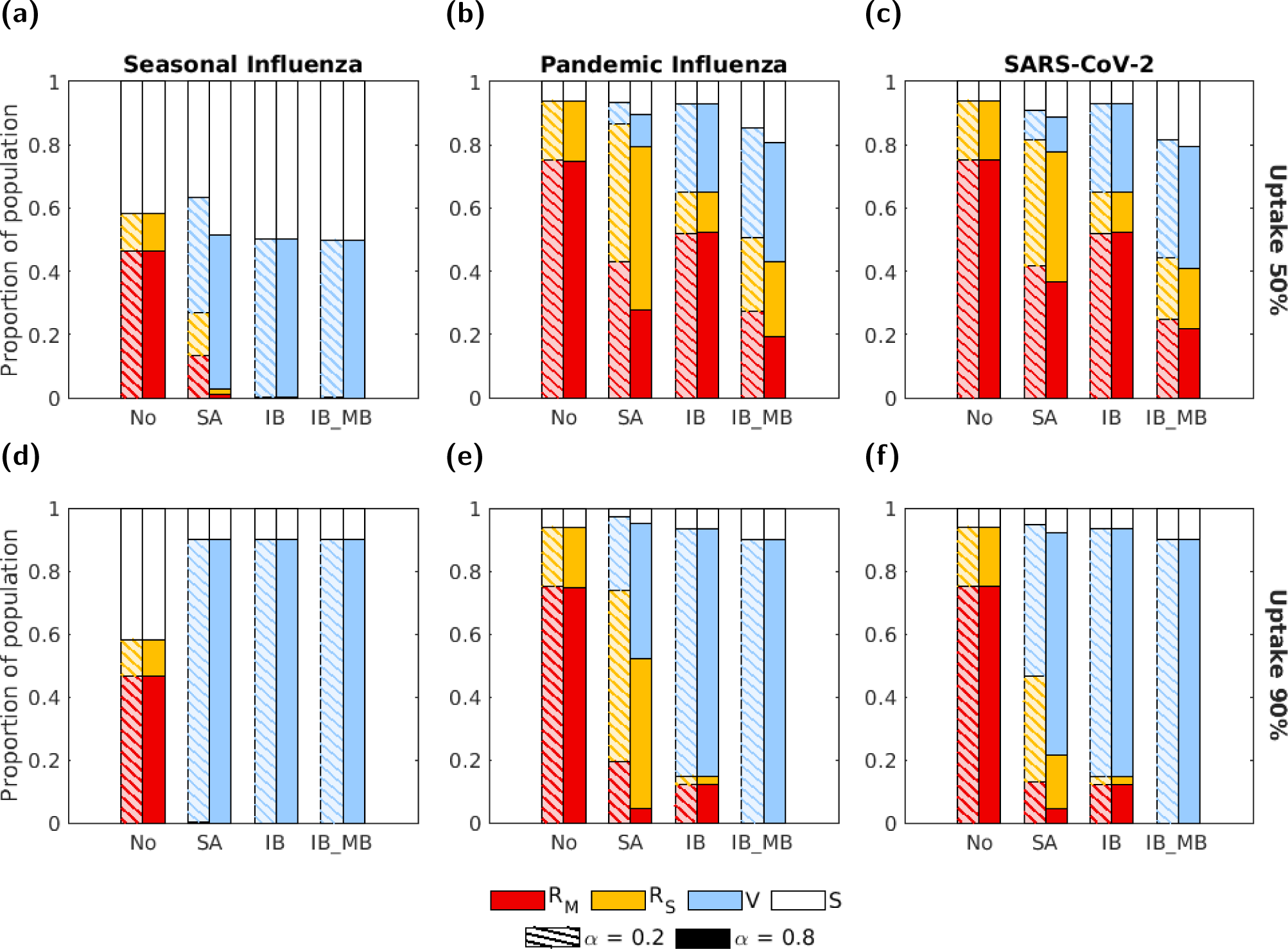
The proportion of the population in each disease state at the end of the outbreak for the four intervention scenarios, two vaccine uptake levels and three disease parameterisations. The four groups of bars correspond to four intervention scenarios: no intervention (No), a symptom-attenuating vaccine (SA), an infection-blocking vaccine (IB) and an infection-blocking vaccine which only admits mild breakthrough infections (IB MB). The two bars in each group correspond to two different strengths of symptom propagation: *α* = 0.2 (left bar with hatched lines) and *α* = 0.8 (right bar with solid fill). Bar shading corresponds to the disease status: red - recovered from severe infection (*R_S_*); yellow - recovered from mild infection (*R_M_*); blue - susceptible and vaccinated (*V*); white - susceptible and not vaccinated (*S*). The two rows correspond to two vaccine uptake levels: **(a-c)** 50%; **(d-f)** 90%. Columns correspond to different disease parameterisations: **(a,d)** seasonal influenza; **(b,e)** pandemic influenza; **(c,f)** SARS-CoV-2. We fixed the vaccine efficacy at 70% and all other parameters as given in Table 1, with *ν* chosen to fix the proportion of cases that were severe equal to 0.8.

Across all scenarios, the IB vaccine resulted in the same epidemiological outcomes for both values of *α*. For the other two vaccine types (SA and IB MB), effectiveness was always higher for the higher *α* value, with the exception of scenarios in which the outbreak was suppressed for both *α* values (*<* 0.01% of the population was infected). Results were similar when considering peak prevalence (Fig. S10). For all intervention scenarios (including no intervention), the duration was higher for *α* = 0.8 compared to *α* = 0.2 for those instances where the outbreak was not effectively prevented (*>*0.01% of the population was infected, Fig. S11). Inspection of temporal profiles of the outbreaks revealed that, in all intervention scenarios, the peak prevalence occurred later for *α* = 0.8 than for *α* = 0.2, even when no intervention was used (Figs. S7 to S9). For the no intervention and IB vaccine scenarios, this delay was a noticeable change in the temporal dynamics between the two *α* values; otherwise, the temporal dynamics largely exhibited similar qualitative behaviour.

We then explored the difference in the number of severe cases prevented by an IB vaccine and an SA vaccine for the three disease parameterisations and two vaccination uptake levels, under a fixed vaccine efficacy (70%) (Fig. 6). We found that the IB vaccine was always more effective at preventing severe cases in the case of seasonal influenza (Figs. 6(a) and 6(d)). For the other two parameter sets, the results were qualitatively similar. For a lower vaccine uptake (50%), the SA vaccine was more effective at reducing severe cases for almost all values of *α* and *ν* (red shaded cells of Figs. 6(b) and 6(c)). In contrast, when the vaccine uptake was high (90%), which vaccine type was most effective at reducing severe cases depended on the values of *α* and *ν*, with the SA vaccine being more effective for larger values of *α* and *ν* (Figs. 6(e) and 6(f)).

**Figure 6:**
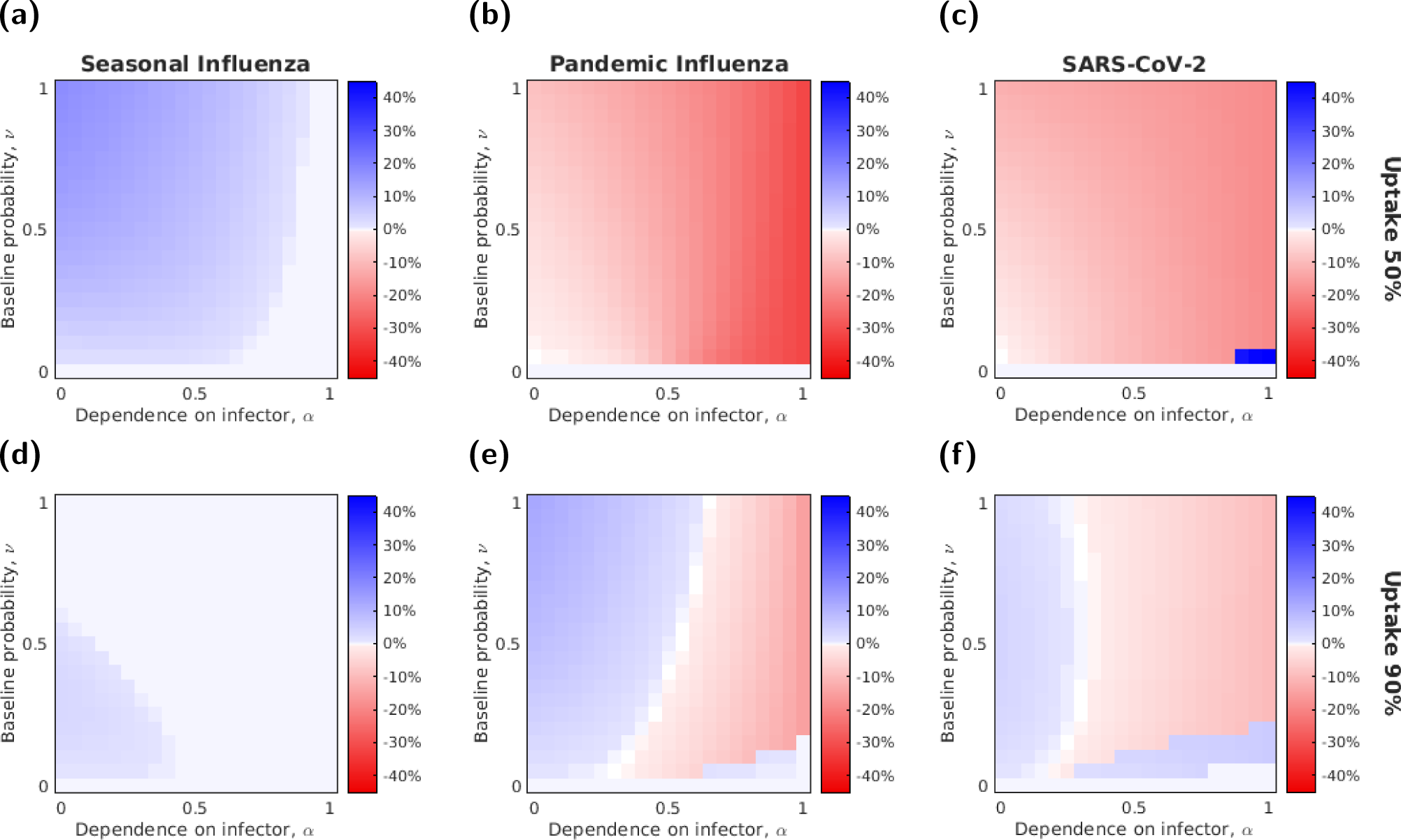
The relative effectiveness in reducing severe cases of a symptom-attenuating and infection-blocking vaccine as a function of *α* and *ν*, given a fixed efficacy (70%). Rows correspond to the two vaccine uptake levels **(a-c)** 50%; **(d-f)** 90%, and columns to the different disease parameterisations (**(a,d)** seasonal influenza; **(b,e)** pandemic influenza; **(c,f)** SARS-CoV-2). Pixel shading denotes (for given combinations of *α*-*ν* values) the difference in the proportion of the population severely infected between vaccine types, such that blue regions show parameter combinations where the infection-blocking vaccine was more effective at reducing the number of severe cases and red regions show those where the symptom-attenuating vaccine was more effective.

### 3.3 Symptom propagation can affect health economic outcomes

Switching attention to how the strength of symptom severity propagation can impact health economic assessments, the differences in the threshold unit intervention cost for the two values of *α* aligned with our previously presented results (Fig. 7).

**Figure 7:**
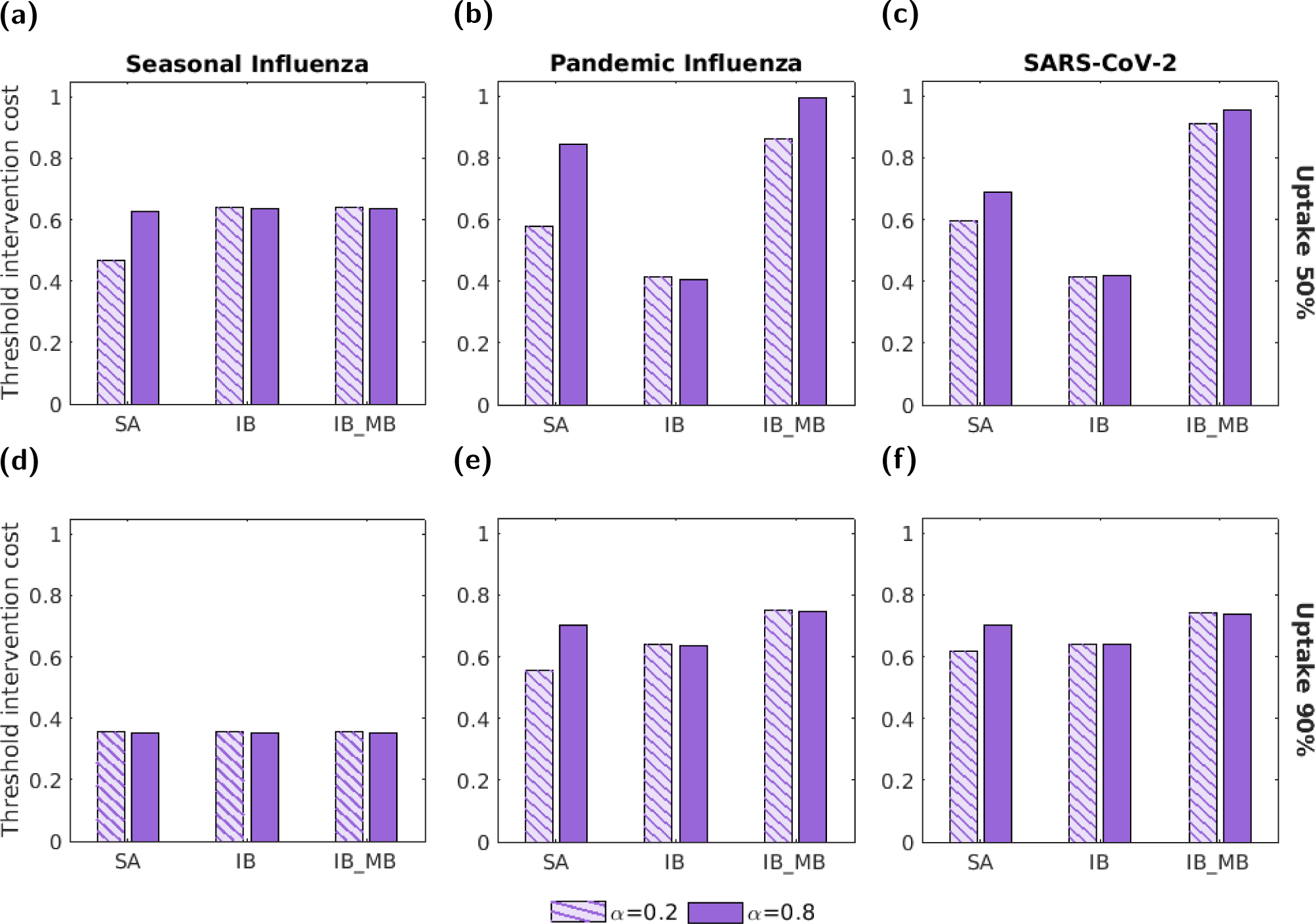
Threshold unit intervention cost for the three types of vaccine, two vaccine uptake levels and the three disease parameterisations. In all panels, we normalised threshold intervention costs by the highest absolute threshold unit intervention cost obtained across the range of tested vaccine uptake values. The three groups of bars correspond to: a symptom-attenuating vaccine (SA), an infection-blocking vaccine (IB) and an infection-blocking vaccine which only admits mild breakthrough infections (IB MB). The two bars in each group correspond to symptom propagation strengths of *α* = 0.2 (left bar, hatched lines) and *α* = 0.8 (right bar, solid fill). The two rows correspond to two vaccine uptake levels: **(a-c)** 50%; **(d-f)** 90%. Columns correspond to differing disease parameterisations: **(a,d)** seasonal influenza; **(b,e)** pandemic influenza; **(c,f)** SARS-CoV-2. Vaccine efficacy was fixed at 70% and all other parameters were as given in Table 1, with *ν* chosen to fix the proportion of cases that were severe equal to 0.8.

When comparing between vaccine types, we found that the IB MB vaccine always had the highest threshold unit intervention cost of the three vaccine types considered. For the pandemic influenza and SARS-CoV-2 parameterisations, SA always had a higher threshold unit intervention cost (and was, therefore, more cost-effective) than IB when *α* = 0.8 (Figs. 7(b), 7(c), 7(e) and 7(f)). When *α* = 0.2, SA was only more cost-effective than IB when uptake was low (50%) (Figs. 7(e) and 7(f)).

When comparing between the two *α* values, there was the least variation observed for the IB vaccine, where the threshold unit intervention cost was roughly the same for the two *α* values in all uptake and disease parameterisation scenarios. This result aligns with our previous finding that *α* had no effect on epidemiological outcomes when an IB vaccine was used (Fig. 5). For the other two vaccine types, SA and IB MB, the threshold unit intervention cost was higher when *α* = 0.8, unless the outbreak was effectively contained (*<* 0.01% of the population was infected) for both *α* values, in which case the threshold unit intervention costs were the same. In all of these scenarios, the relative difference in threshold unit intervention cost was higher for the influenza parameter sets (16%-45% increase, Figs. 7(a), 7(b), 7(d) and 7(e)) than for the SARS-CoV-2 parameter set (Figs. 7(c) and 7(f), 5%-16% increase). The relative difference in outcomes between *α* values was also generally higher for the SA vaccine than for the IB MB vaccine (14%-45% increase for SA vs 5%-16% for IB MB). In all cases, as anticipated the observed differences in threshold unit intervention cost reflect the differences in epidemiological outcomes (Fig. 5).

We then explored how vaccine cost-effectiveness varied for a range of *α* and *ν* values (Figs. S24 to S26). We found that the threshold unit intervention cost increased with both *α* and *ν* irrespective of the action of the vaccine as a consequence of higher values of *α* and *ν* causing a larger proportion of cases to be severe.

We then applied greater scrutiny to how the threshold unit intervention cost depended on the level of uptake for the three vaccine types and three disease parameterisations (Fig. 8). For both seasonal influenza (Figs. 8(a) to 8(c)) and pandemic influenza (Figs. 8(d) to 8(f)), the difference in threshold unit intervention cost (obtained for *α* = 0.2 and *α* = 0.8) decreased as vaccine uptake increased. This was particularly noticeable for the SA vaccine, where the difference decreased somewhat linearly until the point where the values converged. For the SARS-CoV-2 parameterisation, the difference in threshold unit intervention cost between the two *α* values remained relatively constant as the uptake increased, up until the point where the two values converged (Figs. 8(g) to 8(i)).

**Figure 8:**
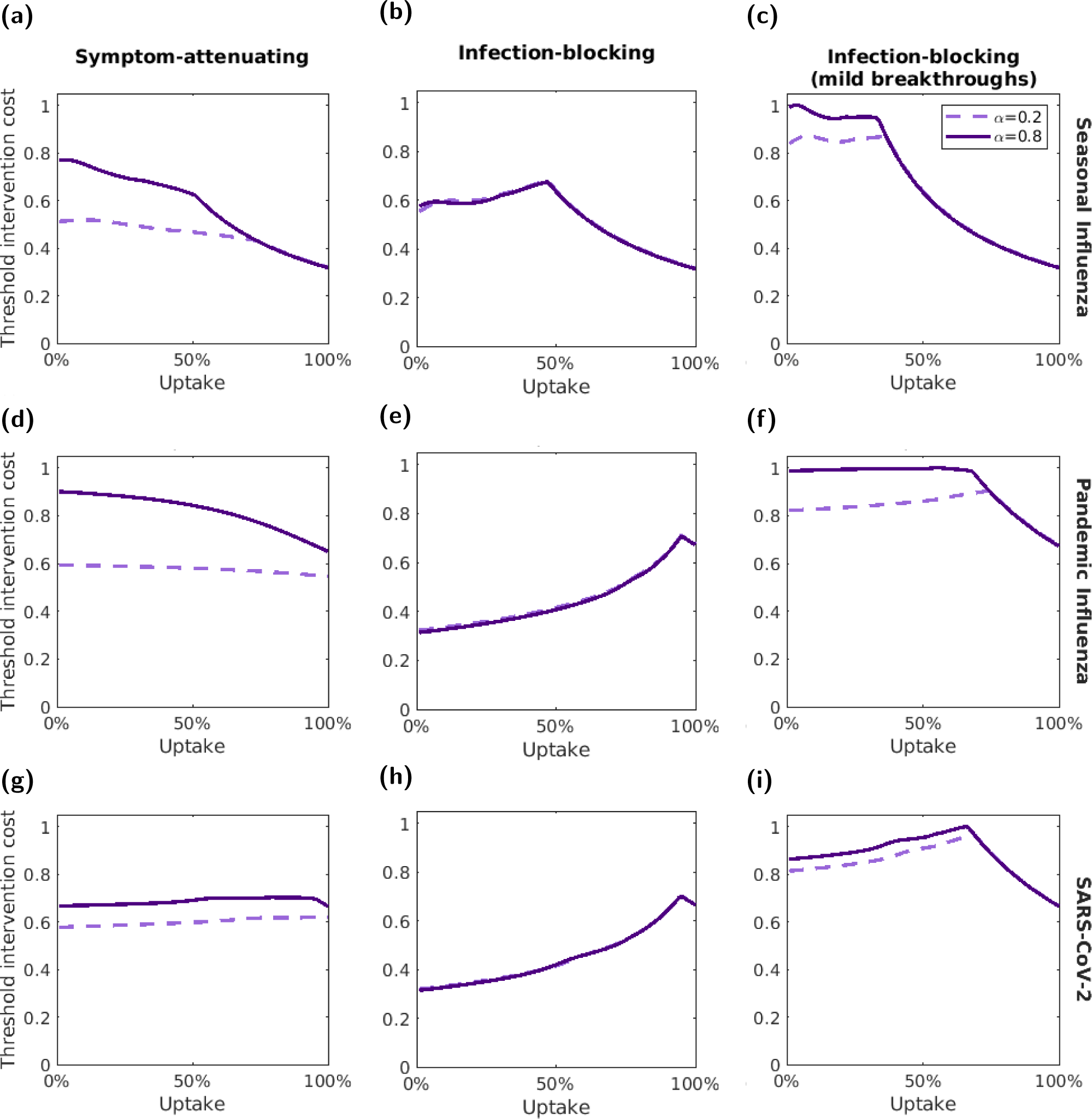
Variation of the threshold unit intervention cost with vaccine uptake. We normalised threshold unit intervention costs for each disease parameterisation; the normalisation constant was the highest absolute threshold unit intervention cost attained for the respective disease parameterisation across the range of tested uptake values. The three rows correspond to the three different disease parameterisations: **(a-c)** seasonal influenza; **(d-f)** pandemic influenza; **(g-i)** SARS-CoV-2. The three columns correspond to: **(a,d,g)** a symptom-attenuating vaccine (SA), **(b,e,h)** an infection-blocking vaccine (IB) and **(c,f,i)** an infection-blocking vaccine that only admits mild breakthrough infections (IB MB). The two lines correspond to two symptom propagation strengths; the dashed, light purple line corresponds to *α* = 0.2 and the solid, dark purple line corresponds to *α* = 0.8. We fixed the vaccine efficacy at 70% and all other parameters values were as given in Table 1, with *ν* chosen to fix the proportion of cases that were severe equal to 0.8.

As previously, we observed different effects of *α* for the different types of vaccine. For the IB vaccine, we found that the threshold unit intervention cost was equal for the two values of *α* at all vaccine uptake levels (Figs. 8(b), 8(e) and 8(h)). For the SA vaccine (Figs. 8(a), 8(d) and 8(g)) and IB MB vaccine (Figs. 8(c), 8(f) and 8(i)), the threshold unit intervention cost remained higher for *α* = 0.8 as the uptake increased, up until the uptake level at which the outbreak was suppressed for both *α* values. After this point, for both *α* values, the threshold unit intervention costs were equal and decreased monotonically with vaccine uptake.

Across all uptake and disease parameterisation scenarios, the most cost-effective uptake (i.e. the uptake that maximised the threshold unit intervention cost) varied minimally with *α*. There was a consistent general relationship between the threshold unit intervention cost and the vaccine uptake between the two *α* values.

We further explored the most cost-effective uptake value for a range of *α* and *ν* values and found that the most cost-effective level of vaccine uptake (i.e the uptake with the highest threshold unit intervention cost) generally remained constant across *α* and *ν* values (Figs. S27 to S29). Some exceptions to this did arise for the vaccines that were symptom-attenuating and infection-blocking with mild break-through infections. For certain disease parameterisations and efficacy values, we found large variation in the most cost-effective uptake, from close to 0% to nearly 100% in some parameterisations. Between scenarios, we did not observe a simple qualitative pattern of variation, although the regions where uptake close to 0% was most cost-effective tended to have lower values of *α* and *ν*.

Lastly, to give insight into how outcomes could differ if symptom propagation was mistakenly omitted from the modelled infection dynamics, we compared between the threshold unit intervention cost of the most cost-effective uptake for a particular value of *α* and the threshold unit intervention cost for the most cost-effective uptake at *α* = 0 (Figs. S30 to S32). We found that generally the difference in threshold unit intervention costs increased with *α* for both the SA vaccine and the IB MB vaccines.

## 4 Discussion

In this paper, we make three main contributions to the literature. Firstly, we introduce a parsimonious and generalisable mechanistic mathematical framework to model infectious disease transmission that incorporates symptom propagation of different strengths via a single parameter, *α*. Secondly, we demonstrate substantial impacts of symptom propagation on epidemiological outcomes. For parameterisations corresponding to seasonal influenza, pandemic influenza and SARS-CoV-2, we demonstrate that, for a given value of *R*_0_, even for a low baseline probability of severe disease, *ν*, (which we conceptualise as relating to the virulence of the pathogen) the proportion of cases that experience severe disease can approach one as the strength of symptom propagation, *α*, increases. Thirdly, we apply three types of intervention, corresponding to three plausible vaccination scenarios (symptom-attenuating, SA, infection-blocking, IB, and infection-blocking with mild breakthrough infections, IB MB), and demonstrate important impacts of symptom propagation on epidemiological and health economic outcomes. We showed that although the strength of symptom propagation had no effect on epidemiological outcomes for IB interventions, differences were seen for interventions that acted to reduce symptom severity, with the effectiveness of these interventions in reducing the number of severe and total cases increasing with the strength of symptom propagation. The strength of symptom propagation also affected the relative effectiveness of SA and IB interventions in reducing severe cases, with the optimal type of intervention dependent on a combination of uptake and *α*. In the health economic analysis, we found that the strength of symptom propagation had important implications for the cost-effectiveness of SA interventions, and to a lesser extent, IB MB interventions. Thus, we demonstrated how symptom propagation influences both epidemiological and health economic outcomes, which can alter the balance between preferred intervention types.

A cornerstone of our encapsulation of symptom propagation was the parameter *α*, the dependence of the symptoms of an infected individual on the symptoms of their infector. Our general finding was that the proportion of cases that were severe increased with *α*. Although the effect of symptom propagation on the proportion of cases that are severe has not received attention in previous modelling studies, this result aligns with suggestions that symptom propagation, at least through a dose-response relationship, could lead to severe outbreaks and intense epidemics [24]. This result also aligns with the findings of Paulo *et al.* [35], who found that the inclusion of a dose-response relationship in their model led to an increase in the incidence of severe disease and higher mortality. The appreciable effect of *α* on epidemiological outcomes motivates the inclusion of symptom propagation in models of infectious disease transmission, both when simulating an outbreak from a given set of parameters and when estimating parameters from an empirical data set. It also highlights the importance of examining in more detail the basic biological mechanisms of symptom propagation for respiratory pathogens.

We found that the proportion of cases that were severe not only increased with *α*, but also with the baseline probability of severe disease, *ν*. In many scenarios, for a given value of *α*, it was possible to compute the value of *ν* that returned a pre-specified proportion of cases that were severe. However, this was limited to relatively low values of *α*, since for high values of *α* the proportion of cases that were severe was large regardless of the value of *ν*. As a result of this, when the proportion of cases that were severe was fixed, the proportion chosen was relatively high (80%) to allow for the consideration of a value of *α* close to one. We acknowledge that it may be unrealistic for such a high percentage of cases to have severe symptoms, with the proportion of cases that are mild estimated to be 43% for influenza (where mild is defined as subclinical) [72] and 44% for SARS-CoV-2 (where mild is defined as asymptomatic) [73]. It may also be the case that such high values of *α* (i.e. high amounts of symptom propagation) are unrealistic, although we know that for certain diseases, such as plague, the value of *α* is close to one [22]. Instead, this overestimation of the proportion of cases that are severe for high values of *α* may be due to our subjective parameterisation of some of the epidemiological parameters, for example, the relative transmissibility of mild and severe disease. Alternatively, it may be that such strengths of symptom propagation are realistic, but are not observed due to behavioural changes not included in this model, since individuals displaying symptoms have been shown to reduce their contact with others in their community [74]. The inclusion of behavioural responses is an important area of infectious disease modelling identified for dedicated research.

When exploring the effect of symptom propagation on the effectiveness of interventions, we found that interventions affecting symptom severity (i.e. the symptom-attenuating intervention or infection-blocking intervention with mild breakthroughs infections) were consistently more effective at reducing cases and were more cost-effective for a higher value of *α*; exceptions to this were when the intervention suppressed the outbreak for both *α* values under consideration (*α* = 0.2 and *α* = 0.8). In contrast, varying *α* had little or no effect on epidemiological outcomes when the intervention was purely infection-blocking. These results suggest that determining the effect, if any, of an intervention on the symptoms experienced by individuals is critical to understand whether symptom propagation is an important consideration when investigating the effectiveness of the intervention.

Given increasing evidence that many interventions used to prevent the spread of disease also reduce symptom severity, it may be the case that symptom propagation should be considered more often than not. Indeed, vaccinated individuals are more likely to have asymptomatic disease in addition to having a lower risk of infection, as was the case for COVID-19 vaccines [75]. Additionally, it has been indicated that non-pharmaceutical interventions such as social distancing and mask-wearing can reduce the infectious dose for onward transmission, leading to less severe disease in those infected [20, 21, 76].

For disease parameterisations with a higher *R*_0_ (corresponding to pandemic influenza and SARS-CoV-2) and a high intervention uptake, comparisons between intervention types showed that given a strong symptom propagation action, a symptom-attenuating intervention was more effective at reducing severe cases than an infection-blocking intervention, even though total outbreak sizes were larger. This finding suggests dual benefits of SA interventions: reduced numbers of severe cases alongside widespread population immunity. Similar effects have been described previously. For example, a so-called “variolation effect” has been discussed in the context of mask-wearing during the COVID-19 pandemic; some authors hypothesise that masks could act to reduce the inoculum dose leading to reduced disease severity in those infected, and that this effect could have been used to generate widespread immunity before vaccines became available [19, 21].

The intervention type ranked as most effective at reducing severe cases (when comparing between them) had a notable dependence on *α*, *ν* and the disease parameterisation. Accordingly, symptom propagation may be an important factor to consider when choosing between intervention types. A renewed analysis of interventions for the containment, suppression and management of respiratory pathogens of public health concern could result, for example via the examination of interventions that can create large-scale population immunity while minimising the number of severe cases. Such modelling analysis is only viable by us taking a contemporary approach to capturing actions of interventions, rather than treating them as being solely infection blocking. The data arising from SARS-CoV-2 vaccines having a stratification of efficacy for different health episode outcomes (infection, symptomatic, hospitalisation, mortality) [14–16] motivates a similar richness of data collection being undertaken for other pathogens. Seasonal influenza is one such pathogen where additional information would be informative; vaccination effectiveness has historically been assessed using a ‘test-negative’ design, meaning patients with influenza-like illness are tested for influenza, with reported vaccine effectiveness usually relating solely to the prevention of symptomatic infection [77], without further stratification of outcomes.

There are a number of limitations to the work conducted in this paper. First, there was some uncertainty in the parameters used in the epidemiological model. We sourced parameters from the literature, where disparate estimates were reported. Certain parameters, such as the relative transmissibility of mild and severe disease, were difficult to measure. Whilst acknowledging that our results may be sensitive to the parameterisations chosen, to retain our focus on the implications of symptom propagation and interventions that had different modes of action, we took a pragmatic approach of considering a single fixed relative transmissibility scaling for each pathogen informed by the available literature. These relative transmissibility scalings were also dissimilar, two for influenza and four for SARS-CoV-2 giving us more breadth in our coverage of disease parameter space. A notable limitation of these choices is that the transmission rates do not account for changes in contact patterns that we would expect to see for those with more severe disease, and hence may be an overestimation of the real values. The inclusion of heterogeneity in contact patterns is highlighted as an area for future work. In future work, it would be valuable to include heterogeneity in contact patterns as a function of symptom severity.

Additionally, whilst our intent is for our model to be generalisable to other respiratory pathogens for which symptom propagation is possible, the assumptions made in this paper may not be well suited to all such pathogens. In particular, the assumption that severe disease is more transmissible and has a longer infectious period may not hold. If this assumption were removed and mild and severe disease were to produce the same number of secondary cases, we would expect the proportion of cases that were severe to be similar regardless of the strength of symptom propagation.

Second, definitions of severity of infection can vary appreciably, showcased by prior work on the two pathogens focused upon in this study (influenza and SARS-CoV-2) [34, 40, 49, 50]. An inevitable consequence was there being uncertainty in quantifying health economic parameters (such as QALY losses and hospitalisation rates) for our “mild” and “severe” infection categorisations. Creating a formal definition of severity in this context requires a deeper biological understanding of how symptom propagation occurs and, in turn, further research. We would encourage a conceptual re-analysis of the symptoms of respiratory infections from a clinical standpoint, leading to a new framework for categorising clinical outcomes informed by an understanding not only of patient-level symptoms, but also symptom propagation and its implications for onward transmission, along with the formulation of associated data collection protocols. Indeed, although we have chosen to focus on only two severity classes, this simplification may not always be appropriate. An extension of this model to include a separate asymptomatic class or a continuum of severity could be explored in future work (or even qualitatively different symptom sets that do not straightforwardly map to levels of severity), with model structural choices informed by data where appropriate.

We also recognise that there are many factors not included in this model which are known to affect symptom severity, and future work could extend the framework presented here to incorporate characteristics such as age, immune status and multiple strains. Age structure could be of particular interest because it has previously been suggested that the combination of age-dependent mixing and age-dependent severity might cause correlations between the severity of the infector and the infectee [40]. As such, the inclusion of age structure could amplify the effects of symptom propagation. Also, as a consequence of not modelling demographic characteristics, we assumed intervention uptake to be uniform across the population. In reality, those identified as risk groups, such as healthcare workers or immuno-compromised people, are likely to be targeted first as part of any intervention policy, as seen in the vaccine roll-out during the COVID-19 pandemic [78]. We expect that such targeting would amplify the increase in intervention effectiveness caused by strong symptom propagation and anticipate this effect could be seen for all intervention types, even purely infection-blocking interventions. Further work is required to investigate these potential dependencies.

In addition to extensions to the deterministic, compartmental ODE model used in this paper, the model framework could be applied to a range of model types [47]. If applied to a stochastic model at a localised spatial level (population size of the order of hundreds rather than millions), we could expect symptom propagation to result in a large variation in the proportion of cases that are severe, depending on the severity of the initial cases. We would likely find that, even for relatively weak symptom propagation, a stochastic model may generate large variation in the proportion of cases that are severe, with the potential for outbreaks to be predominantly severe. The symptom propagation model framework could also be applied to network or spatial models. In these cases, we might expect symptom propagation to result in large spatial heterogeneity in the severity of (local) outbreaks, leading to increased strain on local healthcare services despite the larger-scale outbreak severity being similar to what is predicted by a model with no spatial structure.

Another identified key area of future work is the estimation of the *α* parameter, i.e. the strength of symptom propagation. However, due to the complex nature of symptom severity and the many confounding factors, performing this inference is non-trivial. A large volume of individual data with both information on symptom severity and who infected whom is required. Major challenges include separating symptom propagation from the effects of strains and from the impact of genetic similarity between an individual and the person who infected them (e.g. in the case of related individuals). Nevertheless, the public health benefits of such estimations will make surmounting such challenges rewarding, including informing the relative importance of transmission from those who are asymptomatic (and therefore the optimal approach for contact tracing) and the role of vaccines that may reduce symptom severity as well as infection burden. Close dialogue with appropriate data holders will be a crucial aspect to successfully accomplish these goals.

In summary, these findings demonstrate the importance of including symptom propagation in models of infectious disease transmission to assist decision makers in planning infection control and mitigation strategies, where insights on epidemiological and health economic implications of possible actions are required and where there is evidence to support the presence of symptom propagation for a given pathogen. There are still questions around whether, and to what extent, symptom propagation occurs for various pathogens and, although evidence in the literature supporting symptom propagation is accumulating, we believe it would be beneficial to reduce the uncertainty around this topic. We conclude that the consideration of symptom propagation should be commonplace in the modelling of infectious diseases and in evaluating proposed control policies from a health economics perspective. To heighten the robustness of future modelling analyses, this motivates data collection to promote the use of data-driven models and the development of analytic methods to identify the extent of symptom propagation (i.e. the value of *α*) for pathogens of concern.

## Author contributions

**Phoebe Asplin:** Conceptualisation, Data curation, Formal analysis, Investigation, Methodology, Software, Validation, Visualisation, Writing - Original Draft, Writing - Review & Editing.

**Matt J. Keeling:** Conceptualisation, Methodology, Supervision, Visualisation, Writing - Review & Editing.

**Rebecca Mancy:** Conceptualisation, Methodology, Supervision, Visualisation, Writing - Review & Editing.

**Edward M. Hill:** Conceptualisation, Methodology, Software, Supervision, Visualisation, Writing - Original Draft, Writing - Review & Editing.

## Acknowledgements

We thank Tom Finnie and Fergus Cumming for their helpful comments on the manuscript. We also thank Daniel Haydon, Ciaran McMonagle, Alessandro Vespignani, Jaime Earnest and Bruno Goņcalves for useful discussions of earlier versions of the ideas in this manuscript.

## Data availability

All data utilised in this study are publicly available, with relevant references and data repositories provided.

## Code availability

The code repository for the study is available at:

https://github.com/pasplin/symptom-propagation-mathematical-modelling. Archived code: https://doi.org/10.5281/zenodo.10560986

## S1 Calculating the basic reproduction number, R_0_, and calibrating *β*

The basic reproductive number, *R*_0_, is commonly used in epidemiology to indicate a disease’s potential to spread through a population. It is defined as the average number of secondary cases generated by an average infectious individual in a fully susceptible population. *R*_0_ can be formulated as

*R*_0_ = (average number of infections generated per unit time) *×* (average infectious period).

We calculated the value of *R*_0_ using the next-generation matrix (NGM) approach. This method was developed by Diekmann *et al.* [1] to calculate the value of *R*_0_ in heterogeneous populations where compartments are split into a finer structure (e.g. into age classes). The next generation matrix is defined to be

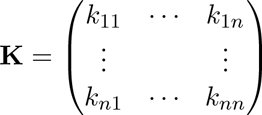

where *k_ij_* is the average number of type-i cases generated by a type-j case in a fully susceptible population. This can be applied to the ODE model by considering a vector containing the number of people in each infectious class, which then multiplies by **K** at each time step. This vector grows at a rate given by the dominant eigenvalue of **K** - the eigenvalue with the largest absolute value - with *R*_0_ being this dominant eigenvalue. In the case of our model,

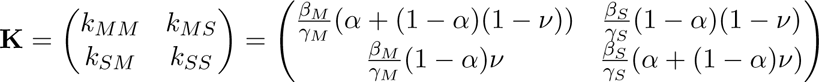

*R*_0_ is an eigenvalue of **K** and thus solves

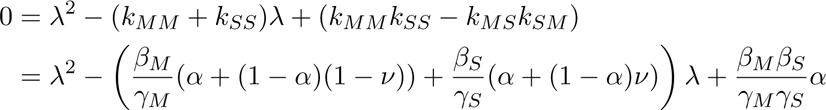

If *β_M_* = *β*, *β_S_* = *rβ*, then the derived equation for *R*_0_ simplifies to

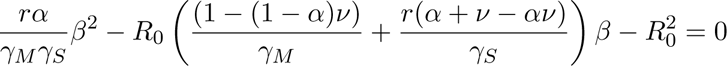

from which, for a given value of *R*_0_, we calculated the required value of *β*.

## S2 Results with fixed *β*

In the main text we fixed the value of *R*_0_ for each parameter set by choosing an appropriate value of *β* for each value of *α*. Here we show our results with fixed *β* instead. For each of the three disease parameterisations we chose *β_M_* to give the stated value of *R*_0_ when *α* = 0: *R*_0_ = 1.5 for seasonal influenza, *R*_0_ = 3.0 for pandemic influenza and SARS-CoV-2.

We found that the value of *R*_0_ increased with *α* (Fig. S1) and similarly with the total outbreak size and peak prevalence (Fig. S2). As with our main text results, the proportion of cases that were severe increased with *α* (Fig. S2). However, the outbreak duration decreased with *α* (Fig. S2)

**Figure S1:**
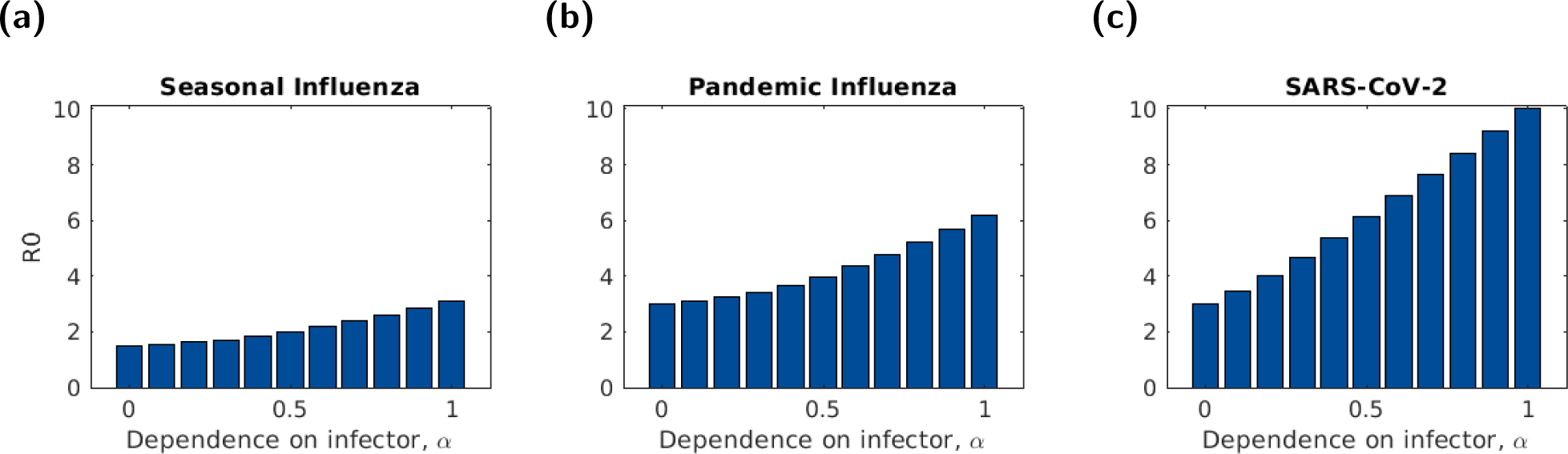
The basic reproduction number, *R*_0_, for three disease parameterisations, plotted for different symptom propagation strengths, *α* and fixed values of *β*. In all panels, *ν* = 0.2. The three disease parameterisations used were: **(a)** Seasonal influenza; **(b)** Pandemic influenza; **(c)** SARS-CoV-2, with parameters as given in Table 1. For each of the three disease parameterisations we chose *β_M_* to give the stated value of *R*_0_ when *α* = 0: *R*_0_ = 1.5 for seasonal influenza, *R*_0_ = 3.0 for pandemic influenza and SARS-CoV-2.

## S3 Health economic model: Parameterisation details

### S3.1 Likelihood of hospitalisation and death

Hospitalisation and death rates were taken from Hill *et al.* [2] for influenza and Moran *et al.* [3] for SARS-CoV-2.

### S3.2 Quality-adjusted life years (QALYs)

We assumed that mild disease had sufficiently minimal symptoms, thereby causing no QALY losses.

We took influenza QALY losses for severe, non-fatal cases were taken from Hill *et al.* [2]. For SARS-CoV-2, we used estimates for years lived with disability (YLD) per case from Moran *et al.* [3]. We used the YLD for “severe” (non-ICU) and “critical” (ICU) hospitalised cases to estimate QALY losses for non-fatal hospitalised cases. We used the YLD for “moderate” and “post-acute consequences” to estimate QALY losses for severe non-hospitalised cases.

The QALY losses per death depend primarily on the number of healthy years lost and therefore the age of the individual. Our model was not stratified by age class, so to estimate the average QALY losses per death, we used estimates of the total QALY losses from deaths during an outbreak and divided by the number of deaths. For influenza, we used data from an outbreak in Spain during the 2009 H1N1 pandemic [4]. For SARS-CoV-2, we used a study of COVID-19 cases in the Republic of Ireland from March 2020 to February 2021 [3].

**Figure S2:**
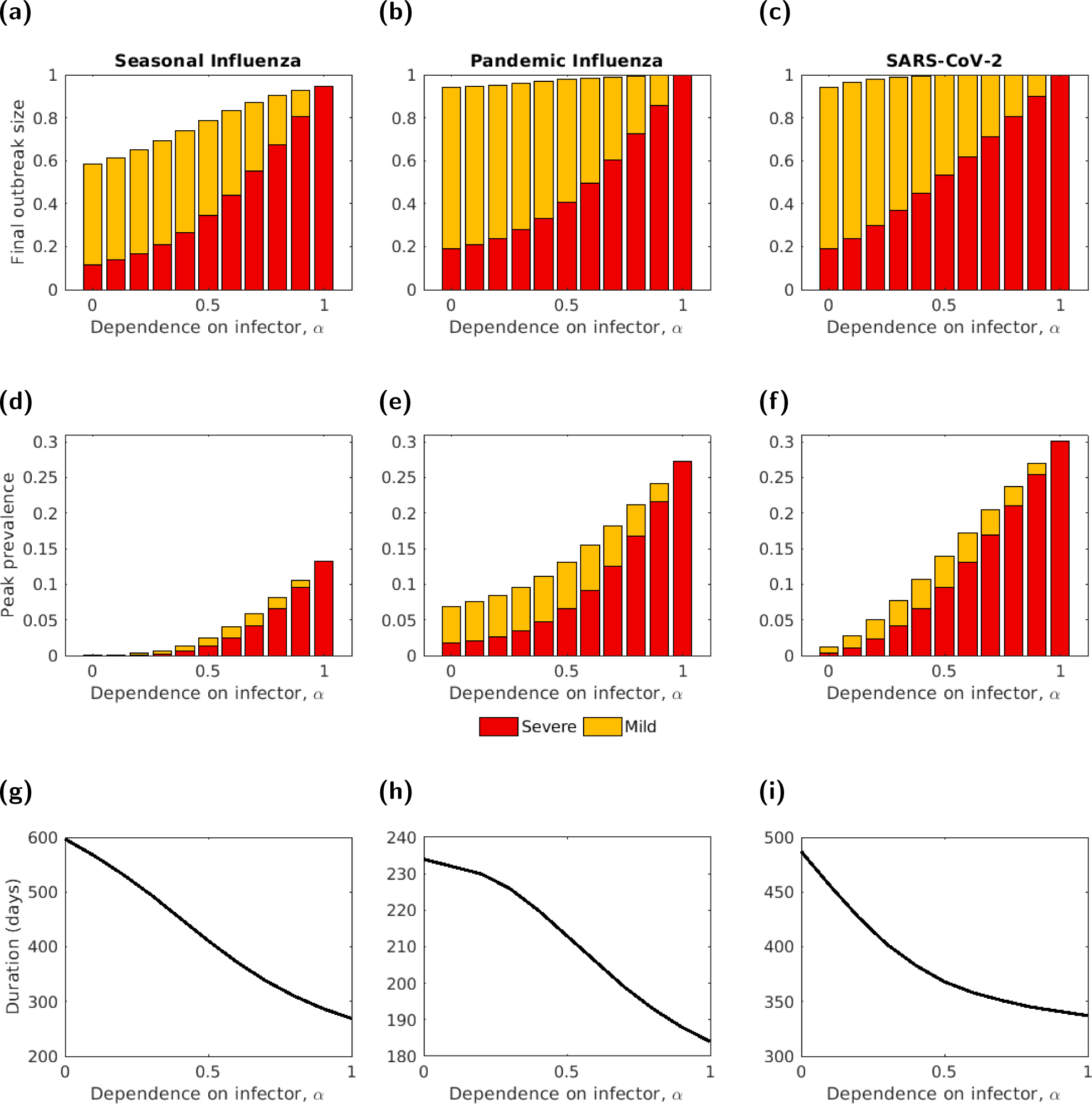
The final outbreak size, peak prevalence and outbreak duration, by severity, for three disease parameterisations, plotted for different symptom propagation strengths, *α* and fixed values of *β*. **(a-c)** Final outbreak size by severity. **(d-f)** Peak prevalence by severity. The intensity of the shading denotes the symptom severity class, with severe cases in red and mild cases in yellow. **(g-i)** Outbreak duration (note the different y-axis scales). In all panels, *ν* = 0.2. The three disease parameterisations used were: **(a,d,g)** Seasonal influenza; **(b,e,h)** Pandemic influenza; **(c,f,i)** SARS-CoV-2, with parameters as given in Table 1. For each of the three disease parameterisations we chose *β_M_* to give the stated value of *R*_0_ when *α* = 0: *R*_0_ = 1.5 for seasonal influenza, *R*_0_ = 3.0 for pandemic influenza and SARS-CoV-2.

### S3.3 Monetary costs

Hospital costs for influenza were taken from Hill *et al.* [2]. Hospital costs for SARS-CoV-2 were estimated under the assumption that costs would be twice as much as influenza, due to the average length of stay being roughly twice as long for SARS-CoV-2 (around 13 days [5]) than for influenza (around 7 days [6]).

### S3.4 Discounting

A common component of health economic modelling is discounting, which assigns a lower value to costs and health outcomes that occur in the future [7]. For the scenarios where we applied a discounting rate of 3.5%, the discounted value (of QALYs lost or monetary costs, as appropriate) in year *y* was given by

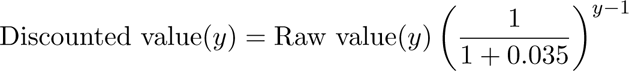

## S4 Alternative infection-blocking and modified breakthrough infection intervention

Throughout this section, we modelled an infection-blocking vaccine for which breakthrough infections were only possible when the infector was a severe case (Fig. S3).

**Figure S3:**
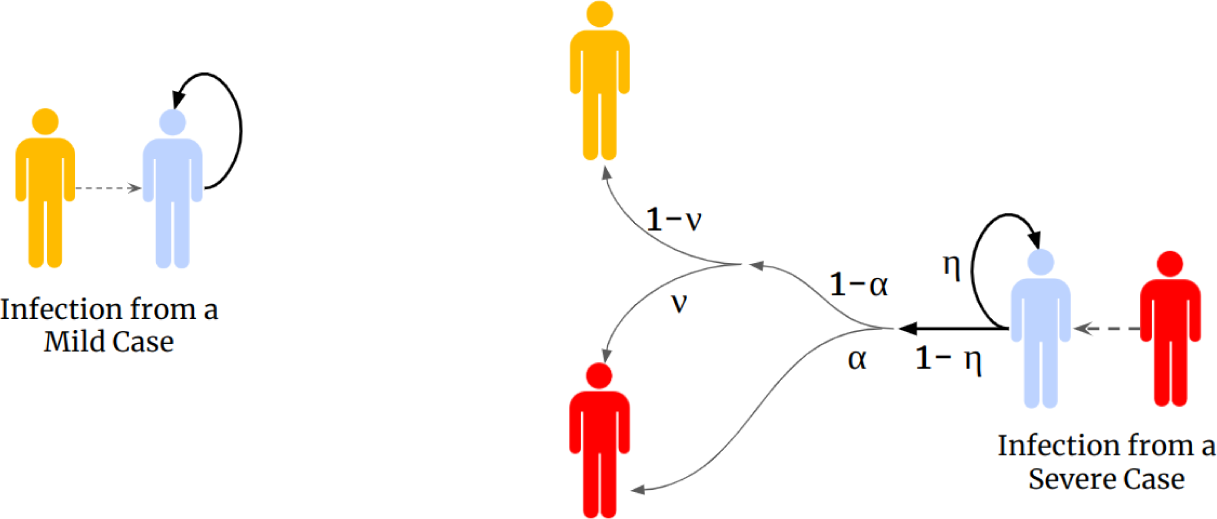
Schematic of the infection-blocking and modified breakthrough infection vaccine. If challenged by infection from a mild case, a vaccinated individual would be guaranteed to be protected. If challenged by infection from a severe case, a vaccinated individual had a probability *η* of their infection being prevented and a probability 1 *− η* of being infected, with their symptom severity then being determined as usual. Yellow shaded individuals correspond to infectious cases with mild symptoms, red shaded individuals correspond to infectious cases with severe symptoms and blue shaded individuals correspond to those who are vaccinated. The values on the arrows show the corresponding probability.

### S4.1 Modified model equations

Under the use of such an intervention, the model dynamics are governed by the following system of ODEs. Recall that the *V* class denotes those who were susceptible and vaccinated, whilst we denote with red font those terms that include the action of the intervention:

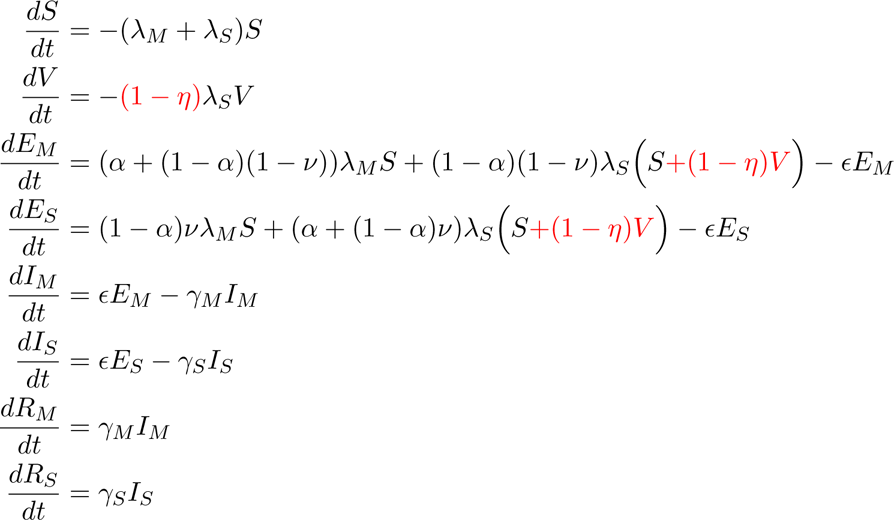

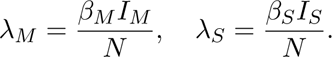

### S4.2 Simulation overview

As for the interventions studied in the main text, we ran a range of values of *α* and *ν* (both between 0 and 1, with an increment of 0.02); for each combination of *α* and *ν*, *β* was chosen to fix *R*_0_ at the desired value in the no intervention case.

Performing analogous methods as for the other interventions, per *α*-*ν* pair we ascertained the vaccine uptake that maximised the intervention unit threshold value. We also explored the model’s sensitivity to the vaccine efficacy, in this case modulating the risk of a breakthrough infection by a vaccinated, severe infection case.

### S4.3 Summary of findings

Across the collection of results, we report no difference in the outcomes between the (solely) infection blocking vaccine and the infection blocking vaccine where only severe cases could cause breakthrough infections (Figs. S4 and S5).

**Figure S4:**
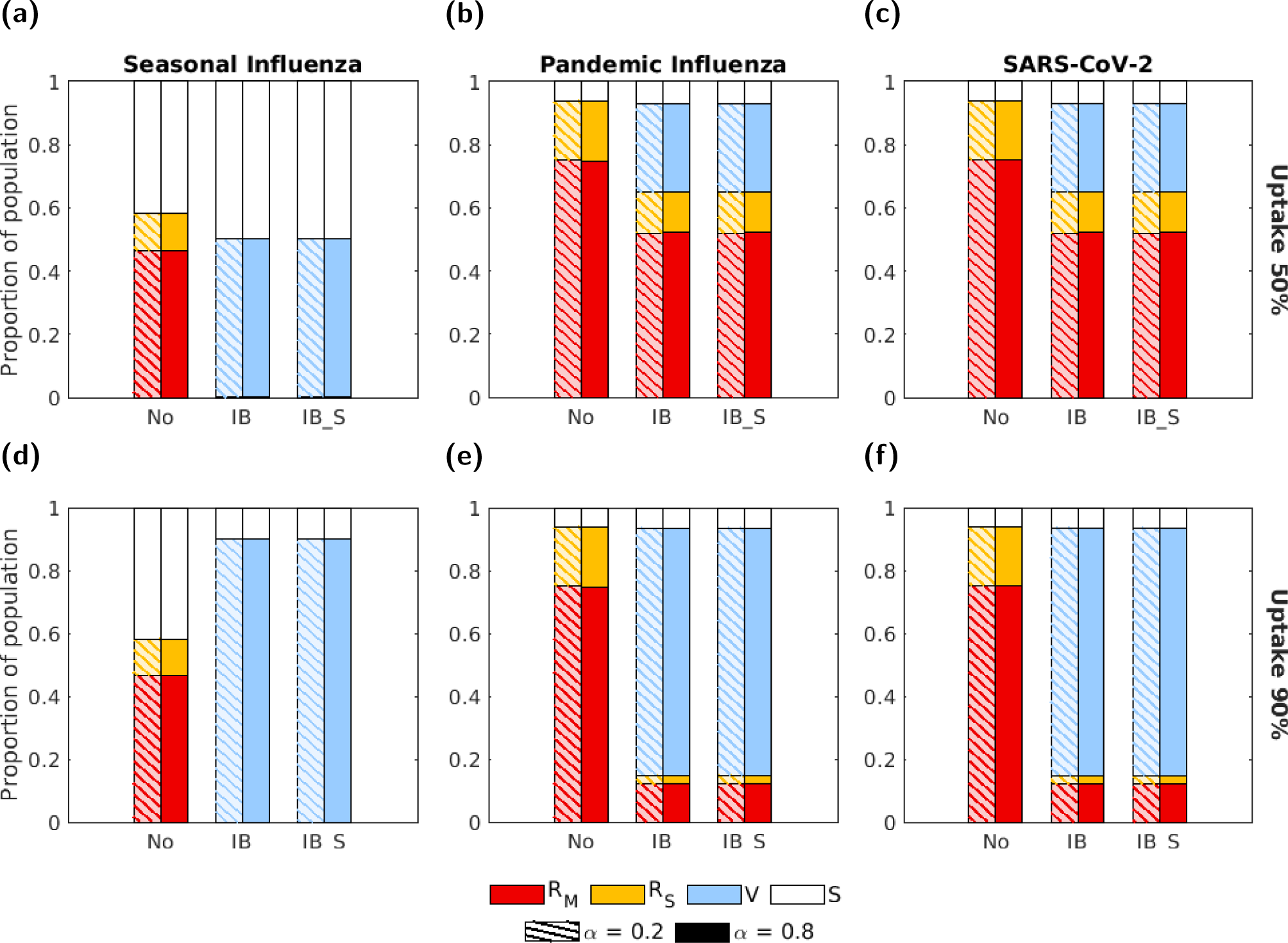
Proportion of the population in each disease state at the end of the outbreak for each disease parameterisation given use of an infection blocking vaccine where only severe cases can cause breakthrough infections. The three groups of bars correspond to three intervention scenarios: no intervention (No), an infection blocking vaccine (IB) and an infection blocking vaccine for which only severe cases can cause breakthrough infections (IB S). The two bars in each group correspond to symptom propagation strengths of *α* = 0.2 (left bar, hatched lines) and *α* = 0.8 (right bar, solid fill). Bar shading corresponds to disease status: red - recovered from severe infection (*R_S_*); yellow - recovered from mild infection (*R_M_*); blue - susceptible and vaccinated (*V*); white - susceptible and not vaccinated (*S*). The two rows correspond to two vaccine uptake levels: **(a-c)** 50%; **(d-f)** 90%. Columns correspond to differing disease parameterisations: **(a,d)** seasonal influenza; **(b,e)** pandemic influenza; **(c,f)** SARS-CoV-2. We fixed the vaccine efficacy at 70% and all other parameters were as given in Table 1, with *ν* chosen to fix the proportion of cases that were severe equal to 0.8.

**Figure S5:**
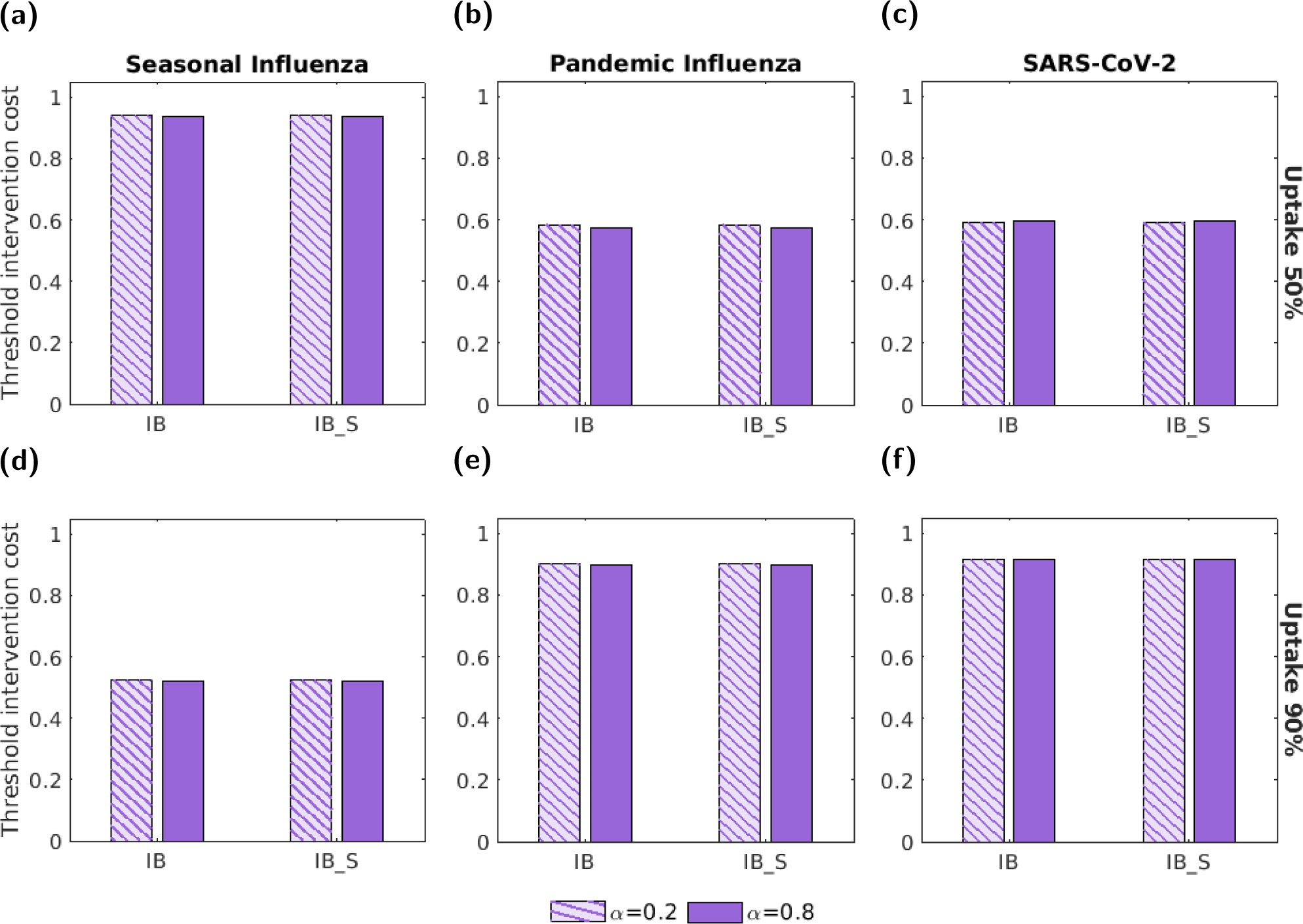
The threshold unit intervention cost, under each disease parameterisation, for an infection blocking vaccine for which only severe cases can cause breakthrough infections. We normalise the threshold unit intervention cost by the highest absolute threshold unit intervention cost attained across the range of tested vaccine uptake values. The two groups of bars correspond to an infection blocking vaccine (IB) and an infection blocking vaccine for which only severe cases can cause breakthrough infections (IB S). The two bars in each group correspond to symptom propagation strengths of *α* = 0.2 (left bar, hatched lines) and *α* = 0.8 (right bar, solid fill). The two rows correspond to two vaccine uptake levels: **(a-c)** 50%; **(d-f)** 90%. Columns correspond to differing disease parameterisations: **(a,d)** seasonal influenza; **(b,e)** pandemic influenza; **(c,f)** SARS-CoV-2. The efficacy was fixed at 70% and all other parameters were as given in Table 1 with *ν* chosen to fix the proportion of cases that were severe equal to 0.8.

## S5 Additional epidemiological outcomes

### S5.1 Final outbreak size and peak prevalence

**Figure S6:**
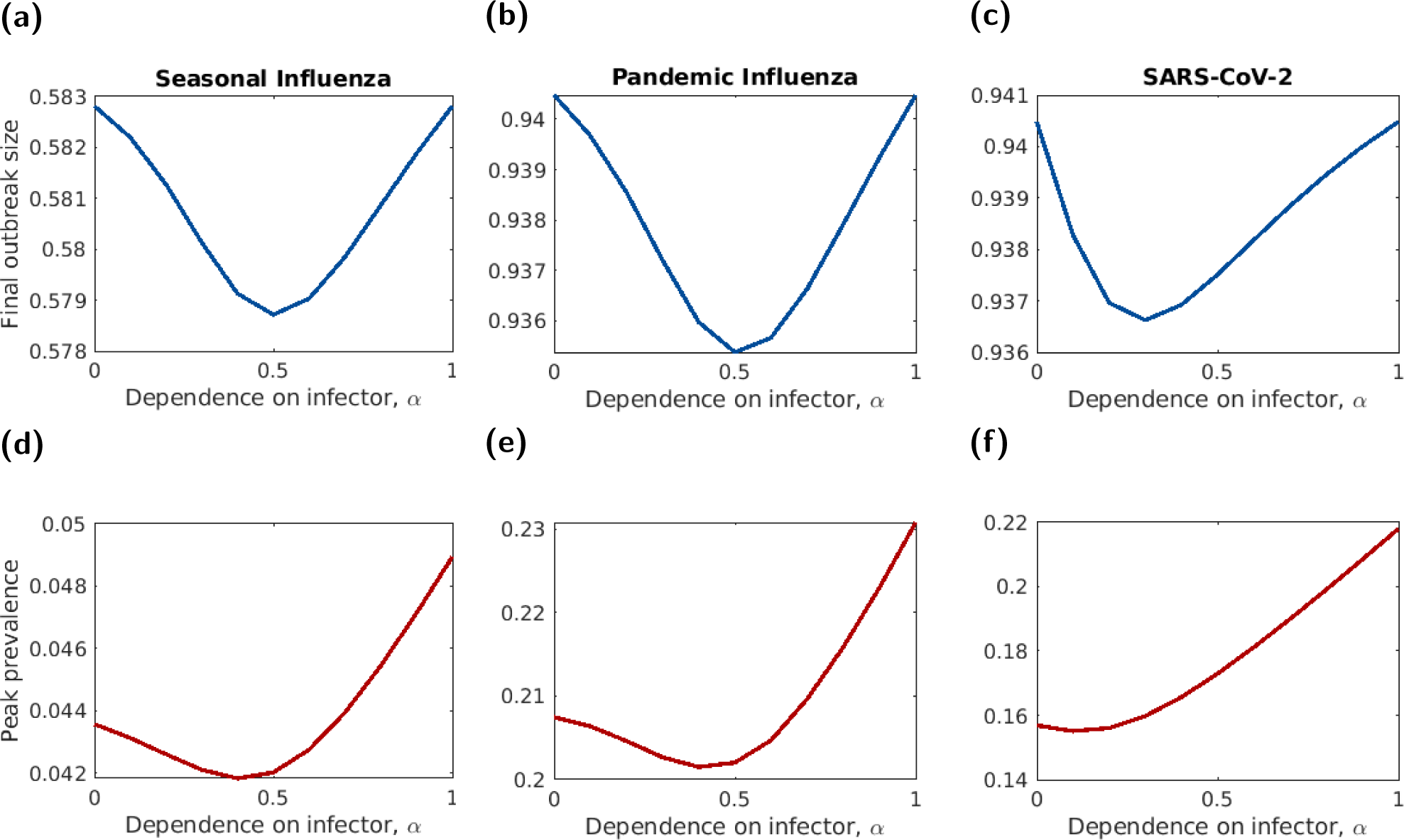
The final outbreak size and peak prevalence for three parameter sets whilst varying the dependence on the infector’s symptom severity, *α*. The top row shows the total proportion of the population infected across both severity classes. The bottom row shows the proportion of the population infected across both severity classes at the peak of the outbreak. Note that the y-axis scale varies between plots in the same row.

### S5.2 Temporal intervention plots

**Figure S7:**
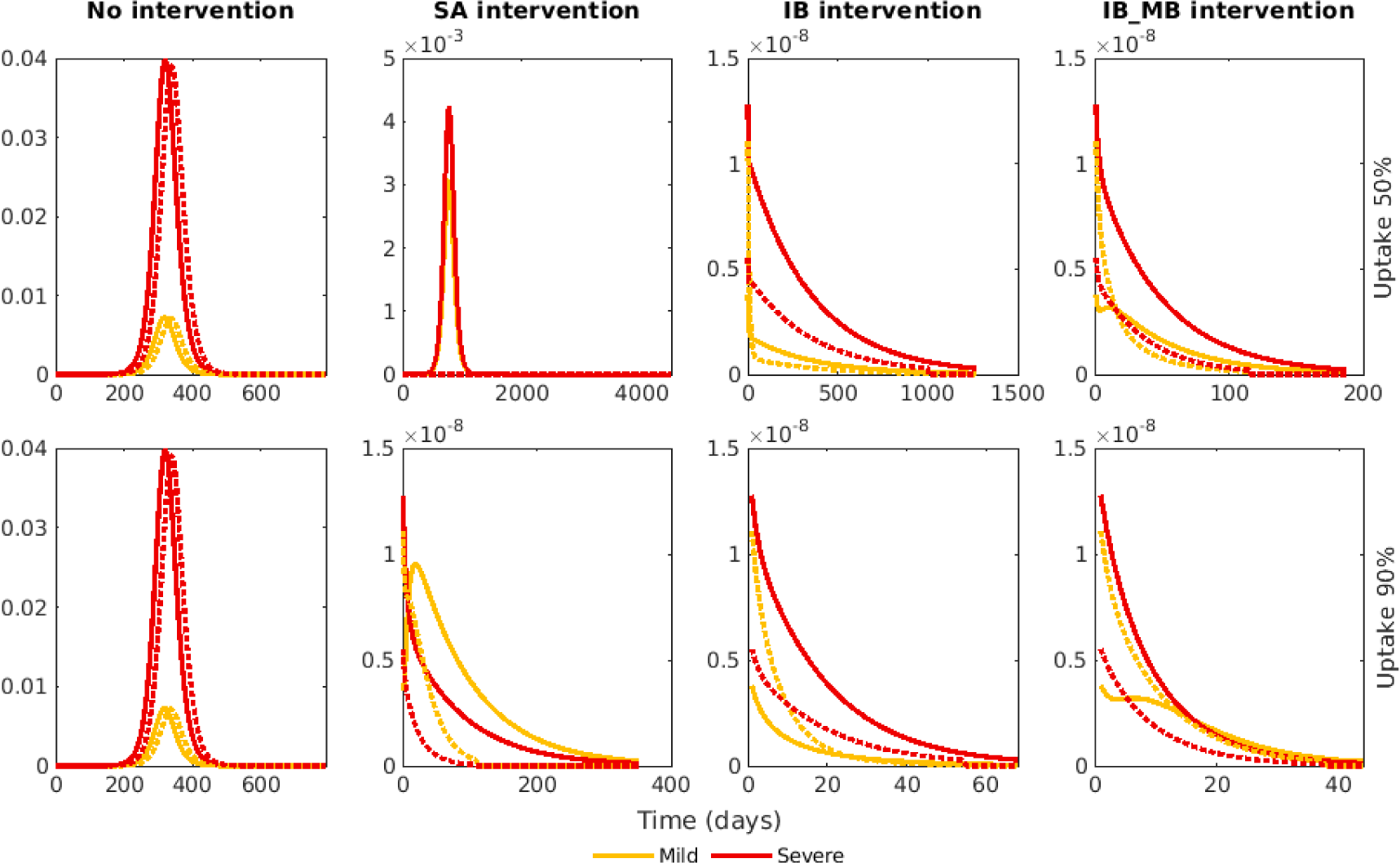
Temporal profiles of infection prevalence in the absence of interventions and our three types of intervention for a pathogen with seasonal influenza-like attributes. The four columns correspond to four intervention scenarios: no intervention, a symptom-attenuating vaccine, an infection-blocking vaccine and an infection-blocking vaccine that only admits mild break-through infections. The rows correspond to two vaccine uptake levels, 50% and 90%. We fixed the vaccine efficacy at 70%. The line colour denotes the disease severity, with the pink line showing the proportion of the population who were infectious with mild disease and the red line showing the proportion of the population who were infectious with severe disease. The line style denotes the value of *α*, with the solid lines corresponding to *α* = 0.2 and the dotted lines corresponding to *α* = 0.8. Throughout, *ν* was chosen to fix the proportion of cases that were severe equal to 0.8. Note the differing scales on the y-axis.

**Figure S8:**
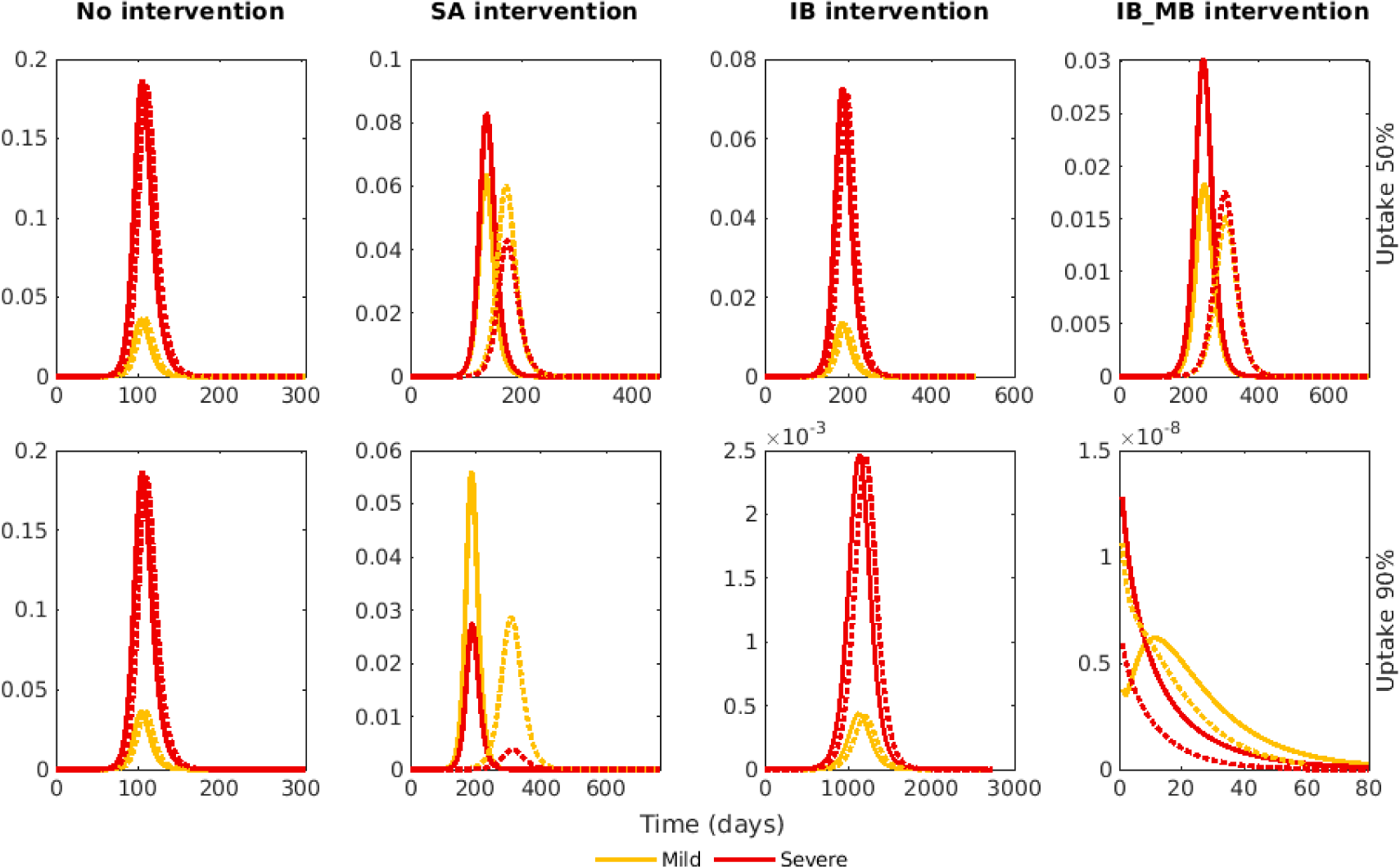
Temporal profiles of infection prevalence in the absence of interventions and our three types of intervention for a pathogen with pandemic influenza-like attributes. The four columns correspond to four intervention scenarios: no intervention, a symptom-attenuating vaccine, an infection-blocking vaccine and an infection-blocking vaccine that only admits mild break-through infections. The rows correspond to two vaccine uptake levels, 50% and 90%. We fixed the vaccine efficacy at 70%. The line colour denotes the disease severity, with the pink line showing the proportion of the population who were infectious with mild disease and the red line showing the proportion of the population who were infectious with severe disease. The line style denotes the value of *α*, with the solid lines corresponding to *α* = 0.2 and the dotted lines corresponding to *α* = 0.8. Throughout, *ν* was chosen to fix the proportion of cases that were severe equal to 0.8. Note the differing scales on the y-axis.

**Figure S9:**
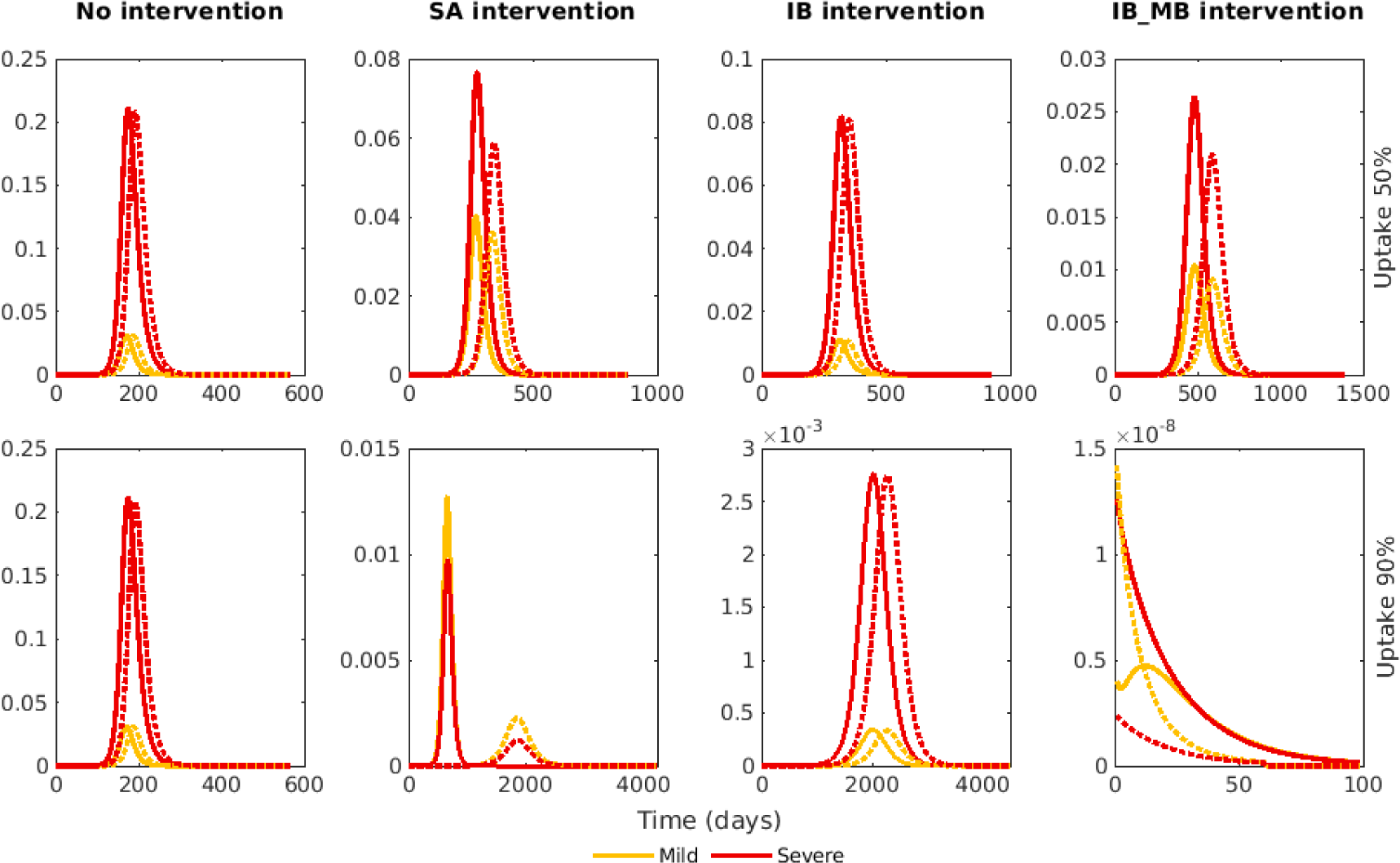
Temporal profiles of infection prevalence in the absence of interventions and our three types of intervention for a pathogen with SARS-CoV-2-like attributes. The four columns correspond to four intervention scenarios: no intervention, a symptom-attenuating vaccine, an infection-blocking vaccine and an infection-blocking vaccine that only admits mild breakthrough infections. The rows correspond to two vaccine uptake levels, 50% and 90%. We fixed the vaccine efficacy at 70%. The line colour denotes the disease severity, with the pink line showing the proportion of the population who were infectious with mild disease and the red line showing the proportion of the population who were infectious with severe disease. The line style denotes the value of *α*, with the solid lines corresponding to *α* = 0.2 and the dotted lines corresponding to *α* = 0.8. Throughout, *ν* was chosen to fix the proportion of cases that were severe equal to 0.8. Note the differing scales on the y-axis.

### S5.3 Additional intervention bar plots

**Figure S10:**
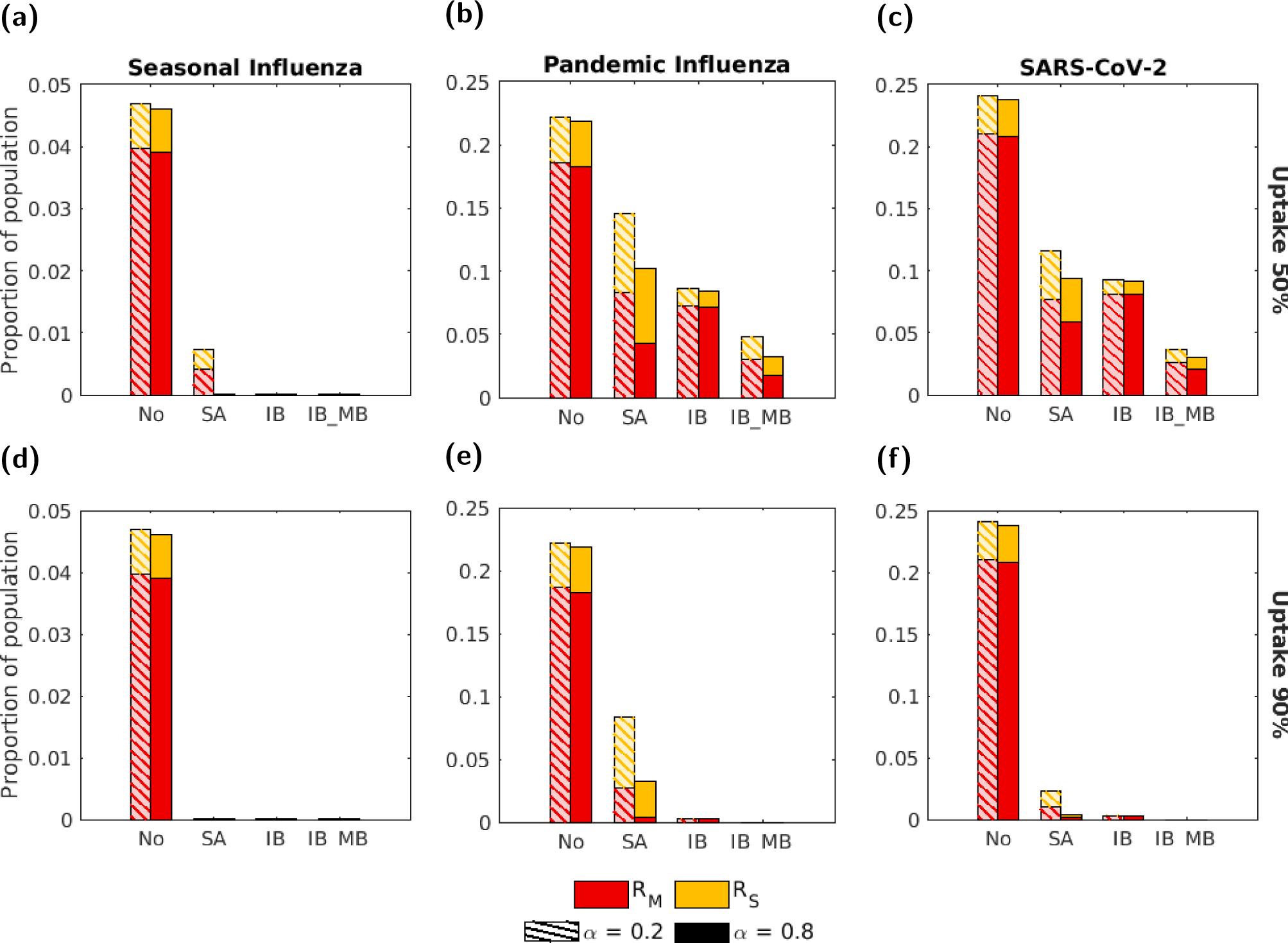
Impact of disease parameterisation, action of intervention and intervention efficacy on peak infection prevalence. The four groups of bars correspond to four intervention scenarios: no intervention (No), a symptom attenuating vaccine (SA), an infection blocking vaccine (IB) and an infection blocking vaccine which only admits mild breakthrough infections (IB MB). The two bars in each group correspond to symptom propagation strengths of *α* = 0.2 (left bar, hatched lines) and *α* = 0.8 (right bar, solid fill). Bar heights correspond to the peak in infection prevalence, with red and yellow bars representing severe and mild cases, respectively. The two rows correspond to two levels of vaccine uptake: **(a-c)** 50%; **(d-f)** 90%. The columns corresponded to one of the disease parameterisations: **(a,d)** seasonal influenza; **(b,e)** pandemic influenza; **(c,f)** SARS-CoV-2. We fixed the vaccine efficacy at 70%. All other parameters were as given in Table 1 with *ν* chosen to fix the proportion of cases that were severe equal to 0.8.

**Figure S11:**
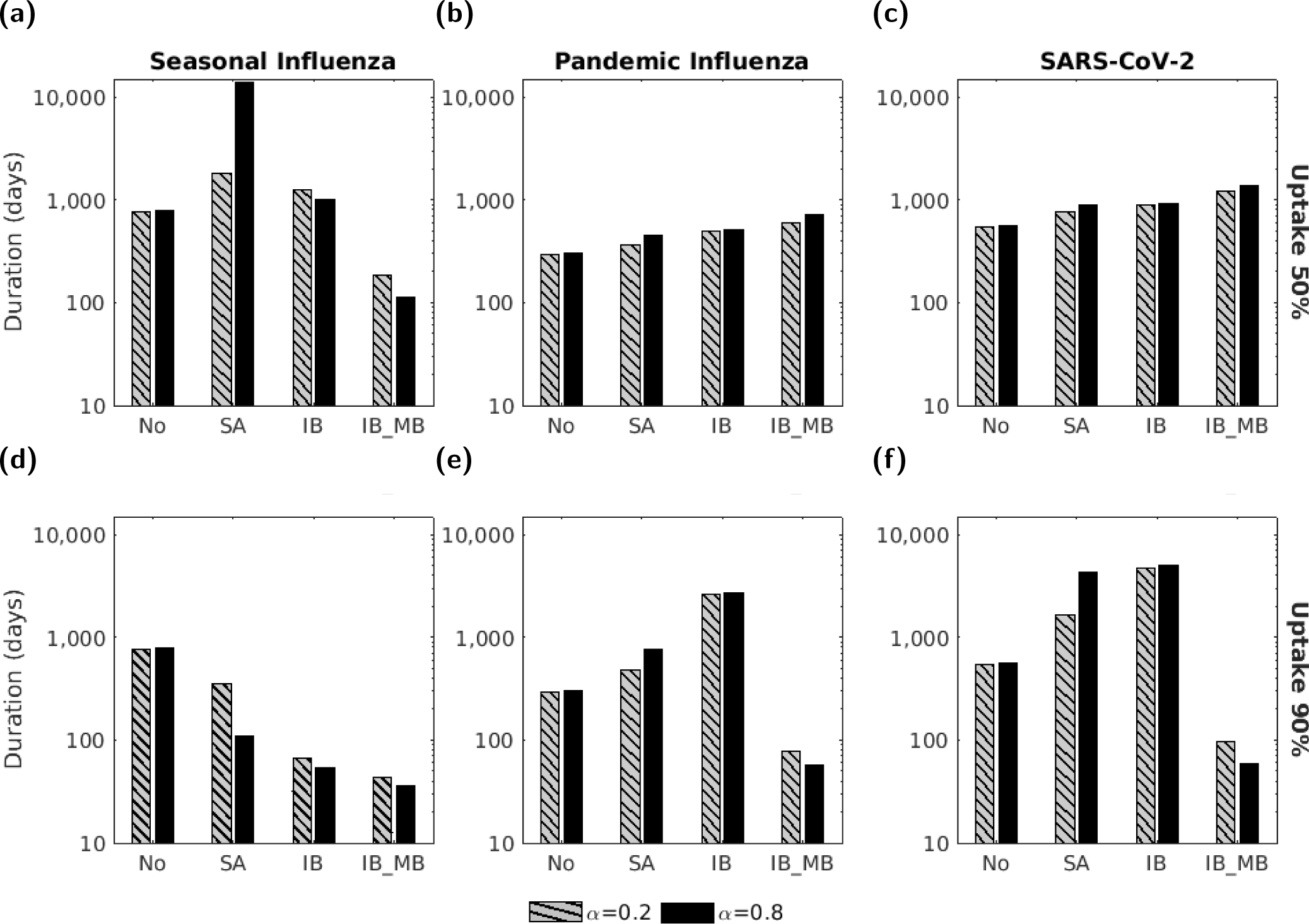
Impact of disease parameterisation, action of intervention and intervention efficacy on outbreak duration. In each panel, the four groups of bars correspond to four intervention scenarios: no intervention (No), a symptom attenuating vaccine (SA), an infection blocking vaccine (IB) and an infection blocking vaccine which only admits mild breakthrough infections (IB MB). The two bars in each group correspond to symptom propagation strengths of *α* = 0.2 (left bar, hatched lines) and *α* = 0.8 (right bar, solid fill). Bar heights correspond to outbreak duration in days. The two rows correspond to two levels of vaccine uptake: **(a-c)** 50%; **(d-f)** 90%. The columns corresponded to one of the disease parameterisations: **(a,d)** seasonal influenza; **(b,e)** pandemic influenza; **(c,f)** SARS-CoV-2. We fixed the vaccine efficacy at 70%. All other parameters were as given in Table 1 with *ν* chosen to fix the proportion of cases that were severe equal to 0.8.

### S5.4 Outcomes conditional on *ν* = 0.2

**Figure S12:**
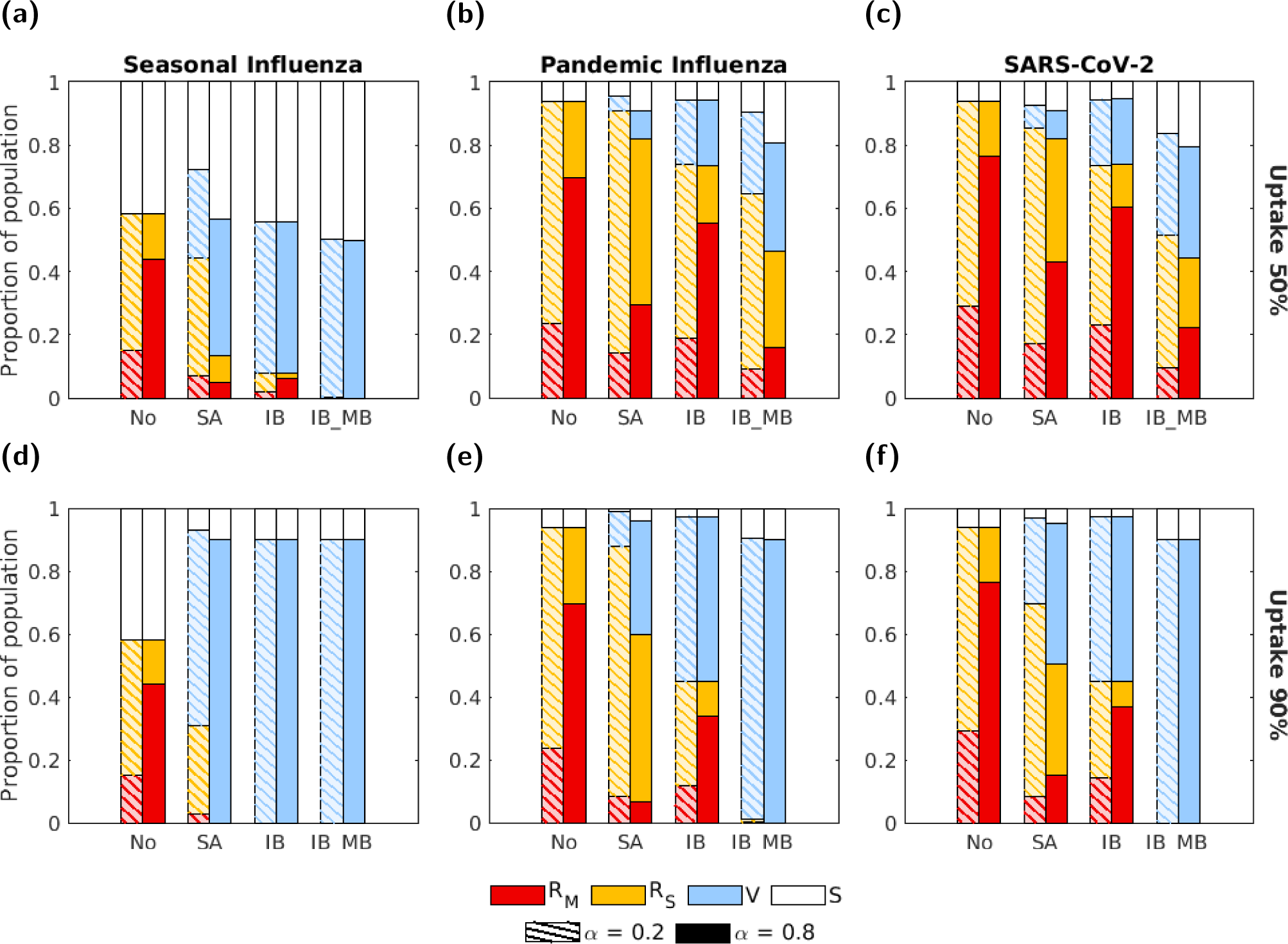
End of outbreak disease status composition under an alternative baseline severity assumption. Analogous to Fig. 5 in the main manuscript, except here we used a fixed value of *ν* = 0.2 (instead of calibrating the value of *ν* to attain 80% of cases being severe in the no intervention scenario). In each panel, the four groups of bars correspond to four intervention scenarios: no intervention (No), a symptom attenuating vaccine (SA), an infection blocking vaccine (IB) and an infection blocking vaccine which only admits mild breakthrough infections (IB MB). The two bars in each group correspond to symptom propagation strengths of *α* = 0.2 (left bar, hatched lines) and *α* = 0.8 (right bar, solid fill). The shading of the bar corresponds to four disease status compartments: red - recovered from severe infection (*R_S_*); yellow - recovered from mild infection (*R_M_*); blue - susceptible and vaccinated (*V*); white - susceptible and not vaccinated (*S*). The two rows correspond to two uptake levels: **(a-c)** 50%; **(d-f)** 90%. Columns correspond to differing disease parameterisations: **(a,d)** seasonal influenza; **(b,e)** pandemic influenza; **(c,f)** SARS-CoV-2. We fixed the vaccine efficacy at 70% and all other parameters were as given in Table 1.

**Figure S13:**
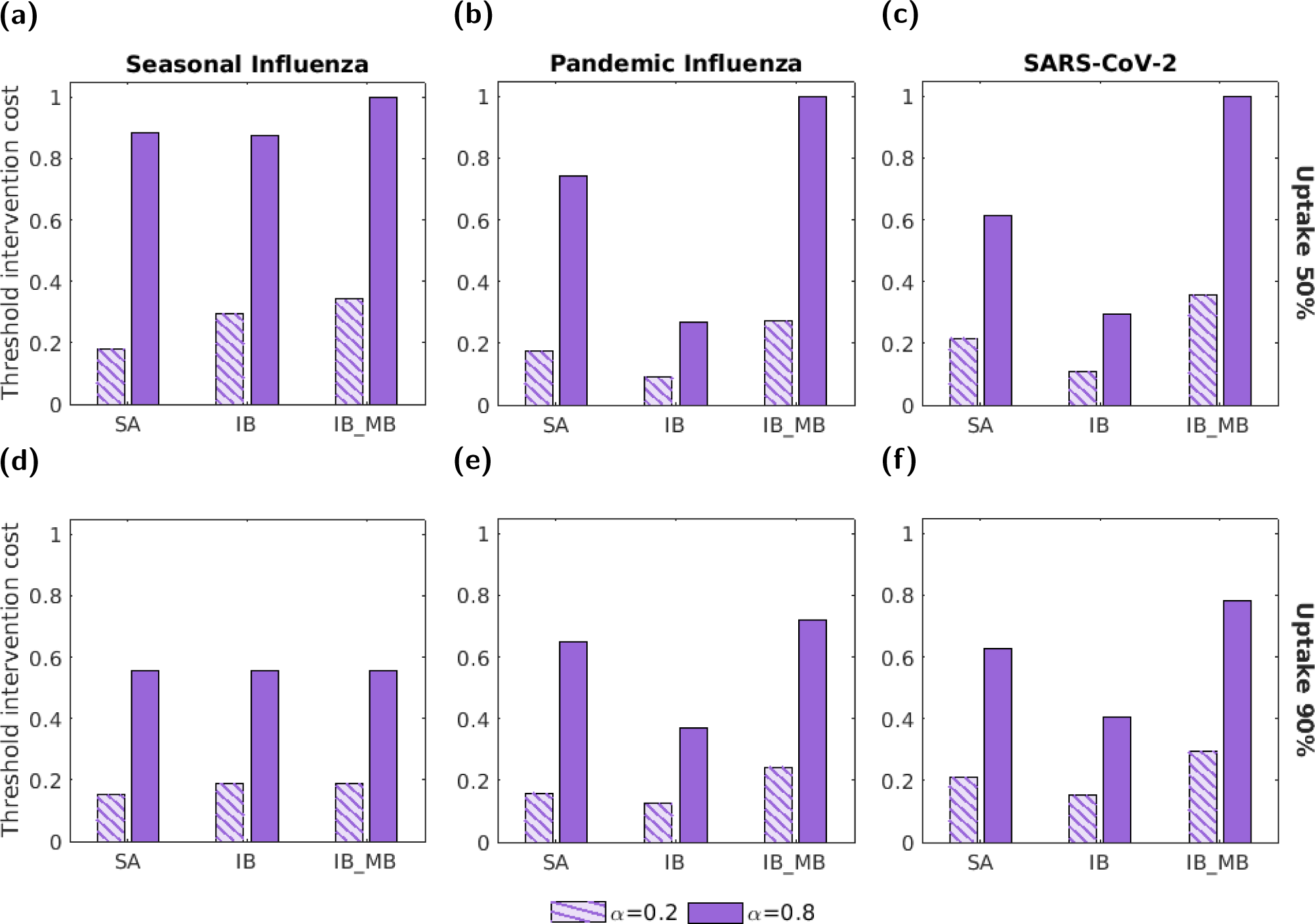
Threshold unit intervention costs for three interventions and three disease parameterisations under an alternative value of *ν*. Analogous to Fig. 7 in the main manuscript, except here we used a fixed value of *ν* = 0.2 (instead of calibrating the value of *ν* to attain 80% of cases being severe in the no intervention scenario). In all panels, we normalised threshold unit intervention costs by the highest absolute threshold unit intervention cost attained across the range of tested intervention uptake values. The three groups of bars correspond to three interventions: a symptom attenuating vaccine (SA), an infection blocking vaccine (IB) and an infection blocking vaccine that only admits mild breakthrough infections (IB MB). The two bars in each group correspond to symptom propagation strengths of *α* = 0.2 (left bar, hatched lines) and *α* = 0.8 (right bar, solid fill). The two rows correspond to two vaccine uptake levels: **(a-c)** 50%; **(d-f)** 90%. Columns correspond to differing disease parameterisations: **(a,d)** seasonal influenza; **(b,e)** pandemic influenza; **(c,f)** SARS-CoV-2. Vaccine efficacies were fixed at 70% and all other parameters were as given in Table 1.

## S6 Sensitivity to discounting

Whilst in the main text we used a discount rate of 3.5% on monetary costs and QALY losses, here we performed similar health economic analyses with no discounting applied (i.e. a discounting rate of 0%).

Overall, when applying no discounting, we found quantitatively similar findings (as in the main manuscript) for the relative threshold unit intervention costs for each combination of disease parameterisation and action of intervention (Figs. S14 and S15).

**Figure S14:**
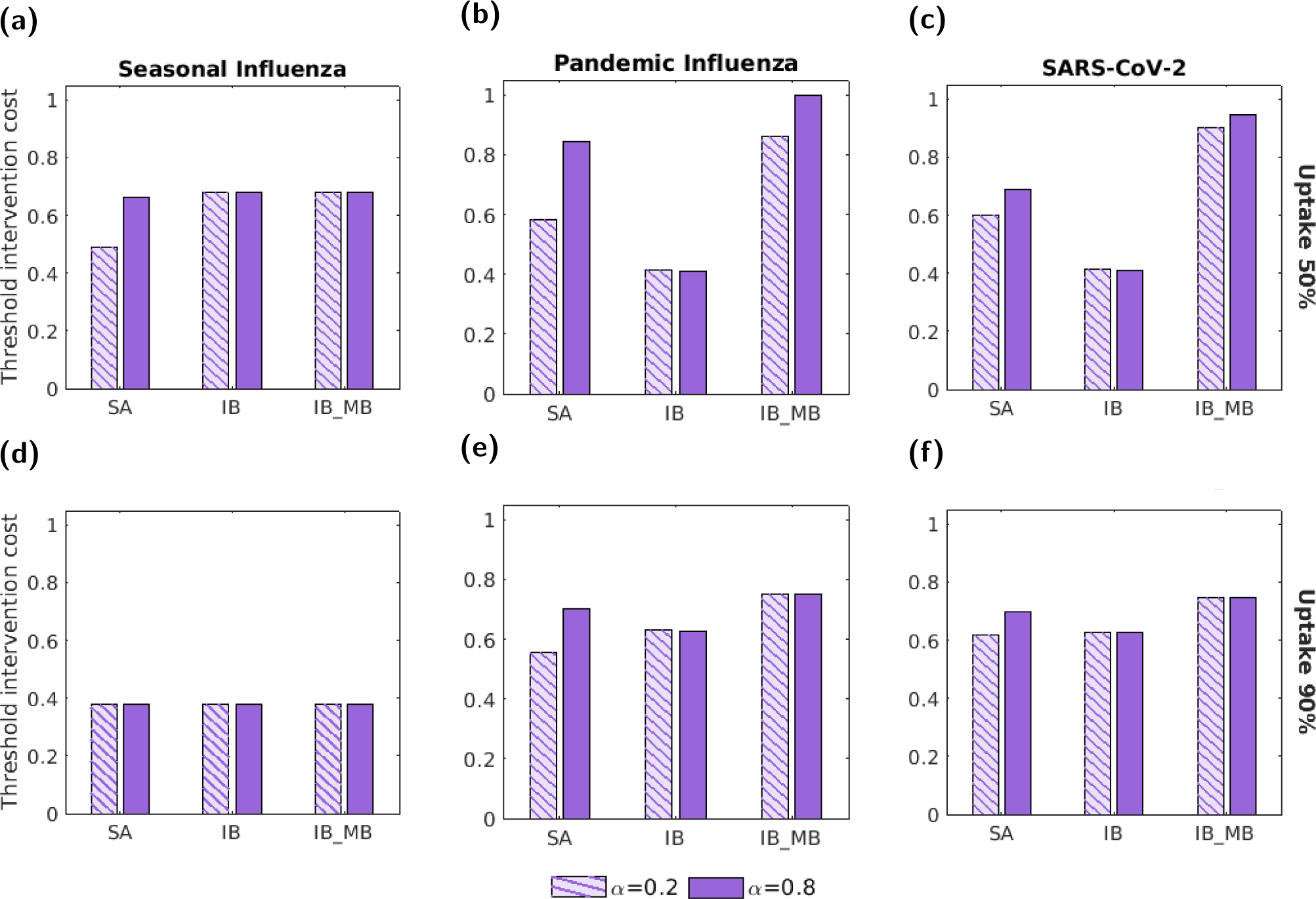
The threshold unit intervention costs for the three interventions and three disease parameterisations with no discounting applied. Analogous to Fig. 7 in the main manuscript, but using a discount rate of 0% (whereas in the main text we used a discount rate of 3.5%). In all panels, we normalised threshold unit intervention costs by the highest absolute threshold unit intervention cost attained across the range of tested intervention uptake values. The three groups of bars correspond to three interventions: a symptom attenuating vaccine(SA), an infection blocking vaccine (IB) and an infection blocking vaccine that only admits mild breakthrough infections (IB MB). The two bars in each group correspond to symptom propagation strengths of *α* = 0.2 (left bar, hatched lines) and *α* = 0.8 (right bar, solid fill). The two rows correspond to two vaccine uptake levels: **(a-c)** 50%; **(d-f)** 90%. Columns correspond to differing disease parameterisations: **(a,d)** seasonal influenza; **(b,e)** pandemic influenza; **(c,f)** SARS-CoV-2. Vaccine efficacies were fixed at 70% and all other parameters were as given in Table 1, with *ν* chosen to fix the proportion of cases that were severe equal to 0.8. The results shown here are qualitatively similar to those obtained when using 3.5% discounting.

**Figure S15:**
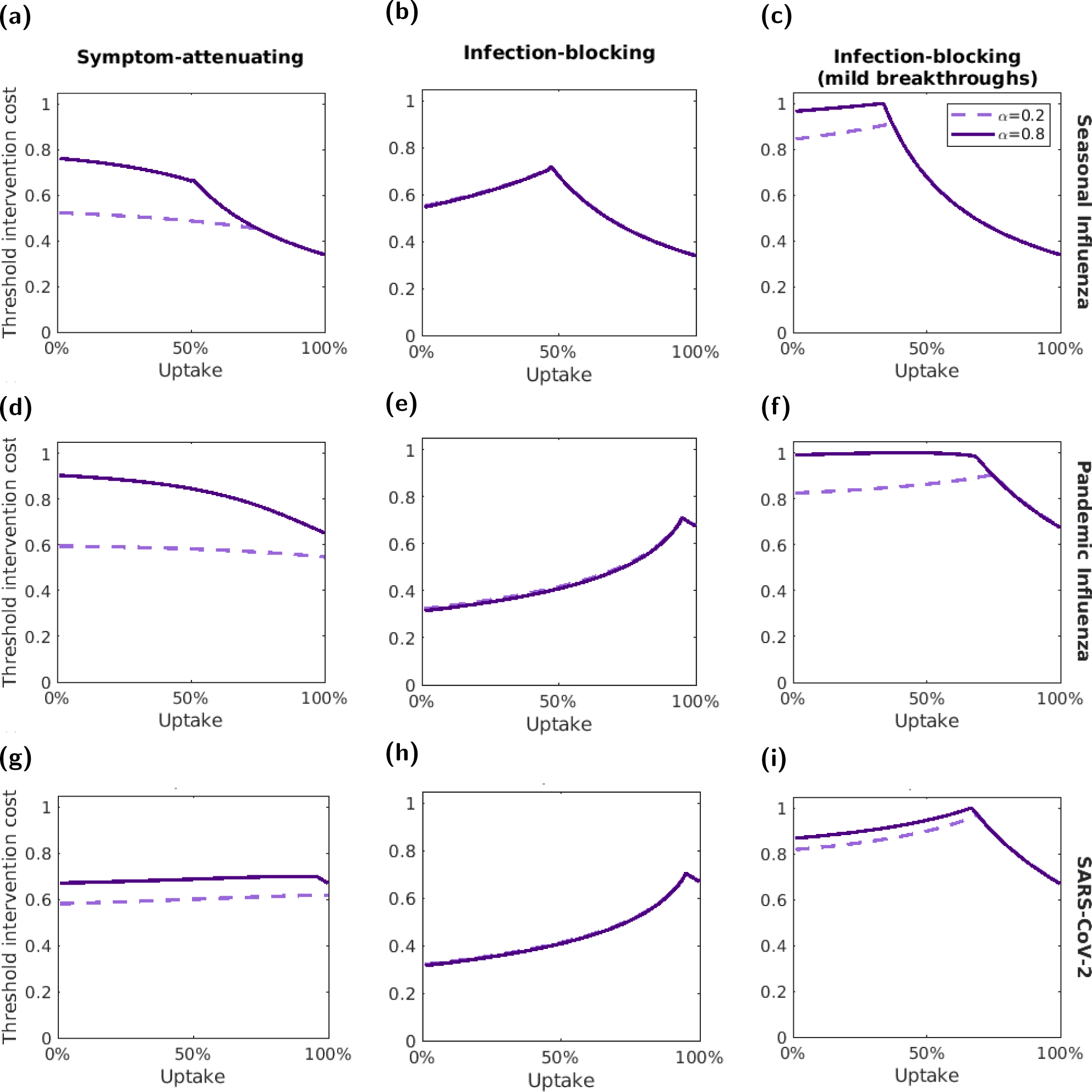
Variation of the threshold unit intervention cost with intervention uptake, with no discounting being applied. Analogous to Fig. 8 in the main manuscript, but using a discount rate of 0% (whereas in the main text we used a discount rate of 3.5%). We normalised threshold unit intervention costs for each disease parameterisation; the normalisation constant was the highest absolute threshold unit intervention cost attained for the respective disease parameterisation across the range of tested intervention uptake values. The three rows correspond to different disease parameterisations: **(a-c)** seasonal influenza; **(d-f)** pandemic influenza; **(g-i)** SARS-CoV-2. The three columns correspond to three interventions: **(a,d,g)** a symptom-attenuating vaccine (SA), **(b,e,h)** an infection-blocking vaccine (IB) and **(c,f,i)** an infection-blocking vaccine that only admits mild breakthrough infections (IB MB). The two lines correspond to two symptom propagation strengths; the dashed, light purple line corresponds to *α* = 0.2 and the solid, dark purple line corresponds to *α* = 0.8. We fixed vaccine efficacies at 70% and all other parameters values were as given in Table 1, with *ν* chosen to fix the proportion of cases that were severe equal to 0.8.

## S7 Sensitivity to intervention efficacy

In the main text we considered vaccination interventions with an efficacy of 70%. Here we performed the same analysis with two alternative vaccine efficacy values: 50% and 90%.

Overall, we found qualitatively similar results for all three vaccine efficacy values. As expected, generally fewer infections were prevented when the efficacy was 50% (Fig. S16) and more infections were prevented when the efficacy was 90% (Fig. S20). However, the patterns between intervention types and the two *α* values were consistent across the three levels of vaccine efficacy.

We did reveal that the relative effectiveness of a symptom attenuating and infection blocking intervention varied with the efficacy value. At 50% efficacy, a symptom-attenuating vaccine was more effective for a larger proportion of the parameter space, as indicated by the increase in areas shaded in red (Fig. S17). At 90% efficacy, an infection-blocking vaccine was more effective for a larger proportion of the parameter space, as indicated by the increase in areas shaded in blue (Fig. S21).

Patterns in threshold unit intervention cost were similar across the three efficacy levels, although the relative cost between the three intervention types varied. At 50% efficacy, the infection blocking vaccine was generally much less cost-effective than the other two types (Figs. S18 and S19), whereas at 90% efficacy the threshold unit intervention cost was more consistent across intervention types (Figs. S22 and S23).

### S7.1 50% efficacy

**Figure S16:**
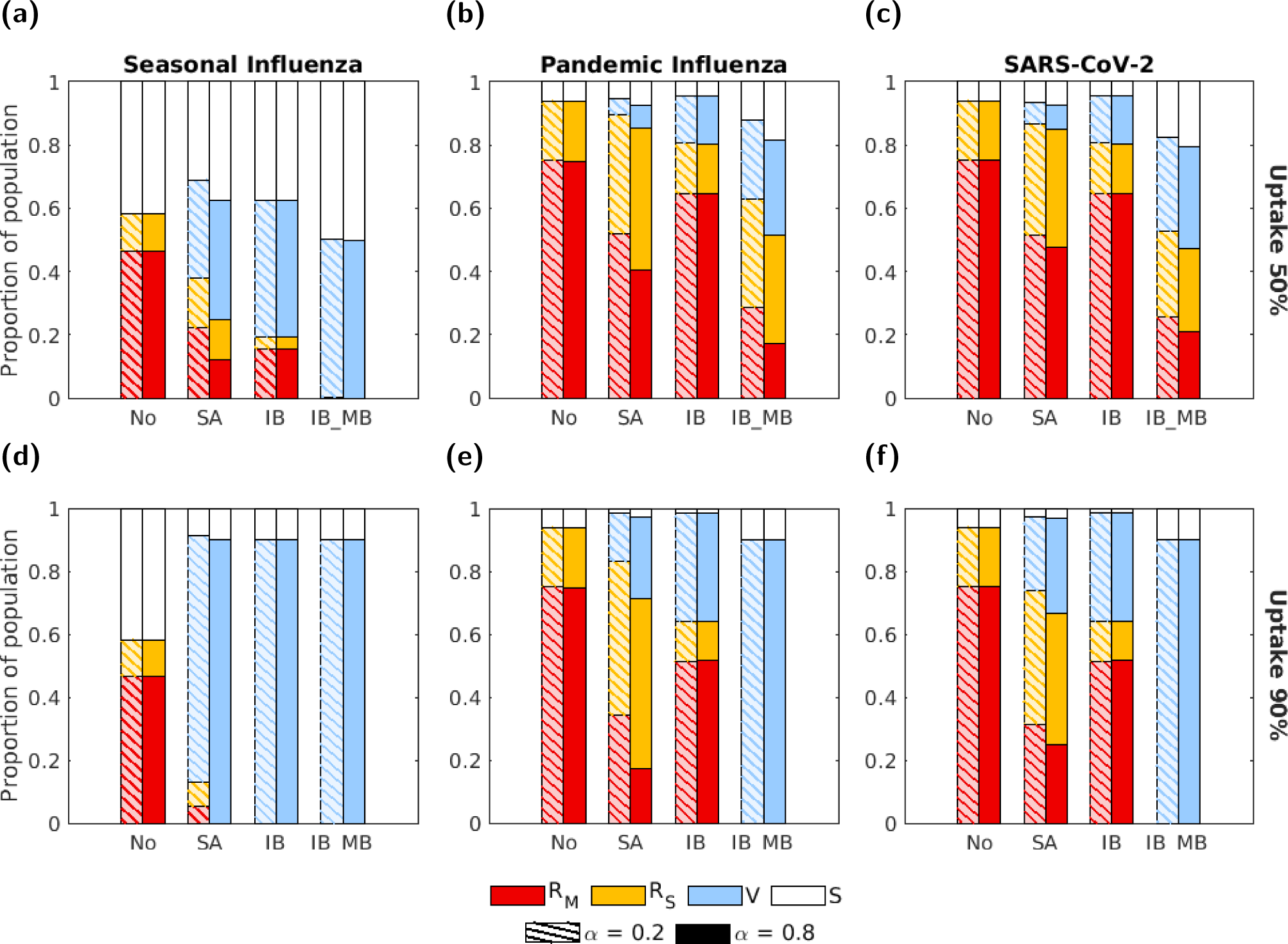
The proportion of the population in each disease state at the end of the outbreak for the four intervention scenarios (with an intervention efficacy of 50%) and three disease parameterisations. The four groups of bars correspond to four intervention scenarios: no intervention (No), a symptom attenuating vaccine (SA), an infection blocking vaccine (IB) and an infection blocking vaccine which only admits mild breakthrough infections (IB MB). The two bars in each group correspond to symptom propagation strengths of *α* = 0.2 (left bar, hatched lines) and *α* = 0.8 (right bar, solid fill). Bar shading corresponds to disease status: red - recovered from severe infection (*R_S_*); yellow - recovered from mild infection (*R_M_*); blue - susceptible and vaccinated (*V*); white - susceptible and not vaccinated (*S*). The two rows correspond to two vaccine uptake levels: **(a-c)** 50%; **(d-f)** 90%. Columns correspond to differing disease parameterisations: **(a,d)** seasonal influenza; **(b,e)** pandemic influenza; **(c,f)** SARS-CoV-2. We fixed vaccination efficacies at 50% and all other parameters were as given in Table 1, with *ν* chosen to fix the proportion of cases that were severe equal to 0.8.

**Figure S17:**
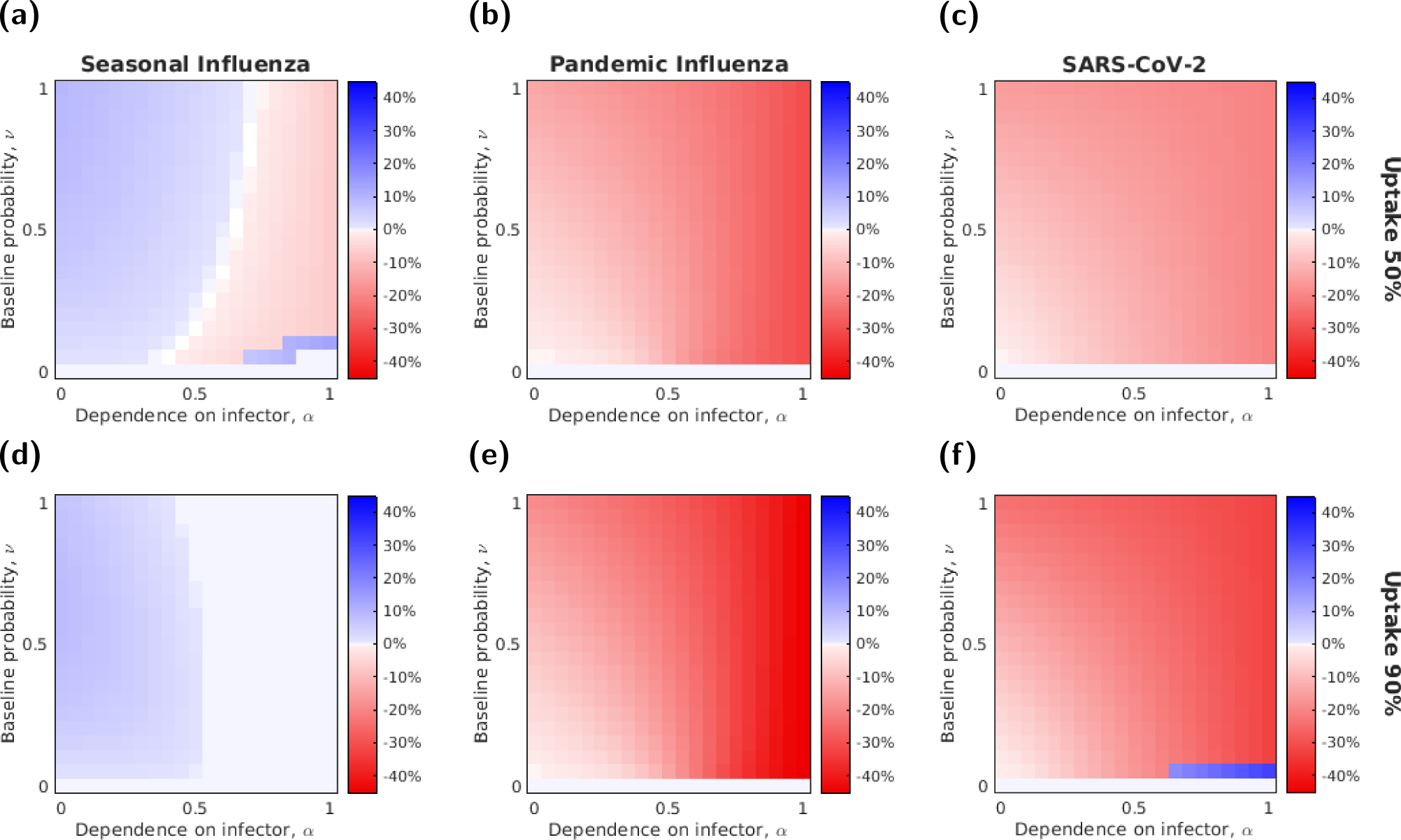
The relative effectiveness of a symptom attenuating and infection blocking vaccine with a fixed efficacy (50%) varies with *α* and *ν*. Each row corresponds to one of the two uptake levels: **(a-c)** 50%; **(d-f)** 90%. Each column corresponds to a different disease parameterisation: **(a,d)** seasonal influenza; **(b,e)** pandemic influenza; **(c,f)** SARS-CoV-2. Cell shading denotes (for that combination of *α*-*ν* value) the difference in the proportion of the population severely infected between when a symptom-attenuating intervention was used and when an infection-blocking intervention was used. The blue shaded cells shows parameter combinations where the infection blocking intervention was more effective at preventing infections, whilst the red shaded cells shows parameter combinations where symptom attenuation was more effective.

**Figure S18:**
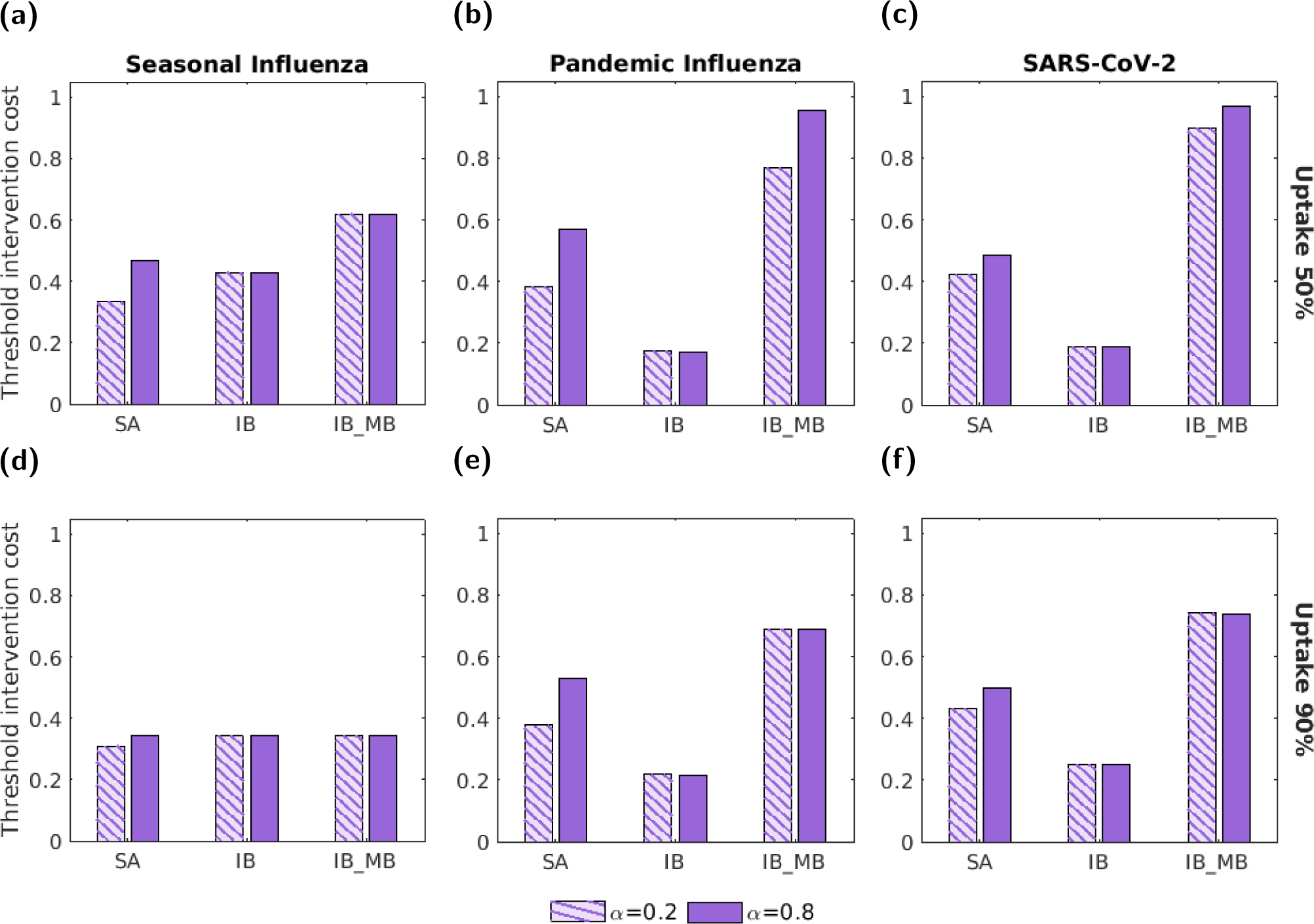
The threshold unit intervention cost for the three vaccine interventions and each disease parameterisation with a vaccine efficacy of 50%. In all panels, we normalised threshold unit intervention cost by the highest absolute threshold unit intervention cost attained across the range of tested intervention uptake values. The three groups of bars correspond to three interventions: a symptom attenuating vaccine (SA), an infection blocking vaccine (IB) and an infection blocking vaccine that only admits mild breakthrough infections (IB MB). The two bars in each group correspond to symptom propagation strengths of *α* = 0.2 (left bar, hatched lines) and *α* = 0.8 (right bar, solid fill). The two rows correspond to two vaccine uptake levels: **(a-c)** 50%; **(d-f)** 90%. Columns correspond to differing disease parameterisations: **(a,d)** seasonal influenza; **(b,e)** pandemic influenza; **(c,f)** SARS-CoV-2. Vaccine efficacies were fixed at 50% and all other parameters were as given in Table 1 with *ν* chosen to fix the proportion of cases that were severe equal to 0.8.

**Figure S19:**
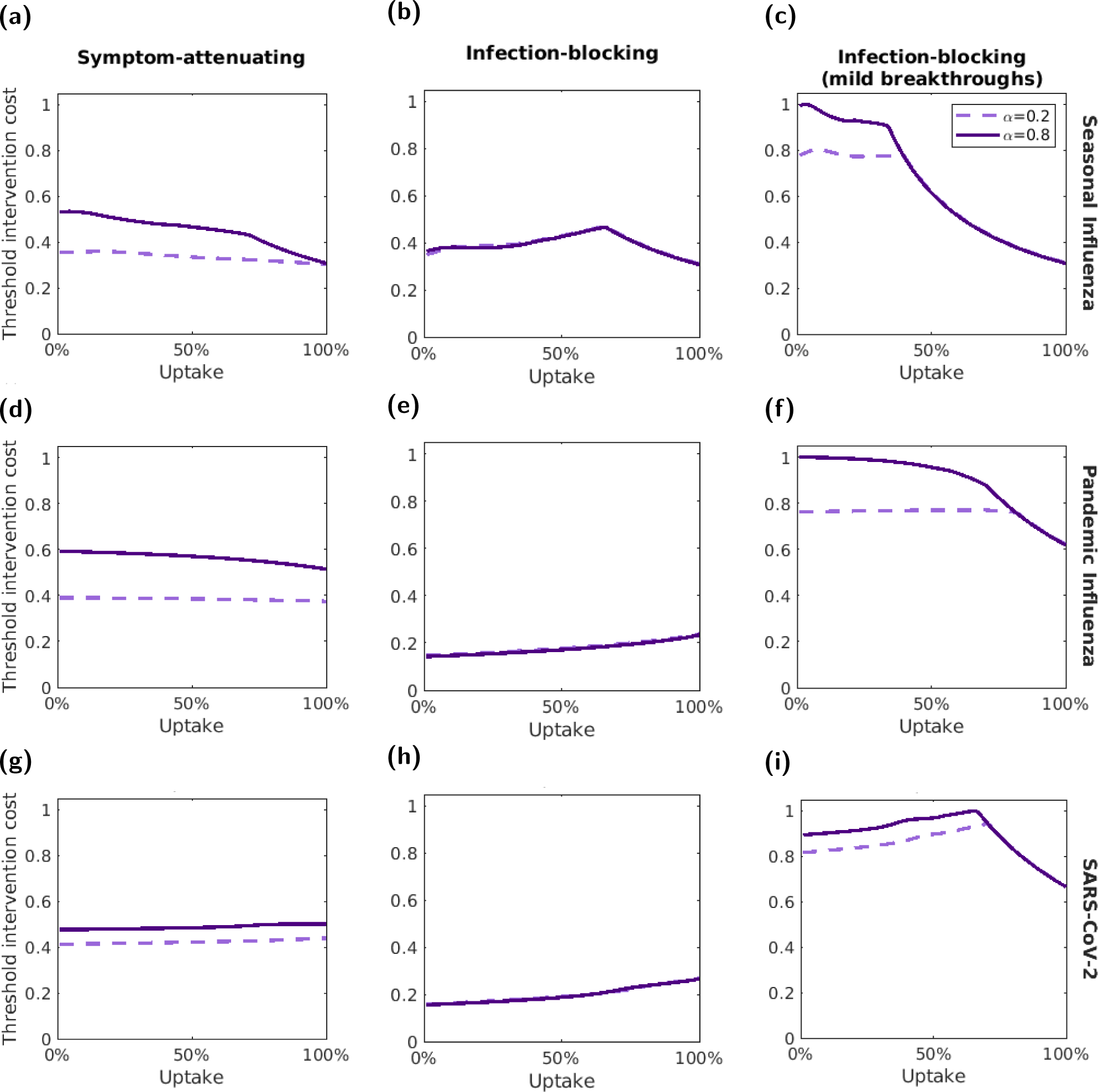
The threshold unit intervention cost varies with vaccine uptake for a vaccine efficacy of 50%. We normalised threshold unit intervention costs for each disease parameterisation; the normalisation constant was the highest absolute threshold unit intervention cost attained for the respective disease parameterisation across the range of tested uptake values. The three rows correspond to the three different disease parameterisations: **(a-c)** seasonal influenza; **(d-f)** pandemic influenza; **(g-i)** SARS-CoV-2. The three columns correspond to three interventions: **(a,d,g)** a symptom-attenuating vaccine (SA), **(b,e,h)** an infection-blocking vaccine (IB) and **(c,f,i)** an infection-blocking vaccine that only admits mild breakthrough infections (IB MB). The two lines correspond to two symptom propagation strengths; the dashed, light purple line corresponds to *α* = 0.2 and the solid, dark purple line corresponds to *α* = 0.8. We fixed the vaccine efficacies at 50% and all other parameters values were as given in Table 1, with *ν* chosen to fix the proportion of cases that were severe equal to 0.8.

### S7.2 90% efficacy

**Figure S20:**
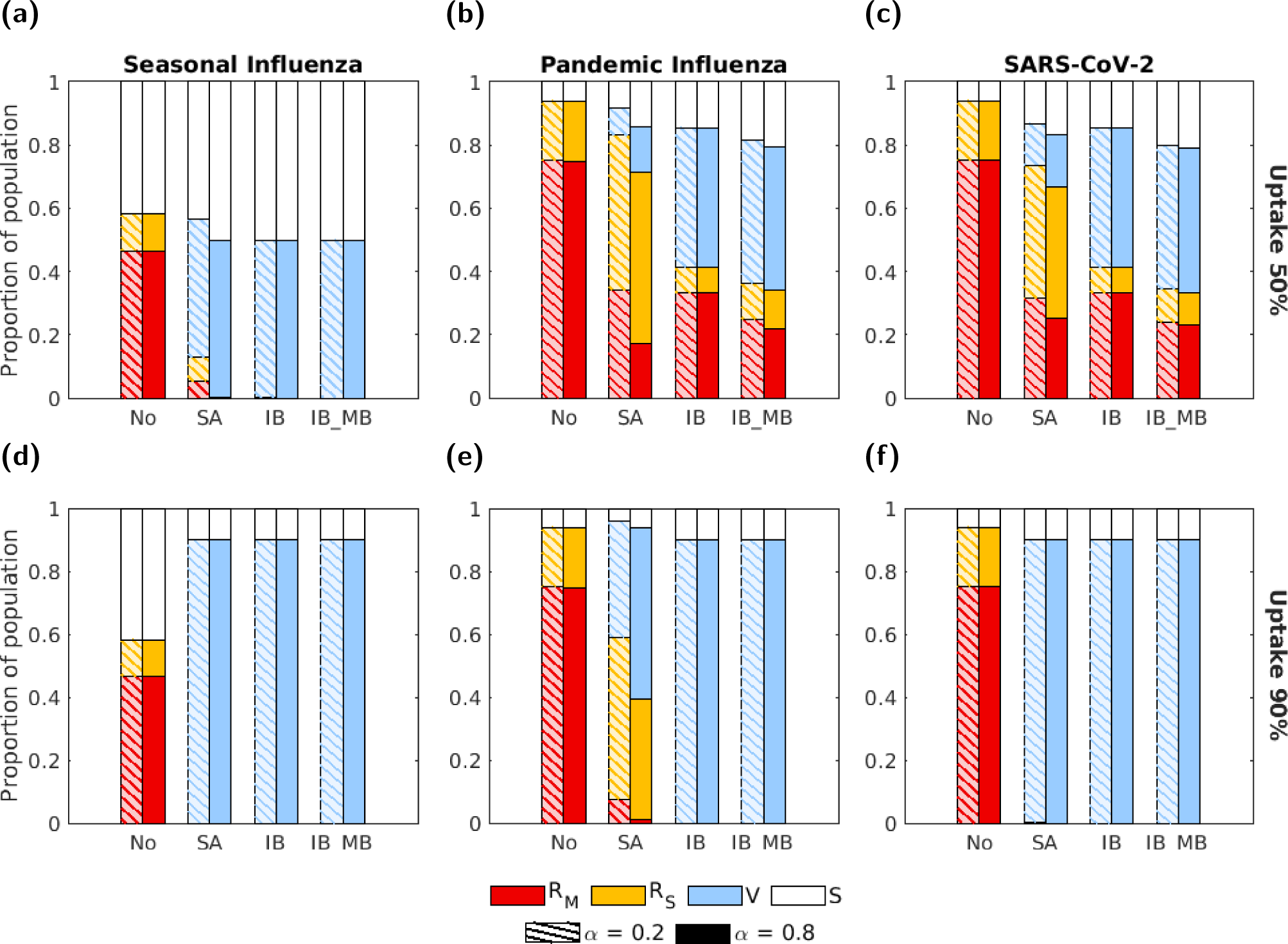
The proportion of the population in each disease state at the end of the outbreak for the four intervention scenarios (with vaccine efficacies of 90%) and three disease parameterisations. The four groups of bars correspond to four intervention scenarios: no intervention (No), a symptom attenuating vaccine (SA), an infection blocking vaccine (IB) and an infection blocking vaccine that only admits mild breakthrough infections (IB MB). The two bars in each group correspond to symptom propagation strengths of *α* = 0.2 (left bar, hatched lines) and *α* = 0.8 (right bar, solid fill). Bar shading corresponds to disease status: red - recovered from severe infection (*R_S_*); yellow - recovered from mild infection (*R_M_*); blue - susceptible and vaccinated (*V*); white - susceptible and not vaccinated (*S*). The two rows correspond to two vaccine uptake levels: **(a-c)** 50%; **(d-f)** 90%. Columns correspond to differing disease parameterisations: **(a,d)** seasonal influenza; **(b,e)** pandemic influenza; **(c,f)** SARS-CoV-2. We fixed the vaccine efficacies at 90% and all other parameters were as given in Table 1, with *ν* chosen to fix the proportion of cases that were severe equal to 0.8.

**Figure S21:**
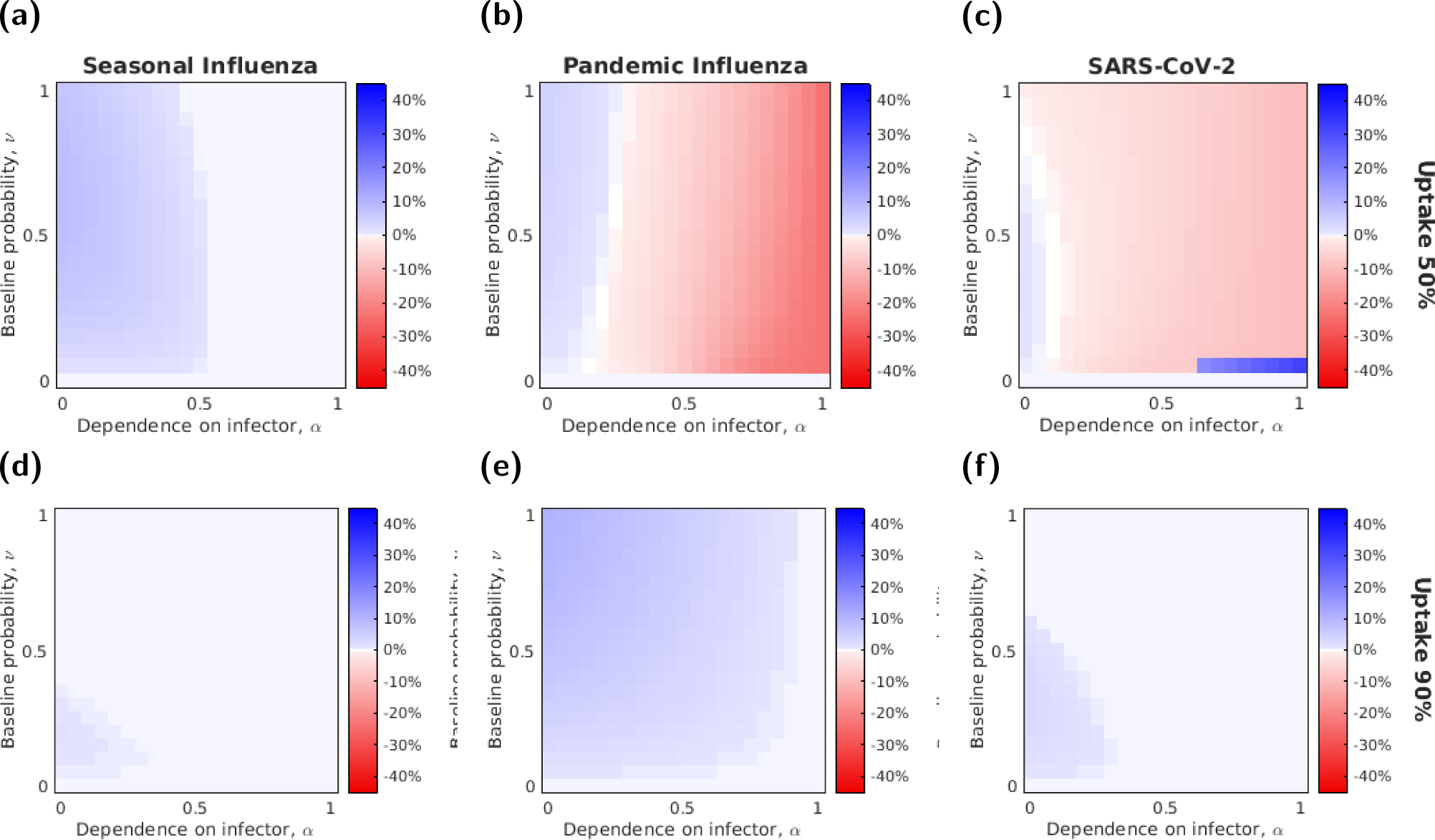
The relative effectiveness of a symptom attenuating and infection blocking intervention with a fixed efficacy (90%) varies with *α* and *ν*. Each row corresponds to one of the two uptake levels: **(a-c)** 50%; **(d-f)** 90%. Each column corresponds to a different disease parameterisation: **(a,d)** seasonal influenza; **(b,e)** pandemic influenza; **(c,f)** SARS-CoV-2. Cell shading denotes (for that combination of *α*-*ν* value) the difference in the proportion of the population severely infected between when a symptom-attenuating intervention was used and when an infection-blocking intervention was used. The blue shaded cells shows parameter combinations where the infection blocking intervention was more effective at preventing infections, whilst the red shaded cells shows parameter combinations where symptom attenuation was more effective.

**Figure S22:**
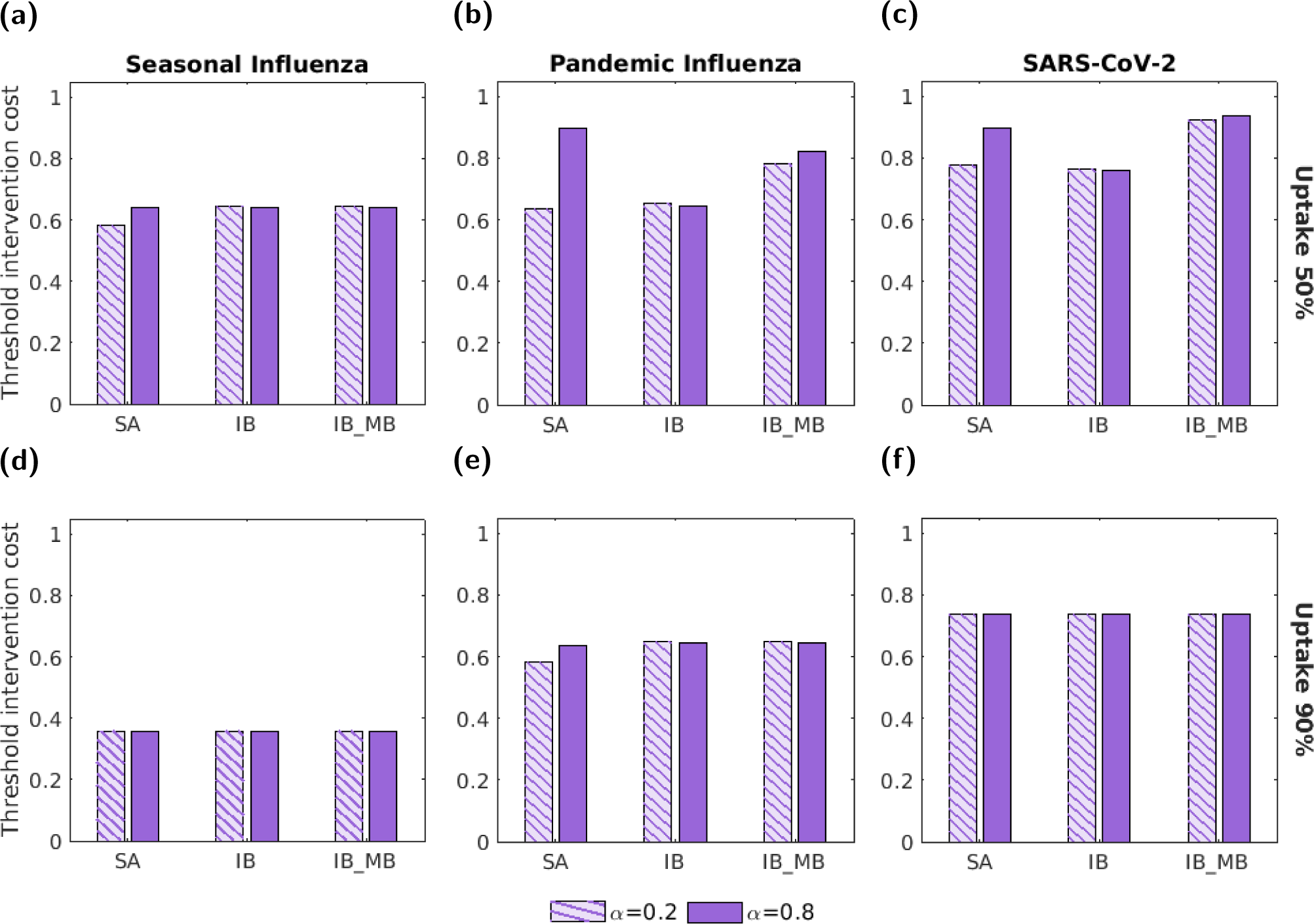
The threshold unit intervention cost for the three vaccine interventions and each disease parameterisation with vaccine efficacies of 90%. In all panels, we normalised threshold unit intervention cost by the highest absolute threshold unit intervention cost attained across the range of tested intervention uptake values. The three groups of bars correspond to three interventions: a symptom attenuating vaccine (SA), an infection blocking vaccine (IB) and an infection blocking vaccine which only admits mild breakthrough infections (IB MB). The two bars in each group correspond to symptom propagation strengths of *α* = 0.2 (left bar, hatched lines) and *α* = 0.8 (right bar, solid fill). The two rows correspond to two vaccine uptake levels: **(a-c)** 50%; **(d-f)** 90%. Columns correspond to differing disease parameterisations: **(a,d)** seasonal influenza; **(b,e)** pandemic influenza; **(c,f)** SARS-CoV-2. Vaccine efficacies were fixed at 90% and all other parameters were as given in Table 1 with *ν* chosen to fix the proportion of cases that were severe equal to 0.8.

**Figure S23:**
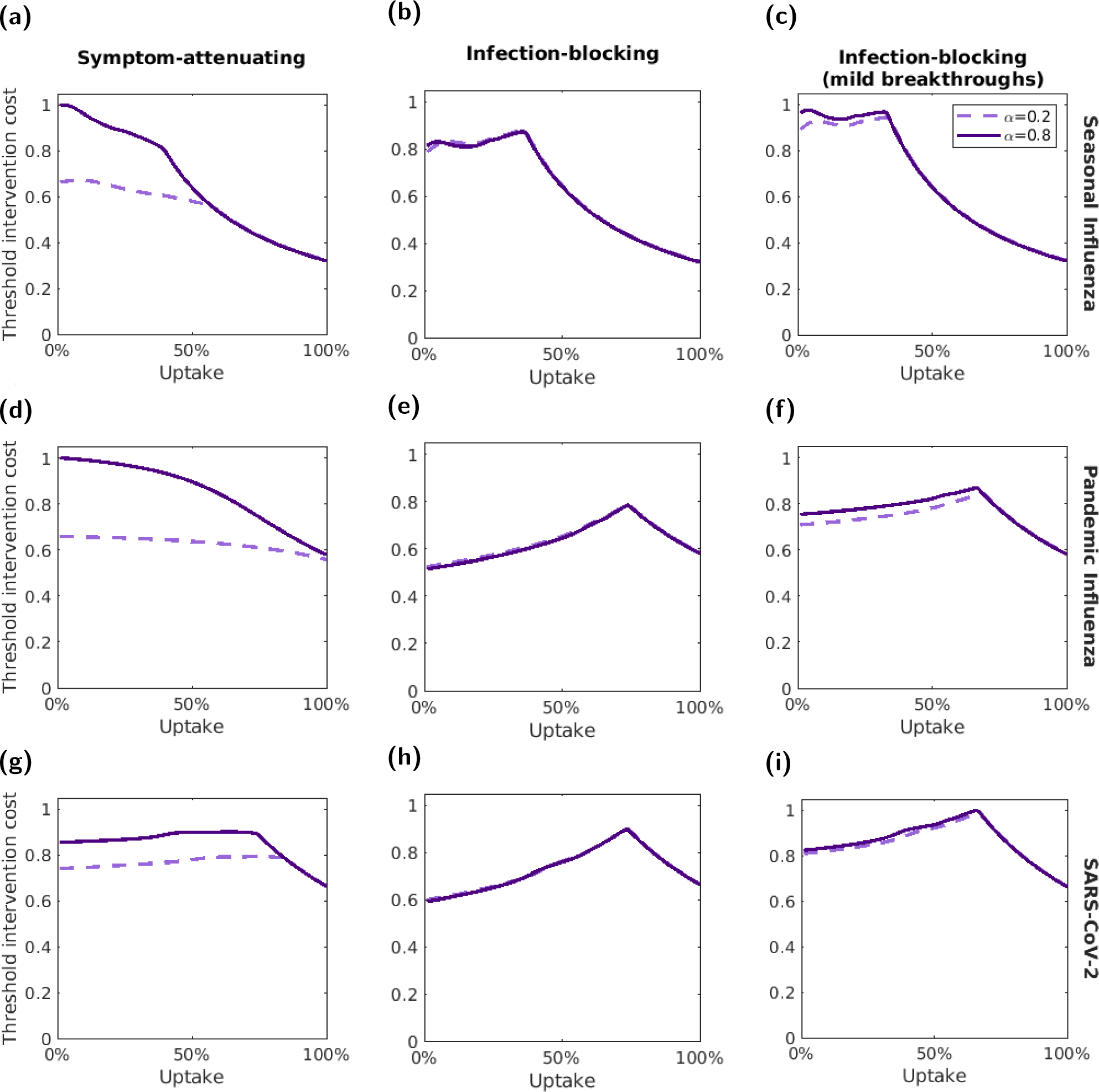
Variation in the threshold unit intervention cost with vaccine uptake for vaccine efficacies of 90%. We normalised threshold unit intervention costs for each disease parameterisation; the normalisation constant was the highest absolute threshold unit intervention cost attained for the respective disease parameterisation across the range of tested vaccine uptake values. The three rows correspond to the three different disease parameterisations: **(a-c)** seasonal influenza; **(d-f)** pandemic influenza; **(g-i)** SARS-CoV-2. The three columns correspond to three interventions: **(a,d,g)** a symptom-attenuating vaccine (SA), **(b,e,h)** an infection-blocking vaccine (IB) and **(c,f,i)** an infection-blocking vaccine that only admits mild breakthrough infections (IB MB). The two lines correspond to two symptom propagation strengths; the dashed, light purple line corresponds to *α* = 0.2 and the solid, dark purple line corresponds to *α* = 0.8. We fixed the vaccine efficacies at 90% and all other parameters values were as given in Table 1, with *ν* chosen to fix the proportion of cases that were severe equal to 0.8.

## S8 Sensitivity in *α* − ν space

### S8.1 Relative threshold unit intervention cost

**Figure S24:**
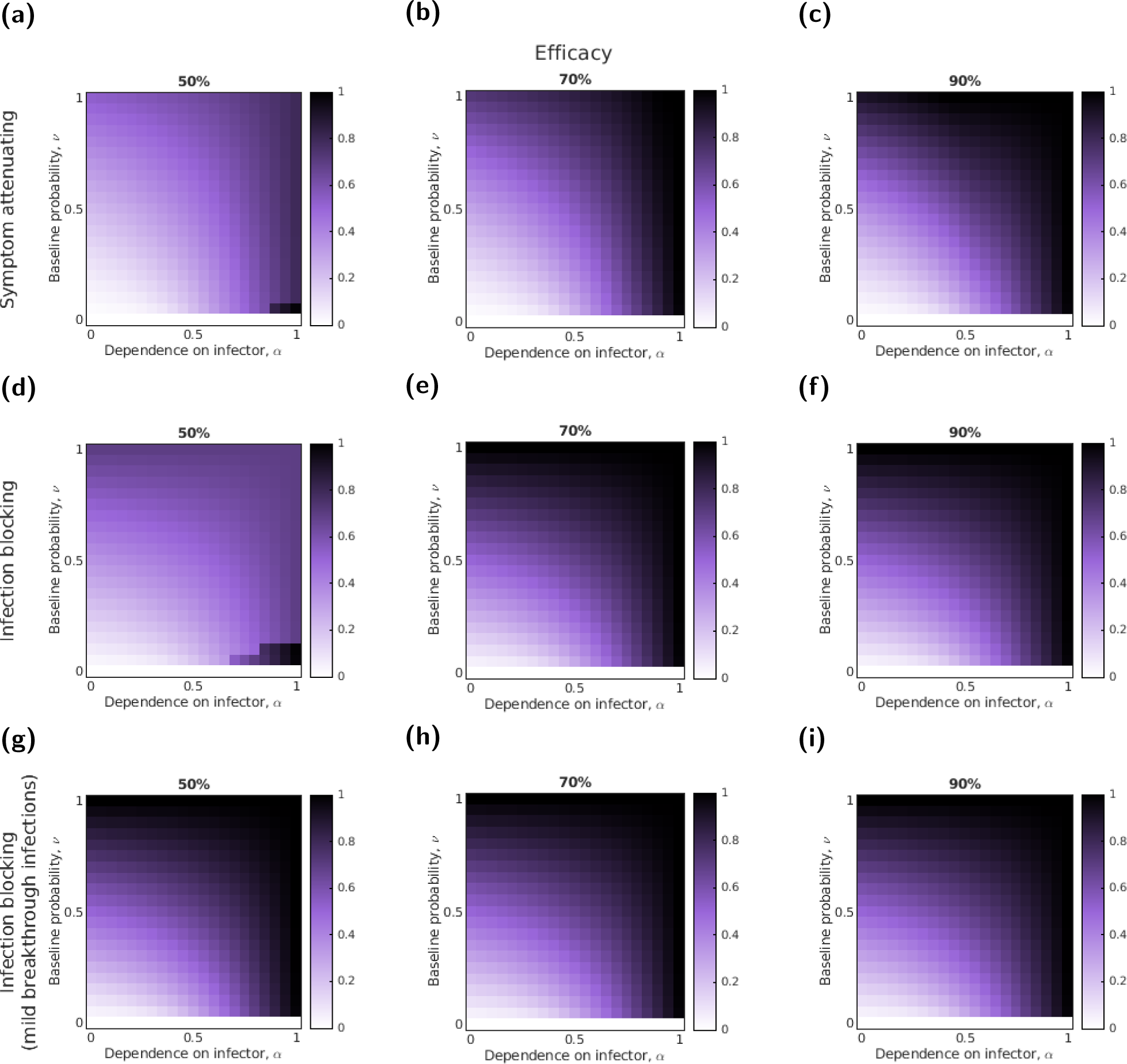
Variation in the threshold unit intervention cost with *α*, *ν* and the efficacy for three intervention types, seasonal influenza-like parameters and a fixed intervention uptake (50%). Cell shading denotes the relative threshold unit intervention cost (normalised by the highest threshold unit intervention cost value). Darker shading corresponds to larger values. The assessed interventions had the following action: **(a-c)** symptom attenuating; **(d-f)** infection blocking; **(g-i)** infection blocking that only admits mild breakthrough infections. We performed evaluations for three efficacies per intervention: **(a,d,g)** 50%; **(b,e,h)** 70%; **(c,f,i)** 90%.

**Figure S25:**
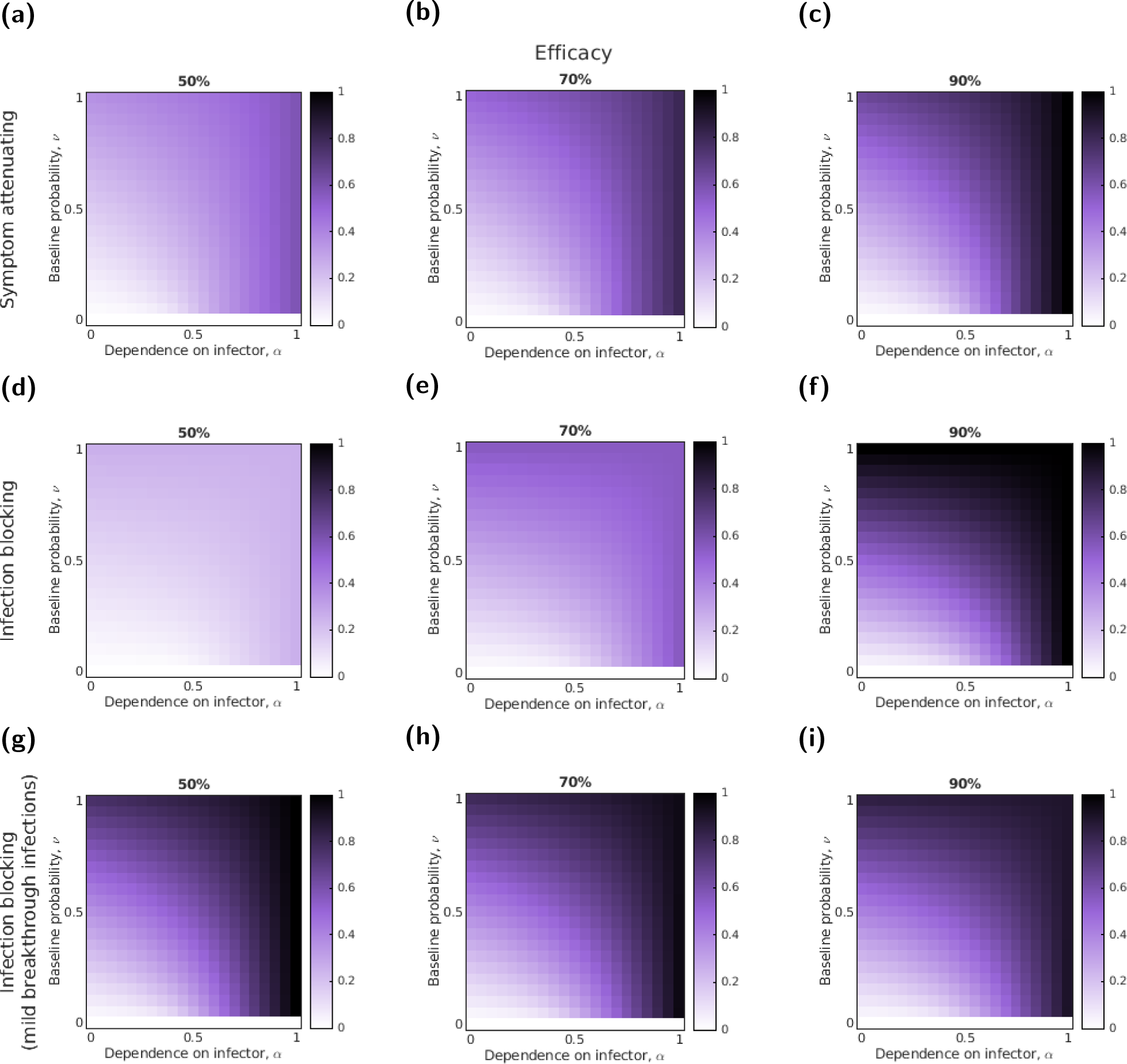
Variation in the threshold unit intervention cost with *α*, *ν* and the efficacy for three intervention types, pandemic influenza-like parameters and a fixed uptake (50%). Cell shading denotes the relative threshold unit intervention cost (normalised by the highest threshold unit intervention cost value). Darker shading corresponds to larger values. The assessed interventions had the following action: **(a-c)** symptom attenuating; **(d-f)** infection blocking; **(g-i)** infection blocking intervention that only admits mild breakthrough infections. We performed evaluations for three efficacies per intervention: **(a,d,g)** 50%; **(b,e,h)** 70%; **(c,f,i)** 90%.

**Figure S26:**
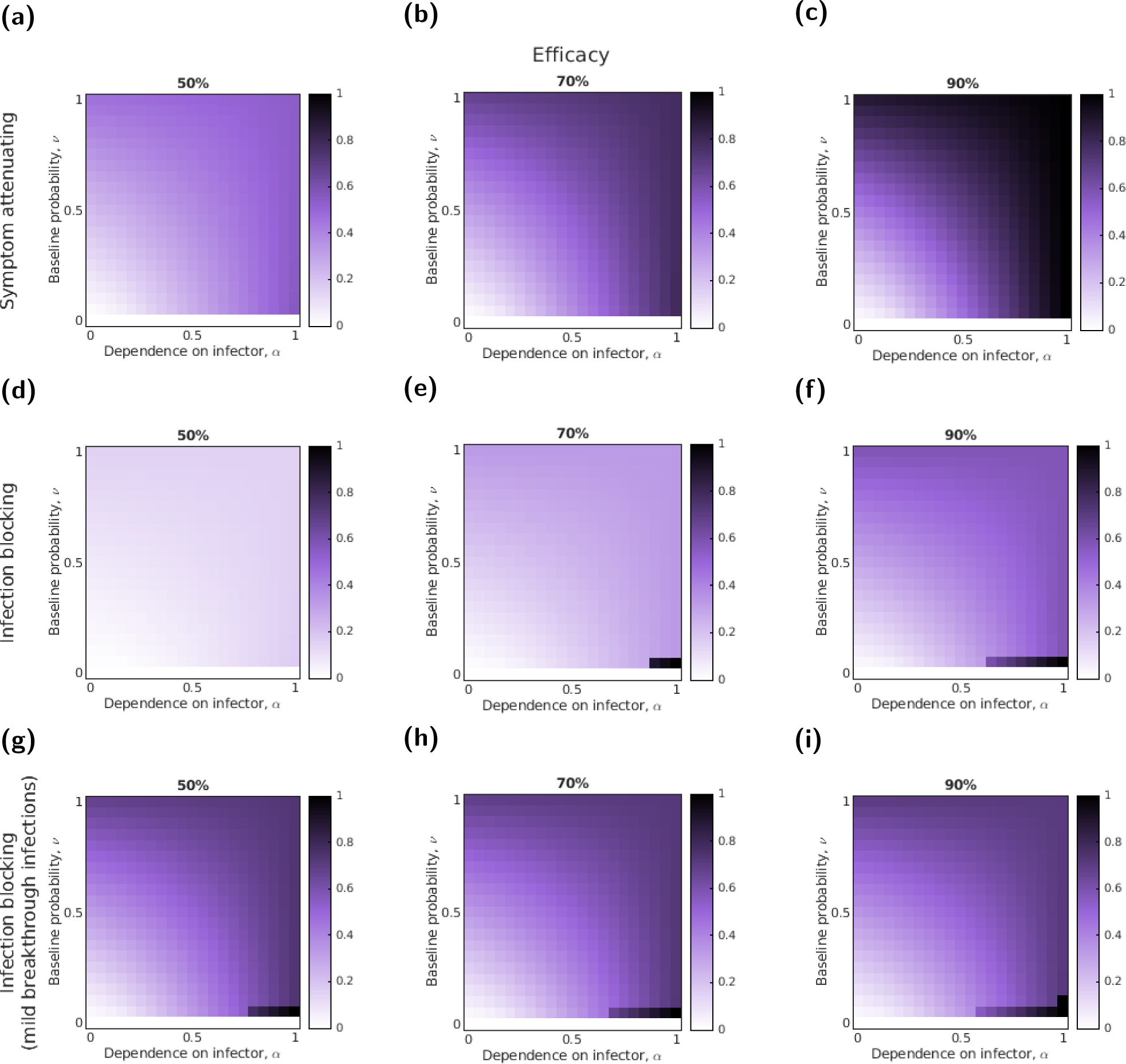
Variation in the threshold unit intervention cost with *α*, *ν* and the efficacy for three intervention types, SARS-CoV-2-like parameters and a fixed uptake (50%). Cell shading denotes the relative threshold unit intervention cost (normalised by the highest threshold unit intervention cost value). Darker shading corresponds to larger values. The assessed interventions had the following action: **(a-c)** symptom attenuating; **(d-f)** infection blocking; **(g-i)** infection blocking that only admits mild breakthrough infections. We performed evaluations for three efficacies per intervention: **(a,d,g)** 50%; **(b,e,h)** 70%; **(c,f,i)** 90%.

### S8.2 Most cost-effective intervention uptake

**Figure S27:**
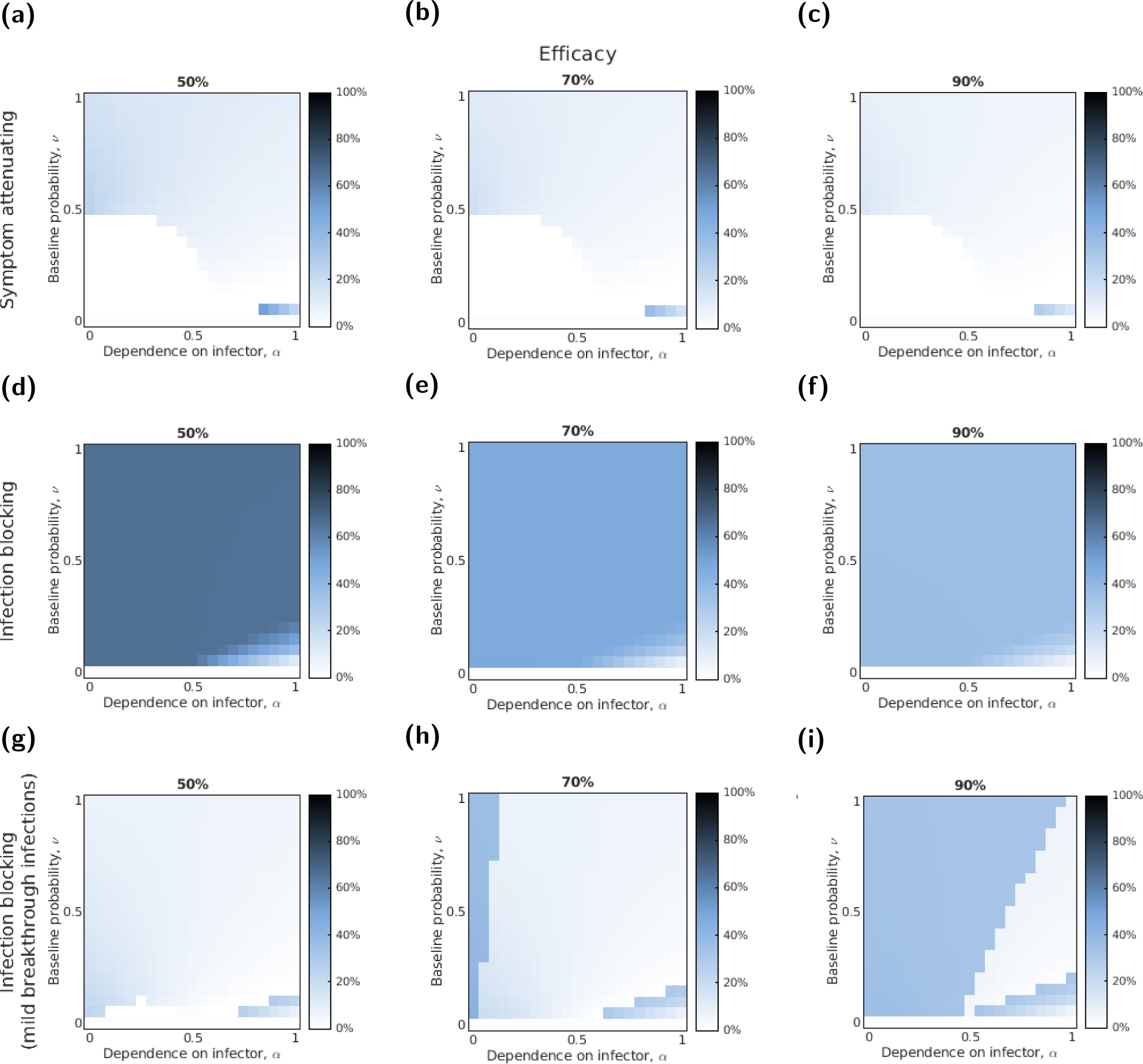
Variation in the most cost-effective vaccine uptake with *α*, *ν* and the efficacy for three intervention types and seasonal influenza-like parameters. Cell shading denotes the uptake at which the intervention was most cost-effective, i.e. the uptake at which the threshold unit intervention cost was at a maximum. Darker shading corresponds to larger values. The assessed interventions had the following action: **(a-c)** symptom attenuating; **(d-f)** infection blocking; **(g-i)** infection blocking that only admits mild breakthrough infections. We performed evaluations for three efficacies per intervention: **(a,d,g)** 50%; **(b,e,h)** 70%; **(c,f,i)** 90%.

**Figure S28:**
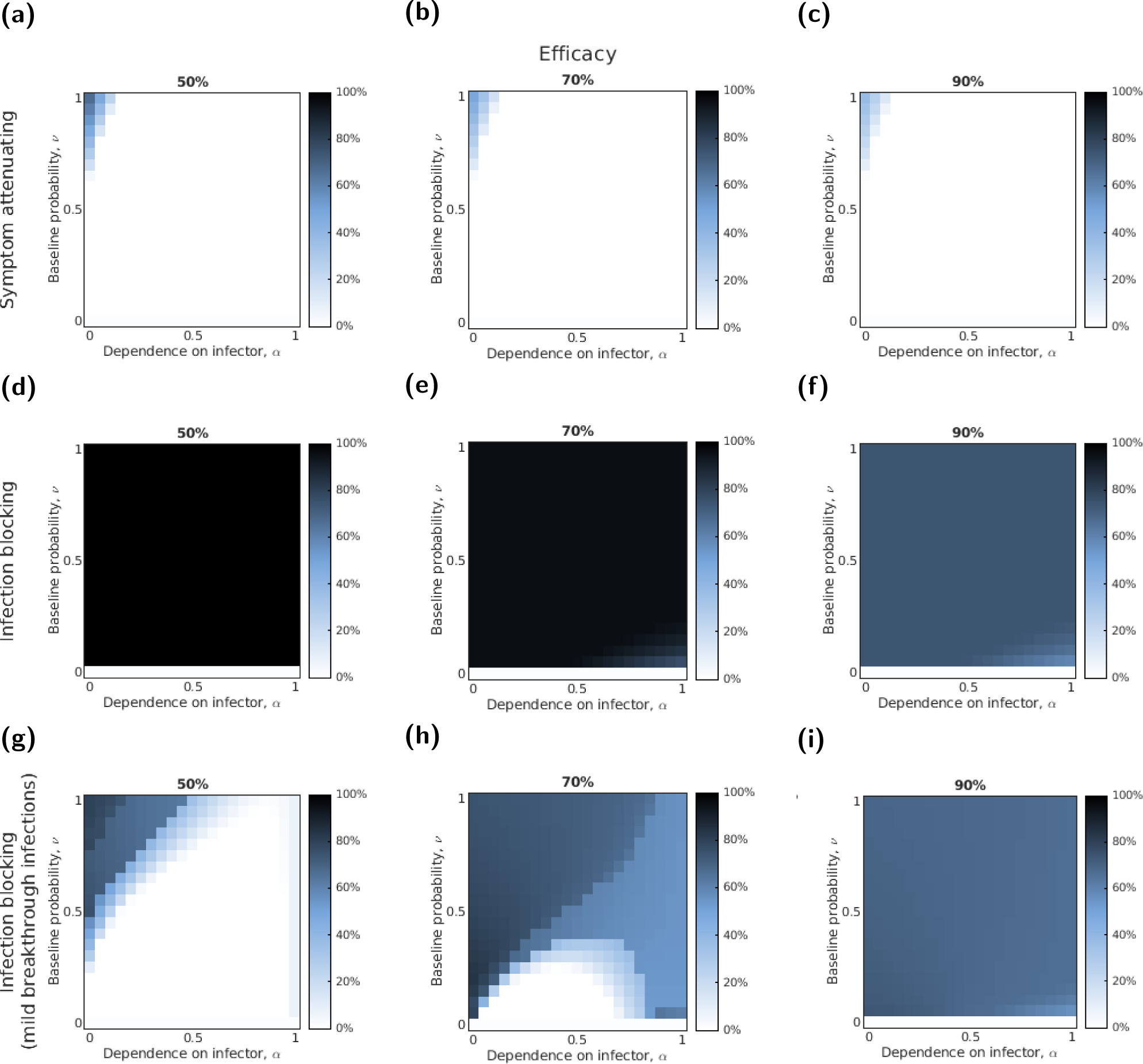
Variation in the most cost-effective vaccine uptake with *α*, *ν* and the efficacy for three intervention types and pandemic influenza-like parameters. Cell shading denotes the uptake at which the intervention was most cost-effective, i.e. the uptake at which the threshold unit intervention cost was at a maximum. Darker shading corresponds to larger values. The assessed interventions had the following action: **(a-c)** symptom attenuating; **(d-f)** infection blocking; **(g-i)** infection blocking that only admits mild breakthrough infections. We performed evaluations for three efficacies per intervention: **(a,d,g)** 50%; **(b,e,h)** 70%; **(c,f,i)** 90%.

**Figure S29:**
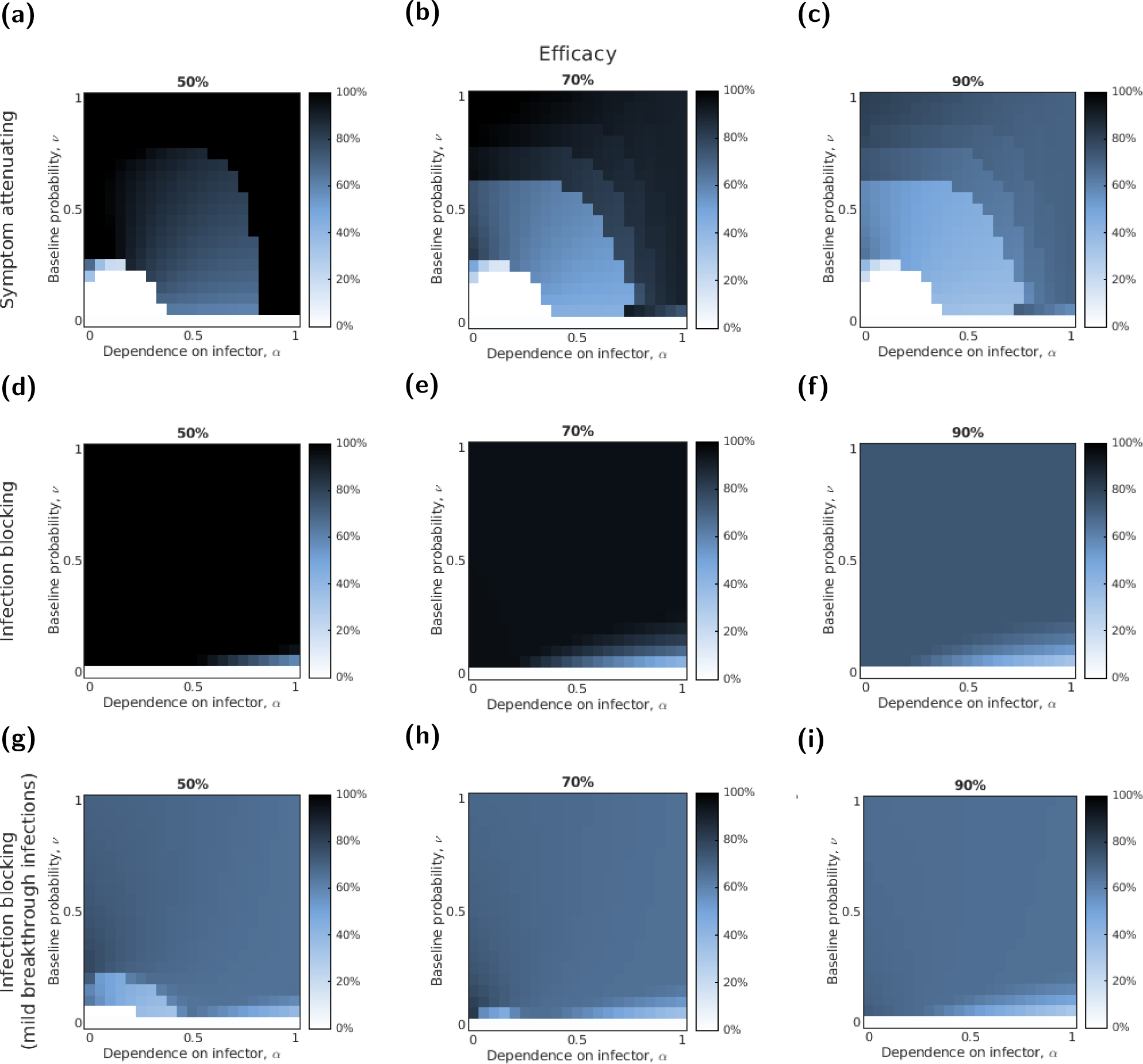
Variation in the most cost-effective vaccine uptake with *α*, *ν* and the efficacy for three intervention types and SARS-CoV-2-like parameters. Cell shading denotes the uptake at which the intervention was most cost-effective, i.e. the uptake at which the threshold unit intervention cost was at a maximum. Darker shading corresponds to larger values. The assessed interventions had the following action: **(a-c)** symptom attenuating; **(d-f)** infection blocking; **(g-i)** infection blocking that only admits mild breakthrough infections. We performed evaluations for three efficacies per intervention: **(a,d,g)** 50%; **(b,e,h)** 70%; **(c,f,i)** 90%.

### S8.3 Implication on threshold unit intervention cost when accounting for symptom propagation

**Figure S30:**
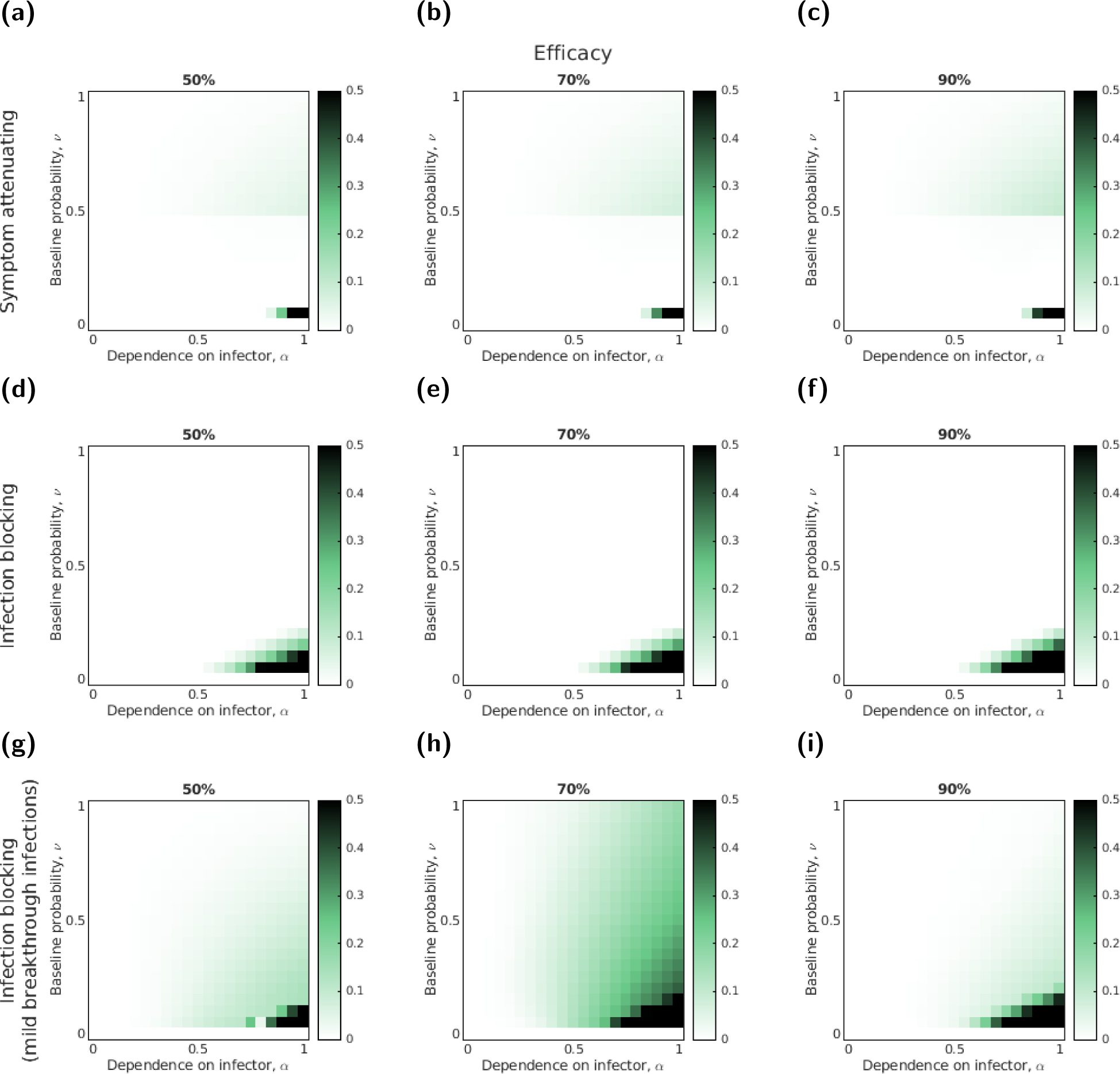
Comparing the threshold unit intervention costs at the most-effective uptake when accounting for symptom propagation to the threshold unit intervention cost at the most cost-effective uptake when there was no symptom propagation (*α* = 0) for three intervention types and seasonal influenza-like parameters. In each panel, darker shading represents a greater disparity in the normalised threshold unit intervention costs between the most cost-effective uptake and the uptake that was most cost-effective when *α* = 0. The assessed interventions had the following action: **(a-c)** symptom-attenuating; **(d-f)** infection-blocking; **(g-i)** infection-blocking that only admits mild breakthrough infections. We performed evaluations for three efficacies per intervention: **(a,d,g)** 50%; **(b,e,h)** 70%; **(c,f,i)** 90%.

**Figure S31:**
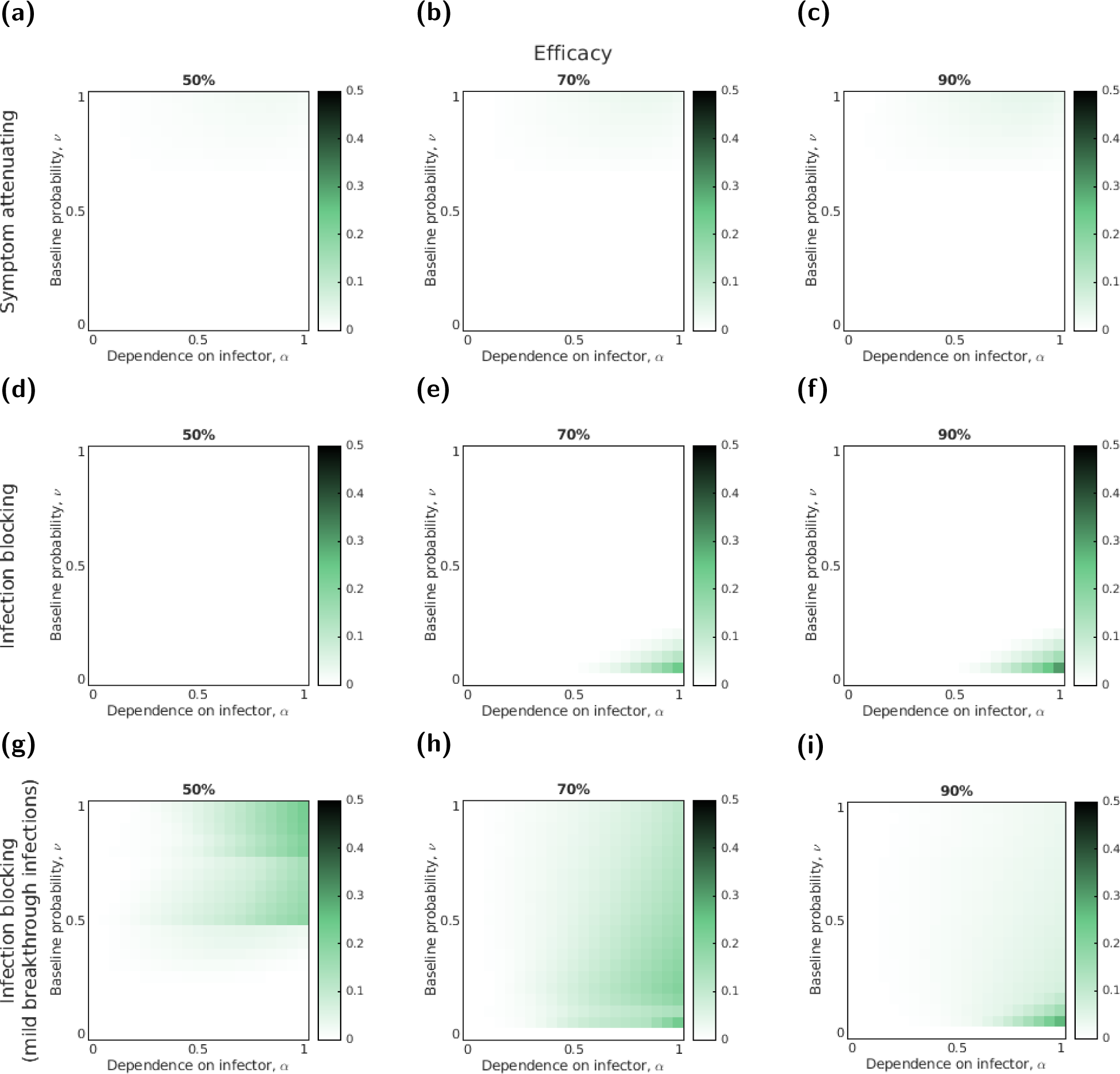
Comparing the threshold unit intervention costs at the most-effective uptake when accounting for symptom propagation to the threshold unit intervention cost at the most cost-effective uptake when there was no symptom propagation (*α* = 0) for three intervention types and pandemic influenza-like parameters. In each panel, darker shading represents a greater disparity in the normalised threshold unit intervention costs between the most cost-effective uptake and the uptake that was most cost-effective when *α* = 0. The assessed interventions had the following action: **(a-c)** symptom-attenuating; **(d-f)** infection-blocking; **(g-i)** infection-blocking that only admits mild breakthrough infections. We performed evaluations for three efficacies per intervention: **(a,d,g)** 50%; **(b,e,h)** 70%; **(c,f,i)** 90%.

**Figure S32:**
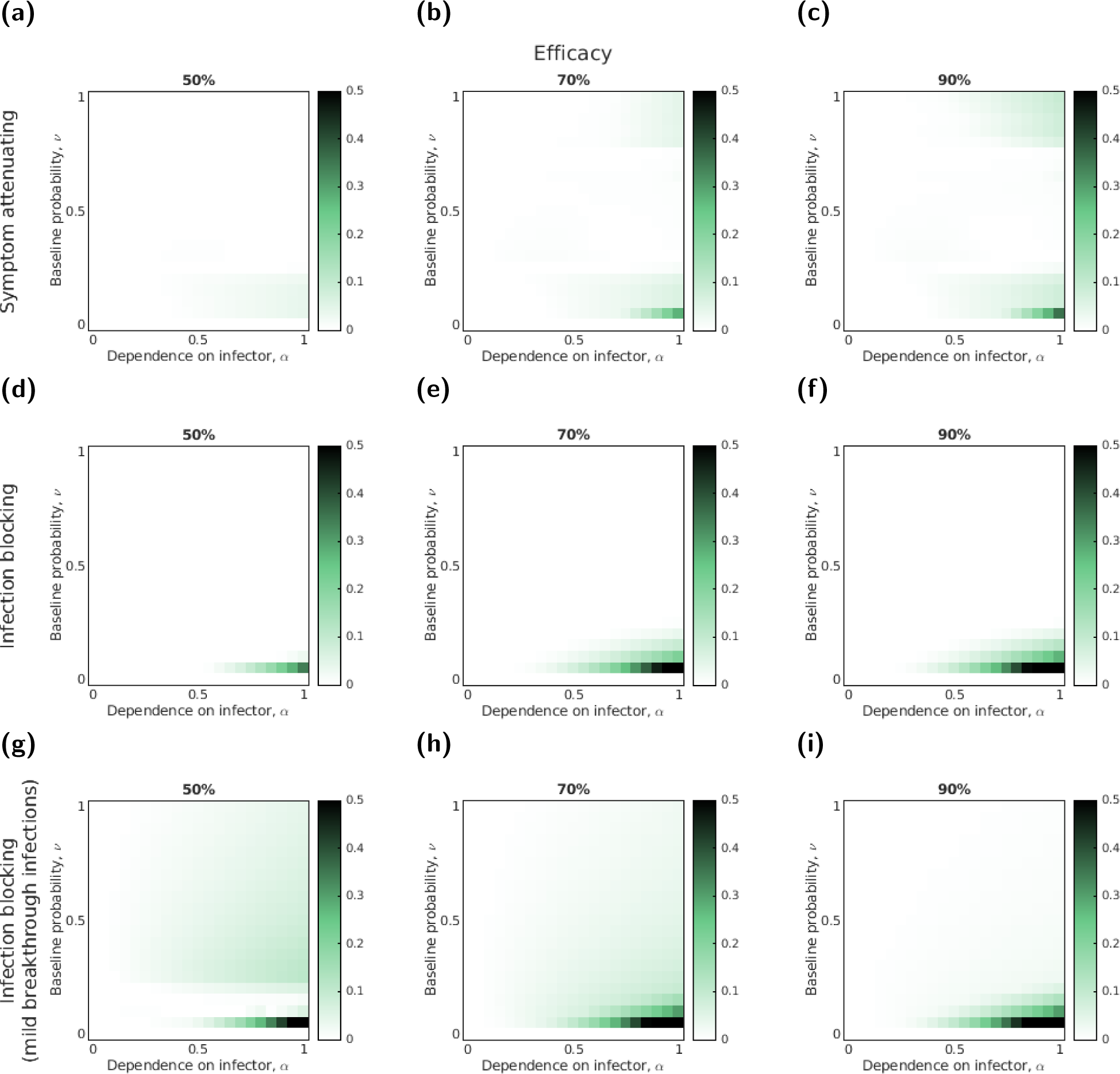
Comparing the threshold unit intervention costs at the most-effective uptake when accounting for symptom propagation to the threshold unit intervention cost at the most cost-effective uptake when there was no symptom propagation (*α* = 0) for three intervention types and SARS-CoV-2-like parameters. In each panel, darker shading represents a greater disparity in the normalised threshold intervention costs between the most cost-effective uptake and the uptake that was most cost-effective when *α* = 0. The assessed interventions had the following action: (a-c) symptom-attenuating; (d-f) infection-blocking; (g-i) infection-blocking that only admits mild breakthrough infections. We performed evaluations for three efficacies per intervention: (a,d,g) 50%; (b,e,h) 70%; (c,f,i) 90%.

## References

[1] Vos T, Lim SS, Abbafati C, Abbas KM, Abbasi M, et al. Global burden of 369 diseases and injuries in 204 countries and territories, 1990–2019: a systematic analysis for the Global Burden of Disease Study 2019. The Lancet 396(10258):1204–1222 (2020). doi:10.1016/S0140-6736(20)30925-9.

[2] Tokars JI, Olsen SJ, Reed C. Seasonal Incidence of Symptomatic Influenza in the United States. Clinical Infectious Diseases 66(10):1511–1518 (2018). doi:10.1093/cid/cix1060.

[3] World Health Organisation. Seasonal Influenza Dashboard (2023). URL https://www.who.int/news-room/fact-sheets/detail/influenza-(seasonal). [Online] (Accessed: 24 January 2024).

[4] Centers for Disease Control and Prevention. 1918 Pandemic (H1N1 virus) | Pandemic Influenza (Flu) (2023). URL https://archive.cdc.gov/#/details?url=https://www.cdc.gov/flu/pandemic-resources/1918-pandemic-h1n1.html. [Online] (Accessed: 24 January 2024).

[5] Dawood FS, Iuliano AD, Reed C, Meltzer MI, Shay DK, et al. Estimated global mortality associated with the first 12 months of 2009 pandemic influenza A H1N1 virus circulation: a modelling study. The Lancet Infectious Diseases 12(9):687–695 (2012). doi:10.1016/S1473-3099(12)70121-4.

[6] World Health Organisation. Coronavirus (COVID-19) Dashboard (2023). URL https://covid19.who.int/. [Online] (Accessed: 24 January 2024).

[7] Gonźalez Ĺopez-Valćarcel B, Vallejo-Torres L. The costs of COVID-19 and the cost-effectiveness of testing. Applied Economic Analysis 29(85):77–89 (2021). doi:10.1108/AEA-11-2020-0162.

[8] Appleby J. The public finance cost of covid-19. BMJ 376:o490 (2022). doi:10.1136/bmj.o490.

[9] Mizgerd JP. Acute Lower Respiratory Tract Infection. New England Journal of Medicine 358(7):716–727 (2008). doi:10.1056/NEJMra074111.

[10] Malosh RE, Martin ET, Ortiz JR, Monto AS. The risk of lower respiratory tract infection following influenza virus infection: A systematic and narrative review. Vaccine 36(1):141–147 (2018). doi:10.1016/j.vaccine.2017.11.018.

[11] Wardlaw TM, Johansson EW, Hodge MJ. Pneumonia: the forgotten killer of children. UNICEF (2006).

[12] Moore S, Hill EM, Dyson L, Tildesley MJ, Keeling MJ. Modelling optimal vaccination strategy for SARS-CoV-2 in the UK. PLOS Computational Biology 17(5):e1008849 (2021). doi:10.1371/journal.pcbi.1008849.

[13] Moore S, Hill EM, Tildesley MJ, Dyson L, Keeling MJ. Vaccination and non-pharmaceutical interventions for COVID-19: a mathematical modelling study. The Lancet Infectious Diseases 21(6):793–802 (2021). doi:10.1016/S1473-3099(21)00143-2.

[14] Bernal JL, Andrews N, Gower C, Robertson C, Stowe J, et al. Effectiveness of the Pfizer-BioNTech and Oxford-AstraZeneca vaccines on covid-19 related symptoms, hospital admissions, and mortality in older adults in England: test negative case-control study. BMJ 373:n1088 (2021). doi:10.1136/bmj.n1088.

[15] Hall VJ, Foulkes S, Saei A, Andrews N, Oguti B, et al. COVID-19 vaccine coverage in health-care workers in England and effectiveness of BNT162b2 mRNA vaccine against infection (SIREN): a prospective, multicentre, cohort study. The Lancet 397(10286):1725–1735 (2021). doi:10.1016/S0140-6736(21)00790-X.

[16] Shrotri M, Krutikov M, Palmer T, Giddings R, Azmi B, et al. Vaccine effectiveness of the first dose of ChAdOx1 nCoV-19 and BNT162b2 against SARS-CoV-2 infection in residents of long-term care facilities in England (VIVALDI): a prospective cohort study. The Lancet Infectious Diseases 21(11):1529–1538 (2021). doi:10.1016/S1473-3099(21)00289-9.

[17] Chan JFW, Yuan S, Zhang AJ, Poon VKM, Chan CCS, et al. Surgical Mask Partition Reduces the Risk of Noncontact Transmission in a Golden Syrian Hamster Model for Coronavirus Disease 2019 (COVID-19). Clinical Infectious Diseases 71(16):2139–2149 (2020). doi:10.1093/cid/ciaa644.

[18] Gandhi M, Beyrer C, Goosby E. Masks Do More Than Protect Others During COVID-19: Reducing the Inoculum of SARS-CoV-2 to Protect the Wearer. Journal of General Internal Medicine 35(10):3063–3066 (2020). doi:10.1007/s11606-020-06067-8.

[19] Levine Z, Earn DJD. Face masking and COVID-19: potential effects of variolation on transmission dynamics. Journal of The Royal Society Interface 19(190):20210781 (2022). doi:10.1098/rsif.2021. 0781.

[20] Soriano V, Ganado-Pinilla P, Sanchez-Santos M, Gómez-Gallego F, Barreiro P, et al. Main differences between the first and second waves of COVID-19 in Madrid, Spain. International Journal of Infectious Diseases 105:374–376 (2021). doi:10.1016/j.ijid.2021.02.115.

[21] Spinelli MA, Glidden DV, Gennatas ED, Bielecki M, Beyrer C, et al. Importance of non-pharmaceutical interventions in lowering the viral inoculum to reduce susceptibility to infection by SARS-CoV-2 and potentially disease severity. The Lancet Infectious Diseases 21(9):e296–e301 (2021). doi:10.1016/S1473-3099(20)30982-8.

[22] Dennis DT, Gage KL, Gratz N, Poland JD, Tikhomirov E. Plague manual: epidemiology, distribution, surveillance and control. World Health Organisation (1999).

[23] Tellier R. COVID-19: the case for aerosol transmission. Interface Focus 12(2) (2022). doi: 10.1098/rsfs.2021.0072.

[24] Van Damme W, Dahake R, van de Pas R, Vanham G, Assefa Y. COVID-19: Does the infectious inoculum dose-response relationship contribute to understanding heterogeneity in disease severity and transmission dynamics? Medical Hypotheses 146:110431 (2021). doi:10.1016/j.mehy.2020.110431.

[25] Couch RB, Gordon Douglas R, Fedson DS, Kasel JA. Correlated Studies of a Recombinant Influenza-Virus Vaccine. III. Protection against Experimental Influenza in Man. Journal of Infectious Diseases 124(5):473–480 (1971). doi:10.1093/infdis/124.5.473.

[26] Hijano DR, Cardenas JBd, Maron G, Garner CD, Ferrolino JA, et al. Clinical correlation of influenza and respiratory syncytial virus load measured by digital PCR. PLOS ONE 14(9):e0220908 (2019). doi:10.1371/journal.pone.0220908.

[27] Liu Y, Yan LM, Wan L, Xiang TX, Le A, et al. Viral dynamics in mild and severe cases of COVID-19. The Lancet Infectious Diseases 20(6):656–657 (2020). doi:10.1016/S1473-3099(20)30232-2.

[28] Pujadas E, Chaudhry F, McBride R, Richter F, Zhao S, et al. SARS-CoV-2 viral load predicts COVID-19 mortality. The Lancet. Respiratory Medicine 8(9):e70 (2020). doi:10.1016/S2213-2600(20)30354-4.

[29] Han A, Czajkowski LM, Donaldson A, Baus HA, Reed SM, et al. A Dose-finding Study of a Wild-type Influenza A(H3N2) Virus in a Healthy Volunteer Human Challenge Model. Clinical Infectious Diseases 69(12):2082–2090 (2019). doi:10.1093/cid/ciz141.

[30] Miller DS, Kok T, Li P. The virus inoculum volume influences outcome of influenza A infection in mice. Laboratory Animals 47(1):74–77 (2013). doi:10.1258/la.2012.011157.

[31] Imai M, Iwatsuki-Horimoto K, Hatta M, Loeber S, Halfmann PJ, et al. Syrian hamsters as a small animal model for SARS-CoV-2 infection and countermeasure development. Proceedings of the National Academy of Sciences 117(28):16587–16595 (2020). doi:10.1073/pnas.2009799117.

[32] Killingley B, Nguyen-Van-Tam J. Routes of influenza transmission. Influenza and Other Respiratory Viruses 7:42–51 (2013). doi:10.1111/irv.12080.

[33] Tellier R. Aerosol transmission of influenza A virus: a review of new studies. Journal of The Royal Society Interface 6(suppl 6) (2009). doi:10.1098/rsif.2009.0302.focus.

[34] Mathews JD, McCaw CT, McVernon J, McBryde ES, McCaw JM. A Biological Model for Influenza Transmission: Pandemic Planning Implications of Asymptomatic Infection and Immunity. PLoS ONE 2(11):e1220 (2007). doi:10.1371/journal.pone.0001220.

[35] Paulo AC, Correia-Neves M, Domingos T, Murta AG, Pedrosa J. Influenza Infectious Dose May Explain the High Mortality of the Second and Third Wave of 1918–1919 Influenza Pandemic. PLOS ONE 5(7):e11655 (2010). doi:10.1371/journal.pone.0011655.

[36] Atkinson MP, Wein LM. Quantifying the Routes of Transmission for Pandemic Influenza. Bulletin of Mathematical Biology 70(3):820–867 (2008). doi:10.1007/s11538-007-9281-2.

[37] Smieszek T, Lazzari G, Salathé M. Assessing the Dynamics and Control of Droplet-and Aerosol-Transmitted Influenza Using an Indoor Positioning System. Scientific Reports 9(1):2185 (2019). doi:10.1038/s41598-019-38825-y.

[38] Andersson J. What drives transmission of severe acute respiratory syndrome coronavirus 2? Journal of Internal Medicine 290(5):949–951 (2021). doi:10.1111/joim.13335.

[39] Earnest JA. Methods matter: computational modelling in public health policy and planning. PhD, University of Glasgow (2016). URL https://eleanor.lib.gla.ac.uk/record=b3161242.

[40] Harris JD, Park SW, Dushoff J, Weitz JS. How time-scale differences in asymptomatic and symptomatic transmission shape SARS-CoV-2 outbreak dynamics. Epidemics 42:100664 (2023). doi:10.1016/j.epidem.2022.100664.

[41] Hill EM, Petrou S, Forster H, Lusignan Sd, Yonova I, et al. Optimising age coverage of seasonal influenza vaccination in England: A mathematical and health economic evaluation. PLOS Computational Biology 16(10):e1008278 (2020). doi:10.1371/journal.pcbi.1008278.

[42] Thorrington D, van Leeuwen E, Ramsay M, Pebody R, Baguelin M. Assessing optimal use of the standard dose adjuvanted trivalent seasonal influenza vaccine in the elderly. Vaccine 37(15):2051– 2056 (2019). doi:10.1016/j.vaccine.2019.03.002.

[43] Kohli M, Maschio M, Becker D, Weinstein MC. The potential public health and economic value of a hypothetical COVID-19 vaccine in the United States: Use of cost-effectiveness modeling to inform vaccination prioritization. Vaccine 39(7):1157–1164 (2021). doi:10.1016/j.vaccine.2020.12.078.

[44] Neilan AM, Losina E, Bangs AC, Flanagan C, Panella C, et al. Clinical Impact, Costs, and Cost-effectiveness of Expanded Severe Acute Respiratory Syndrome Coronavirus 2 Testing in Massachusetts. Clinical Infectious Diseases 73(9):e2908–e2917 (2021). doi:10.1093/cid/ciaa1418.

[45] Zala D, Mosweu I, Critchlow S, Romeo R, McCrone P. Costing the COVID-19 Pandemic: An Exploratory Economic Evaluation of Hypothetical Suppression Policy in the United Kingdom. Value in Health 23(11):1432–1437 (2020). doi:10.1016/j.jval.2020.07.001.

[46] Anderson RM, May RM. Understanding the AIDS Pandemic. Scientific American 266(5):58–67 (1992).

[47] Keeling M, Rohani P. Modelling Infectious Diseases in Humans and Animals. Princeton University Press (2008).

[48] Little JW, Gordon RD, Hall WJ, Roth FK. Attenuated influenza produced by experimental intranasal inoculation. Journal of Medical Virology 3(3):177–188 (1979). doi:10.1002/jmv.1890030303.

[49] Cowling BJ, Ip DKM, Fang VJ, Suntarattiwong P, Olsen SJ, et al. Aerosol transmission is an important mode of influenza A virus spread. Nature Communications 4(1):1935 (2013). doi: 10.1038/ncomms2922.

[50] Byrne AW, McEvoy D, Collins AB, Hunt K, Casey M, et al. Inferred duration of infectious period of SARS-CoV-2: rapid scoping review and analysis of available evidence for asymptomatic and symptomatic COVID-19 cases. BMJ Open 10(8):e039856 (2020). doi: 10.1136/bmjopen-2020-039856.

[51] Chowell G, Viboud C, Simonsen L, Miller M, Alonso WJ. The reproduction number of seasonal influenza epidemics in Brazil, 1996-2006. Proceedings. Biological Sciences 277(1689):1857–1866 (2010). doi:10.1098/rspb.2009.1897.

[52] Gran JM, Iversen B, Hungnes O, Aalen OO. Estimating influenza-related excess mortality and reproduction numbers for seasonal influenza in Norway, 1975-2004. Epidemiology and Infection 138(11):1559–1568 (2010). doi:10.1017/S0950268810000671.

[53] Gurav YK, Chadha MS, Tandale BV, Potdar VA, Pawar SD, et al. Influenza A(H1N1)pdm09 outbreak detected in inter-seasonal months during the surveillance of influenza-like illness in Pune, India, 2012-2015. Epidemiology and Infection 145(9):1898–1909 (2017). 10.1017/S0950268817000553.

[54] Ward MP, Maftei D, Apostu C, Suru A. Estimation of the basic reproductive number (R0) for epidemic, highly pathogenic avian influenza subtype H5N1 spread. Epidemiology & Infection 137(2):219–226 (2009). doi:10.1017/S0950268808000885. Publisher: Cambridge University Press.

[55] Chowell G, Nishiura H, Bettencourt LM. Comparative estimation of the reproduction number for pandemic influenza from daily case notification data. Journal of The Royal Society Interface 4(12):155–166 (2006). doi:10.1098/rsif.2006.0161. Publisher: Royal Society.

[56] White LF, Wallinga J, Finelli L, Reed C, Riley S, et al. Estimation of the reproductive number and the serial interval in early phase of the 2009 influenza A/H1N1 pandemic in the USA. Influenza and Other Respiratory Viruses 3(6):267–276 (2009). doi:10.1111/j.1750-2659.2009.00106.x. eprint: https://onlinelibrary.wiley.com/doi/pdf/10.1111/j.1750-2659.2009.00106.x.

[57] D’Arienzo M, Coniglio A. Assessment of the SARS-CoV-2 basic reproduction number, R0, based on the early phase of COVID-19 outbreak in Italy. Biosafety and Health 2(2):57–59 (2020). doi:10.1016/j.bsheal.2020.03.004.

[58] Petersen E, Koopmans M, Go U, Hamer DH, Petrosillo N, et al. Comparing SARS-CoV-2 with SARS-CoV and influenza pandemics. The Lancet Infectious Diseases 20(9):e238–e244 (2020). doi:10.1016/S1473-3099(20)30484-9. Publisher: Elsevier.

[59] Liu Y, Gayle AA, Wilder-Smith A, Rocklöv J. The reproductive number of COVID-19 is higher compared to SARS coronavirus. Journal of Travel Medicine 27(2):taaa021 (2020). 10.1093/jtm/taaa021.

[60] Cao B, Li XW, Mao Y, Wang J, Lu HZ, et al. Clinical Features of the Initial Cases of 2009 Pandemic Influenza A (H1N1) Virus Infection in China. New England Journal of Medicine 361(26):2507–2517 (2009). doi:10.1056/NEJMoa0906612.

[61] Letizia AG, Ramos I, Obla A, Goforth C, Weir DL, et al. SARS-CoV-2 Transmission among Marine Recruits during Quarantine. New England Journal of Medicine 383(25):2407–2416 (2020). doi:10.1056/NEJMoa2029717.

[62] Hardelid P, Fleming DM, McMenamin J, Andrews N, Robertson C, et al. Effectiveness of pandemic and seasonal influenza vaccine in preventing pandemic influenza A(H1N1)2009 infection in England and Scotland 2009-2010. Eurosurveillance 16(2):19763 (2011). doi:10.2807/ese.16.02.19763-en.

[63] Pebody RG, Andrews N, Fleming DM, McMENAMIN J, Cottrell S, et al. Age-specific vaccine effectiveness of seasonal 2010/2011 and pandemic influenza A(H1N1) 2009 vaccines in preventing influenza in the United Kingdom. Epidemiology & Infection 141(3):620–630 (2013). doi:10.1017/S0950268812001148.

[64] Andrews N, McMenamin J, Durnall H, Ellis J, Lackenby A, et al. Effectiveness of trivalent seasonal influenza vaccine in preventing laboratory-confirmed influenza in primary care in the United Kingdom: 2012/13 end of season results. Eurosurveillance 19(27):20851 (2014). doi: 10.2807/1560-7917.ES2014.19.27.20851.

[65] Kaplan RM. Utility assessment for estimating quality-adjusted life years. In FA Sloan, editor, Valuing Health Care, pages 31–60. Cambridge University Press, 1 edition (1995). ISBN 978-0-521-57646-8 978-0-521-47020-9 978-0-511-62581-7. doi:10.1017/CBO9780511625817.003. URL https://www.cambridge.org/core/product/identifier/CBO9780511625817A019/type/book_part.

[66] Hollmann M, Garin O, Galante M, Ferrer M, Dominguez A, et al. Impact of Influenza on Health-Related Quality of Life among Confirmed (H1N1)2009 Patients. PLoS ONE 8(3):e60477 (2013). doi:10.1371/journal.pone.0060477.

[67] National Institute for Health and Care Excellence. Guide to the processes of technology appraisal (2018).

[68] Moran DP, Pires SM, Wyper GMA, Devleesschauwer B, Cuschieri S, et al. Estimating the Direct Disability-Adjusted Life Years Associated With SARS-CoV-2 (COVID-19) in the Republic of Ireland: The First Full Year. International Journal of Public Health 0 (2022). doi:10.3389/ijph.2022.1604699r.

[69] Vekaria B, Overton C, Wisniowski A, Ahmad S, Aparicio-Castro A, et al. Hospital length of stay for COVID-19 patients: Data-driven methods for forward planning. BMC Infectious Diseases 21(1):700 (2021). doi:10.1186/s12879-021-06371-6.

[70] National Institute for Health and Care Excellence. Guide to the processes of technology appraisal (2018). URL https://www.nice.org.uk/Media/Default/About/what-we-do/NICE-guidance/NICE-technology-appraisals/technology-appraisal-processes-guide-apr-2018.pdf. [Online] (Accessed: 24 January 2024).

[71] National Institute for Health and Care Excellence. Discounting task and finish group report (2020). URL https://www.google.com/url?sa=t&rct=j&q=&esrc=s&source=web&cd=&ved=2ahUKEwjS_aO10_r_AhXqTkEAHQ7LDOkQFnoECBIQAQ& url=https://www.nice.org.uk%2FMedia%2FDefault%2FAbout%2Fwhat-we-do%2Four-programmes%2Fnice-guidance%2Fchte-methods-consultation%2FDiscounting-task-and-finish-group-report.docx&usg=AOvVaw0W-X_CPsnuXp6EZBvSrDNY&opi=89978449. [Online] (Accessed: 24 January 2024).

[72] Furuya-Kanamori L, Cox M, Milinovich GJ, Magalhaes RJS, Mackay IM, et al. Heterogeneous and Dynamic Prevalence of Asymptomatic Influenza Virus Infections. Emerging Infectious Diseases 22(6):1052–1056 (2016). doi:10.3201/eid2206.151080.

[73] Wang B, Andraweera P, Elliott S, Mohammed H, Lassi Z, et al. Asymptomatic SARS-CoV-2 Infection by Age: A Global Systematic Review and Meta-analysis. Pediatric Infectious Disease Journal 42(3):232–239 (2023). doi:10.1097/INF.0000000000003791.

[74] Stockmaier S, Stroeymeyt N, Shattuck EC, Hawley DM, Meyers LA, et al. Infectious diseases and social distancing in nature. Science 371(6533):eabc8881 (2021). doi:10.1126/science.abc8881.

[75] Antonelli M, Penfold RS, Merino J, Sudre CH, Molteni E, et al. Risk factors and disease profile of post-vaccination SARS-CoV-2 infection in UK users of the COVID Symptom Study app: a prospective, community-based, nested, case-control study. The Lancet Infectious Diseases 22(1):43–55 (2022). doi:10.1016/S1473-3099(21)00460-6.

[76] Bielecki M, Zust R, Siegrist D, Meyerhofer D, Crameri GAG, et al. Social Distancing Alters the Clinical Course of COVID-19 in Young Adults: A Comparative Cohort Study. Clinical Infectious Diseases 72(4):598–603 (2021). doi:10.1093/cid/ciaa889.

[77] Sullivan SG, Feng S, Cowling BJ. Potential of the test-negative design for measuring influenza vaccine effectiveness: a systematic review. Expert Review of Vaccines 13(12):1571–1591 (2014). doi:10.1586/14760584.2014.966695.

[78] UK Health Security Agency. COVID-19 vaccination programme information for healthcare practitioners (2023). URL https://www.gov.uk/government/publications/covid-19-vaccination-programme-guidance-for-healthcare-practitioners. [Online] (Accessed: 24 January 2024).

## Supplementary References

[1] Diekmann O, Heesterbeek J, Metz J. On the definition and the computation of the basic reproduction ratio R 0 in models for infectious diseases in heterogeneous populations. Journal of Mathematical Biology 28(4) (1990). doi:10.1007/BF00178324.

[2] Hill EM, Petrou S, Forster H, Lusignan Sd, Yonova I, et al. Optimising age coverage of seasonal influenza vaccination in England: A mathematical and health economic evaluation. PLOS Computational Biology 16(10):e1008278 (2020). doi:10.1371/journal.pcbi.1008278. Publisher: Public Library of Science.

[3] Moran DP, Pires SM, Wyper GMA, Devleesschauwer B, Cuschieri S, et al. Estimating the Direct Disability-Adjusted Life Years Associated With SARS-CoV-2 (COVID-19) in the Republic of Ireland: The First Full Year. International Journal of Public Health 0 (2022). doi:10.3389/ijph.2022.1604699.

[4] Hollmann M, Garin O, Galante M, Ferrer M, Dominguez A, et al. Impact of Influenza on Health-Related Quality of Life among Confirmed (H1N1)2009 Patients. PLoS ONE 8(3):e60477 (2013). doi:10.1371/journal.pone.0060477.

[5] Vekaria B, Overton C, Wísniowski A, Ahmad S, Aparicio-Castro A, et al. Hospital length of stay for COVID-19 patients: Data-driven methods for forward planning. BMC Infectious Diseases 21(1):700 (2021). doi:10.1186/s12879-021-06371-6.

[6] Lina B, Georges A, Burtseva E, Nunes MC, Andrew MK, et al. Complicated hospitalization due to influenza: results from the Global Hospital Influenza Network for the 2017–2018 season. BMC Infectious Diseases 20(1):465 (2020). doi:10.1186/s12879-020-05167-4.

[7] Smith, DH, Gravelle. The practice of discounting in economic evaluations of healthcare interventions. Int J Technol Assess Health Care 17(2):236–243 (2001). doi:10.1017/s0266462300105094.

